# Cost-effective boosting allocations in the post-Omicron era of COVID-19 management

**DOI:** 10.1101/2023.11.14.23298536

**Authors:** Thao P. Le, Eamon Conway, Edifofon Akpan, Isobel Abell, Patrick Abraham, Christopher M. Baker, Patricia T. Campbell, Deborah Cromer, Michael J. Lydeamore, Yasmine McDonough, Ivo Mueller, Gerard Ryan, Camelia Walker, Yingying Wang, Natalie Carvalho, Jodie McVernon

## Abstract

**Background:** Following widespread exposure to Omicron variants, COVID-19 has transitioned to endemic circulation. Populations now have diverse infection and vaccination histories, resulting in heterogeneous immune landscapes. Careful consideration of vaccination is required through the post-Omicron phase of COVID-19 management to minimise disease burden. We assess the impact and cost-effectiveness of targeted COVID-19 vaccination strategies to support global vaccination recommendations.

**Methods:** We integrated immunological, transmission, clinical and cost-effectiveness models, and simulated populations with different characteristics and immune landscapes. We calculated the expected number of infections, hospitalisations and deaths for different vaccine scenarios. Costs (from a healthcare perspective) were estimated for exemplar country income level groupings in the Western Pacific Region. Results are reported as incremental costs and disability-adjusted life years averted compared to no additional vaccination. Parameter and stochastic uncertainty were captured through scenario and sensitivity analysis.

**Findings:** Across different population demographics and income levels, we consistently found that annual elder-targeted boosting strategies are most likely to be cost-effective or cost-saving, while paediatric programs are unlikely to be cost-effective. Results remained consistent while accounting for uncertainties in the epidemiological and economic models. Half-yearly boosting may only be cost-effective in higher income settings with older population demographics and higher cost-effectiveness thresholds.

**Interpretation:** The seresults demonstrate the value of continued booster vaccinations to protect against severe COVID-19 disease outcomes across high and middle-income settings and show that the biggest health gains relative to vaccine costs are achieved by targeting older age-groups.

**Funding:** Funded by the World Health Organization.

**Research in context**

Evidence before this study
With COVID-19 now globally endemic, populations exhibit varying levels of natural and vaccine-acquired immunity to SARS-CoV-2. With widespread, if variable, immunity resulting in reduced severity of COVID-19 disease, re-evaluation of the ongoing value of vaccination is required. COVID-19 vaccination strategies must consider the cost-effectiveness of gains from vaccination given prior immunity, and in the context of income and health system capacity to manage COVID-19 alongside other pressing concerns.
Few articles examine cost-effectiveness of COVID-19 vaccination strategies in populations with diverse characteristics and waning hybrid immunity, though there is a large body of literature that considers some combination of these elements or focus on one particular country. Consensus is that allocating vaccine doses to older age groups and those at higher risk of severe disease is most beneficial, albeit assuming either only past natural immunity or no waning immunity. These studies have either not included a cost-effectiveness analysis or, where present, have typically assumed a base case zero-vaccination scenario.

Added value of this study
We consider the contemporary situation where populations have varying degrees of hybrid immunity resulting from both prior infection and vaccination, and where the relevant cost-effectiveness analysis considers only future primary and booster doses in the population. We describe multiple demographics, using exemplar ‘older’ and ‘younger’ populations, in conjunction with low to high past vaccination coverage, low to high past natural infection incidence, and low to high income levels. Under these settings, we determine the cost-effectiveness of a range of targeted boosting strategies (who, when, what).

Implications of all the available evidence
Our study highlights how future COVID-19 booster doses targeted towards older age groups at risk of severe outcomes can be cost-effective or cost-saving in high-income settings with populations that have a higher proportion of individuals at risk. In younger, lower-resourced settings, annual boosting of older age groups may still be cost-effective or cost-saving in some scenarios. We consistently find that pediatric vaccination is not cost-effective. Given the benefits of vaccination, especially to reduce severe disease, we show the importance of ongoing global efforts to provide and equitably distribute vaccines and strengthen adult immunisation programs.

## 1. Introduction

Since the emergence of SARS-CoV-2 in late 2019, the world has experienced multiple epidemic waves of COVID-19 disease and diverse evolutionary variants of SARS-CoV-2.^1^ In parallel to the evolution of the virus, a range of COVID-19 vaccines have been developed,^2^ and there have been multiple rounds of vaccination across the world.

Both prior infection and vaccination can reduce the chance of future infection and severity of outcomes, combining to form “hybrid immunity”^3^ against COVID-19. In the post-Omicron era, most populations have high levels of past infection across multiple epidemic waves, creating exposure-derived ‘natural’ immunity. Vaccine coverage has been variable due to inequities of access, eligibility and uptake, with consequences for hybrid immunity landscapes. ^4;5;6^ Unfortunately, all forms of immunity wane over time, enabling possible reinfection within a matter of months, though protection against severe outcomes is longer lived. ^6;7^

How can COVID-19 vaccines incorporated into routine immunisation schedules help minimise the impact of recurring epidemic waves and promote resilience against future variants? Heterogeneous population experiences of infection and vaccination, and the irregular emergence of immune escape variants, makes anticipating the timing, magnitude and clinical burden of future epidemics challenging. The World Health Organization (WHO) recently reviewed evidence for the impact and cost-effectiveness of COVID-19 booster vaccine strategies. Key questions to inform strategic guidelines included: the incremental benefits of boosters, identification of optimal vaccine target groups in high seroprevalence settings, ongoing vaccine-preventable disease burden, optimum boosting strategy including frequency for priority populations, and the cost effectiveness of those vaccination strategies. We need flexible frameworks to investigate this multi-dimensional problem space.

As one of several groups commissioned by WHO to support decision making, we adapted an existing model representing diverse population and hybrid immunity landscapes^5^ to address questions relevant to future COVID-19 vaccine prioritisation. ^8^ We mimicked realistic epidemic exposure histories and specified timings for the emergence of immune escape variants. We also proposed plausible vaccine rollout and coverage assumptions. We developed exemplar demographies and costings based on high and low-middle income countries in the Western Pacific Region. Our findings played a role in informing the WHO updated COVID-19 vaccination guidance for March 2023. ^9^

## 2. Methods

Our modelling pipeline is depicted in Figure 1. First, the immunological model informs an infection transmission/dynamics model within a mechanical agent-based model. The outputs of the agent-based model are input to a clinical pathways model to obtain clinical outcomes. These clinical outcomes then link to a cost-effectiveness model evaluating alternative vaccination strategies. Based on the problem space, we configure our model using numerous parameters, including population distribution, vaccine program and health systems costs for exemplar country contexts.

**Figure 1:**
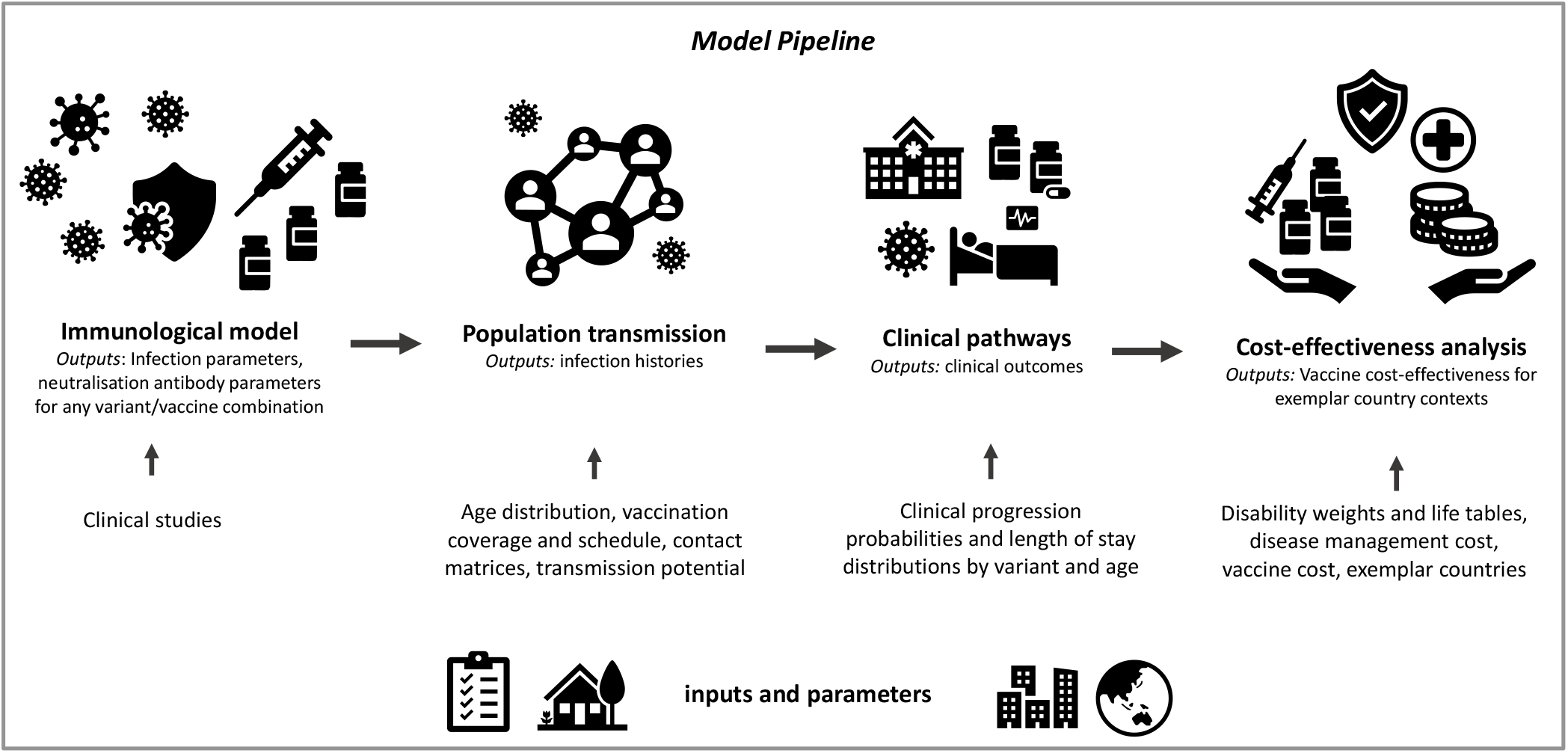
Simulation and analysis model framework, from immunological model, to cost-effectiveness analysis. The immunological model converts vaccinations and infections into neutralising antibody parameters, which influence outbreak dynamics and clinical outcomes. The simulation model has two parts: first an agent-based model of population level transmission, with multiple primary inputs including vaccination and demography, followed by a clinical pathways model, which generates outcomes using the timeseries of symptomatic infections, neutralising antibodies and age-dependent clinical progression probabilities. The resultant clinical outcomes are used in the cost-effectiveness analysis, along with other input parameters such as vaccine and disease management costs.

Our methods extend previous work^5^ with adjusted scenarios to answer policy-relevant questions, and include cost-effective analyses. Full model details can be found in the appendix and references.^5;10^

### 2.1. Immunological model

Our immunological model is based on the established correlation between neutralising antibodies and protection against COVID-19^11;12^. We use the model developed by Khoury et al.^11^ and Cromer et al.^12^ as implemented by Golding et al.^13^. This model maps the dynamics of antibodies over the first 250 days since an immune event to protection from disease, including infection acquisition, symptomatic disease, hospitalisation, and death. The model includes the efficacy of a range of vaccines in the presence and absence of infection.^13^ For this work, we assume a fixed decay rate. See Table B1 in the appendix for the immunological parameters.

Evolutionary variants interact with our model through changes in the baseline transmissibility of the virus, and through their ability to ‘escape’ host immunity. While many early variants had transmission advantages over their predecessors, the Omicron variant and its sub-lineages developed significant ability to escape host immunity to other variants. Our model’s parameters are defined to incorporate the properties of the Delta variant. We then incorporate an ‘escape parameter’ to account for the difference between the Omicron BA.1/2-like variant and the Delta variant, and another ‘escape parameter’ for difference between the Omicron BA.4/5-like Variant and Omicron BA.1/2. We estimated escape as a latent parameter in a model based on reinfection and reproduction of the Omicron and Delta variants in South Africa.^13^ Functionally, this escape parameter results in reducing effective individual neutralisation titre against new variants.

For bivalent vaccines, Khoury et al.^7^ found that variant adapted vaccines produced, on average, 1.61-fold higher titers than ancestral vaccines. We therefore implement bivalent boosters by using this multiplier on top of the neutralisation titres given by an ancestral mRNA booster vaccine.

### 2.2. Population transmission model

We model the transmission of COVID-19 with an agent-based model adapted from the work of Conway et al.^10;14^ Each simulated individual has their own neutralising antibodies, age, and history of vaccination and infection exposure. Transmission is simulated by directly modelling contact between infectious and susceptible individuals, where the probability of transmission, infection and symptomatic disease is determined by neutralising antibody levels.

We configured two distinct populations, representing typical ‘younger’ and ‘older’ demographics found within the Western Pacific Region.^5^

We implement baseline hybrid immunity through the first 1.5 years by rolling out vaccinations and introducing infection transmission. Our populations have one of three past vaccination coverage levels, serving as a proxy for both health system capacity and access: low (20% coverage), medium (50% coverage) and high (80% coverage). We introduce Omicron BA1/2 circulation at around 7 months to develop natural immunity.

Note that populations have either low (15%-45%) or high (80%-100%) past attack rates, corresponding to low or high seroprevalence. We implemented this by varying the transmission potential parameter. This accounts for other factors that can affect transmission potential which we do not explicitly model, such as climate, housing, and population density.^15^

After the baseline hybrid immunity is achieved, we test different boosting vaccination strategies between 1.5–3 years, detailed in Section 2.5. The timing of emergence of the immune escape variant (Omicron BA4/5), at either 1.5 years, 2 years, or 2.5 years is also varied. This allows the observation of resurgent epidemic waves driven by waning and/or immune escape. The immune escape variant has a transmission potential multiplier (1.3) on top of the baseline transmission potential.

### 2.3. Clinical pathways

The clinical pathways model is based on the framework proposed by Knock et al.^16^. We extend this model, building upon previous work,^14;17^ to transform it into an agent-based model, and utilise the ages and neutralising antibody titres of individuals from the population transmission model to generate clinical trajectories.^10^ The outcomes for individuals include whether they experienced severe disease, required intensive care unit (ICU) admission, or died. The duration of hospitalisation in general wards and ICU follow a Gamma distribution, with means and variances sampled from estimates during the Australian Omicron outbreak.^18^ Details of the clinical pathways model are given here.^10^.

Clinical outcomes are age-dependent. This key assumption, informed by clinical data, suggests that older age groups are at higher risk of severe disease and could have greater benefits from protective vaccination. This premise significantly contributes to our results, particularly regarding cost-effectiveness of vaccination in different population demographics.

### 2.4. Cost-effectiveness analysis

The cost-effectiveness model uses outputs from the clinical pathways model. The cost-effectiveness analysis is conducted from a healthcare system perspective, including the following direct medical cost categories:

1. Programmatic costs related to the vaccination intervention, including vaccine dose costs, wastage, and delivery costs; and
2. Disease management costs at home, in outpatient and inpatient settings for symptomatic COVID-19 related illness.

Costs are estimated for exemplar countries in the Western Pacific Region. We categorize exemplar countries into three distinct groupings with different demographics, health system strength, income levels, and vaccine coverage: Group A (high-income, older population, strong health systems, high vaccination coverage); Group B (upper and lower middle-income, younger population, varied health systems, medium or high vaccination coverage); and Group C (lower-middle income, younger population, weak health systems, low vaccination coverage). Details of the specific costings used in this work are in Table 1.

**Table 1.** Source of costings for the cost-effectiveness analysis model. Country groups refer to Group A (high-income, older population, strong health systems, high vaccination coverage); Group B (upper and lower middle-income, younger population, varied health systems, medium or high vaccination coverage); and Group C (lower-middle income, younger population, weak health systems, low vaccination coverage). Full costing details are contained in appendix Section D.2.

| Cost | Source | Notes |
| --- | --- | --- |
| **Vaccine dose prices** | WHO COVID-19 Vaccine Price Report | Assuming 10% dose wastage rate from UNICEF reports |
| **Vaccine delivery costs** |  |  |
| Group A | Governmental reports and peer-reviewed studies |  |
| Group B | Griffiths et al. <sup>19,20</sup> |  |
| Group C | Same as Group B, except doubled at 20% coverage | No country-specific estimates at 20% coverage |
| **Disease management costs** |  |  |
| Group A | Publicly available medical fee schedules, published studies, WHO-CHOICE database |  |
| Group B/C | Torres-Rueda et al. <sup>21</sup> |  |
| **COVID-19 related deaths** | Torres-Rueda et al. <sup>21</sup> | Only includes cost of body bags |

Health outcomes are presented as disability-adjusted life years using disability weights from the Global Burden of Disease study and Japanese disability weight measurement studies.^22;23^ Duration of illness estimates based on illness severity are from previous studies,^24;25;26;27^ and estimates of life years lost due to premature mortality are from WHO life tables for each 10-year age band. We use the Japan life table for Group A and the global lower-middle income life table for Groups B and C.^28^ Future costs and health outcomes are discounted by 3%. We report costs in 2020 United States dollars (USD).

We present cost-effectiveness results of each vaccination scenario as incremental cost effectiveness ratios (ICERs) compared to a ‘do-no-further-vaccination’ scenario. These ICERs are compared to a range of recently proposed country specific cost-effectiveness thresholds (CETs) based on health opportunity costs.^29^ We adapted CETs based on 2020 GDP per capita data from the World Bank. Average CETs for Group A are $19,000-$30,000, $200-$1,600 for Group B, and $100-$1,000 for Group C. If a scenario’s ICER falls below the thresholds provided, it is considered likely to be cost-effective.

Deterministic sensitivity analysis and probabilistic sensitivity analysis were conducted to consider the uncertainty in parameters, including reduced care-seeking and/or access to donated vaccines in lower-income settings (see appendix Section E).

### 2.5. Vaccination scenarios

#### 2.5.1. High vaccination coverage scenarios

In high coverage settings (older Group A and younger Group B demographics) we consider three boosting strategies: pediatric boosting (ages 5-15), high-risk boosting (65+ in the older population and 55+ in the younger population), and random boosting. We fix the number of vaccine doses (11, 000) across these scenarios to focus on the impact of vaccine allocation.

We also explore the timing and frequency of high-risk boosting, with boosting occurring at 1.75, 2.0, 2.25, or 2.5 years, or every 6 months starting from 1.75 years.

We also compare additional boosting strategies, where we boost the 65+ age group and expand booster eligibility to younger age groups. This allows us to test the limits of cost effectiveness of extending coverage to lower-risk groups. We do not fix the number of vaccine doses across these scenarios.

#### 2.5.2. Low-medium vaccination coverage scenarios

When primary coverage is lower (younger demographic countries in Group B with medium coverage and Group C with low coverage), we explore three vaccination strategies: new pediatric primary vaccination, high risk boosting (older first), and new random primary vaccinations. We fix the number of vaccine doses (11, 000) across these scenarios.

We also consider the impact of switching from monovalent to bivalent vaccines, given that bivalent vaccines are being administered globally.^2;7^ We anticipate that bivalent boosting would have the biggest impact in populations with relatively low vaccine and infection derived immunity.^7^

## 3. Results

### 3.1. High vaccination coverage scenarios

#### 3.1.1. Comparing target use groups

Figure 2 compares the impacts of alternative vaccine allocations in older and younger populations depending on the time of immune escape emergence (1.5 vs 2.5 years), given past (1.5 years) high seroprevalence. We find limited impact of different strategies on infection dynamics. In the late (2.5 years) immune escape scenarios (panels (c) and (d)), the epidemic peaks are shifted to the right, but overall the infection curves maintain the same qualitative shape.

**Figure 2:**
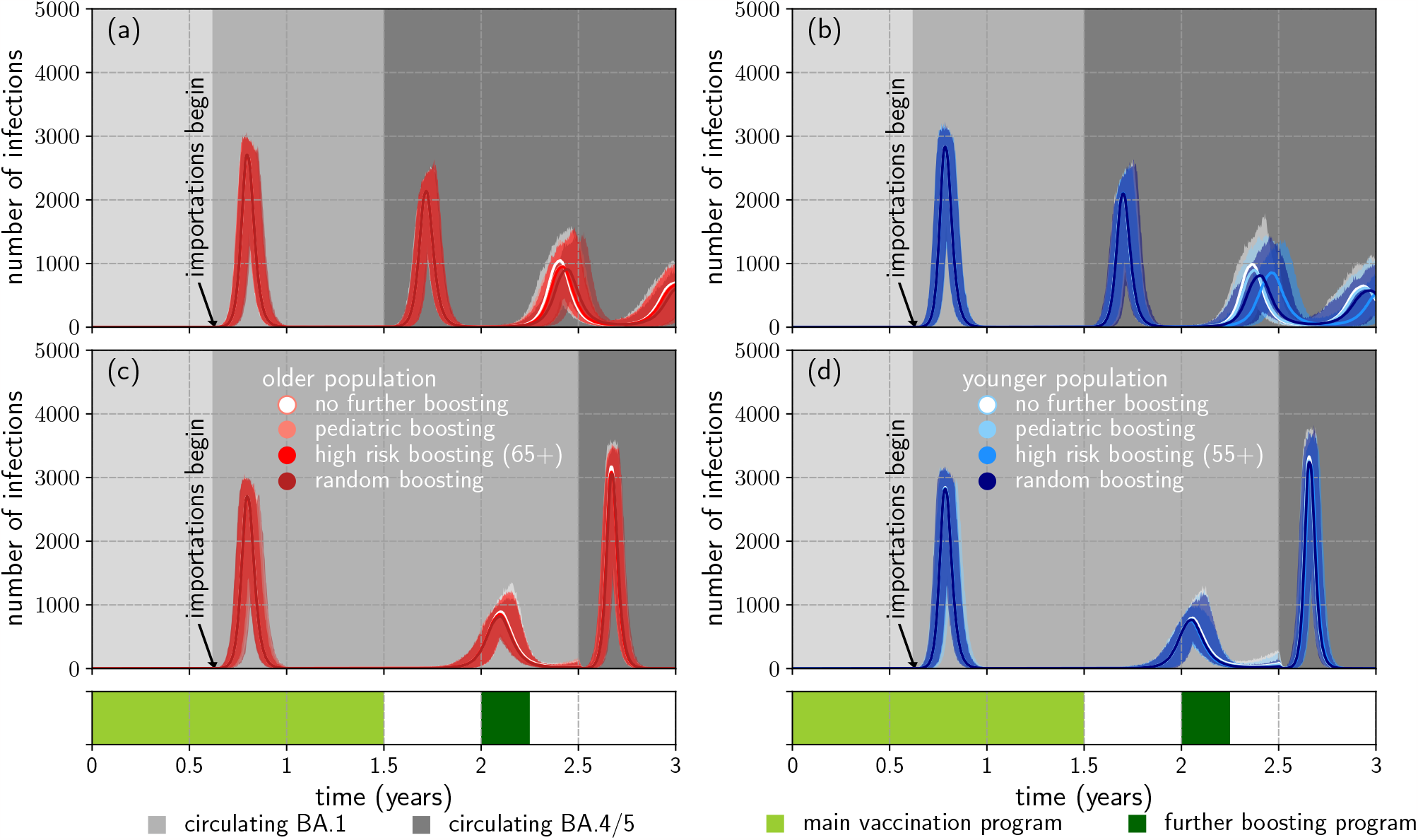
Outbreaks in high transmission settings with high vaccination coverage, for older and younger demographics with early and late seeding of an immune evading variant (dark grey shading). **(a)** older population with early immune escape (1.5 years); **(b)** younger population with early immune escape (1.5 years); **(c)** older population with late immune escape (2.5 years); **(d)** younger population with late immune escape (2.5 years). Scenarios **(a)**-**(d)** are run with strategies of no further boosting, pediatric boosting (ages 5-15), high-risk boosting (65+ in the older population, 55+ in the younger population) and random boosting at 2 years. Scenarios are presented with lines representing pointwise medians from 1000 simulations and shaded regions representing the minimum and maximum from the simulations. The medium grey and dark grey background define the currently circulating variant, Omicron BA.1-like and BA.4/5-like respectively. The impact of boosting on infections is limited.

We find that vaccination has greater impact on severe outcomes. Across the scenarios, high-risk boosting averts the most severe disease (Table 2).

**Table 2.** Median deaths between 1.5 - 3 years in high transmission settings with high vaccination coverage from 5000 simulations (5 clinical pathway simulations are produced per infection transmission simulation), with 0.025 and 0.975 quantiles. Early immune escape occurs at 1.5 years; late immune escape occurs at 2.5 years. Each scenario is run with four different boosting strategies at 2 years.

| Population demographics | Immune escape | Boosting strategy | Median deaths |
| --- | --- | --- | --- |
| (a) older | early | no further boosting | 40.0 (28.0, 53.0) |
|  |  | pediatric (ages 5-15) | 40.0 (29.0, 53.0) |
|  |  | high-risk (65+) | 34.0 (23.0, 46.0) |
|  |  | random | 39.0 (27.0, 52.0) |
| (b) younger | early | no further boosting | 11.0 (5.0, 18.0) |
|  |  | pediatric (ages 5-15) | 12.0 (6.0, 18.0) |
|  |  | high-risk (55+) | 9.0 (4.0, 15.0) |
|  |  | random | 11.0 (5.0, 18.0) |
| (c) older | late | no further boosting | 49.0 (36.0, 62.0) |
|  |  | pediatric (ages 5-15) | 49.0 (36.0, 62.0) |
|  |  | high-risk (65+) | 34.0 (23.0, 47.0) |
|  |  | random | 44.0 (32.0, 59.0) |
| (d) younger | late | no further boosting | 15.0 (8.0, 24.0) |
|  |  | pediatric (ages 5-15) | 17.0 (9.0, 25.0) |
|  |  | high-risk (55+) | 9.0 (4.0, 16.0) |
|  |  | random | 15.0 (8.0, 23.0) |

In Figure 3(a), we find that in older populations, boosting is more cost-effective when it occurs prior to immune escape. High-risk boosting is likely to be highly cost-effective or cost-saving, while pediatric boosting does not appear to be cost-effective. Random boosting does worse than high-risk boosting. Cost-effectiveness of high-risk boosting is driven primarily by vaccine program (delivery and dose) costs, followed by disease management costs in general ward (see Fig. E5 in the appendix).

**Figure 3:**
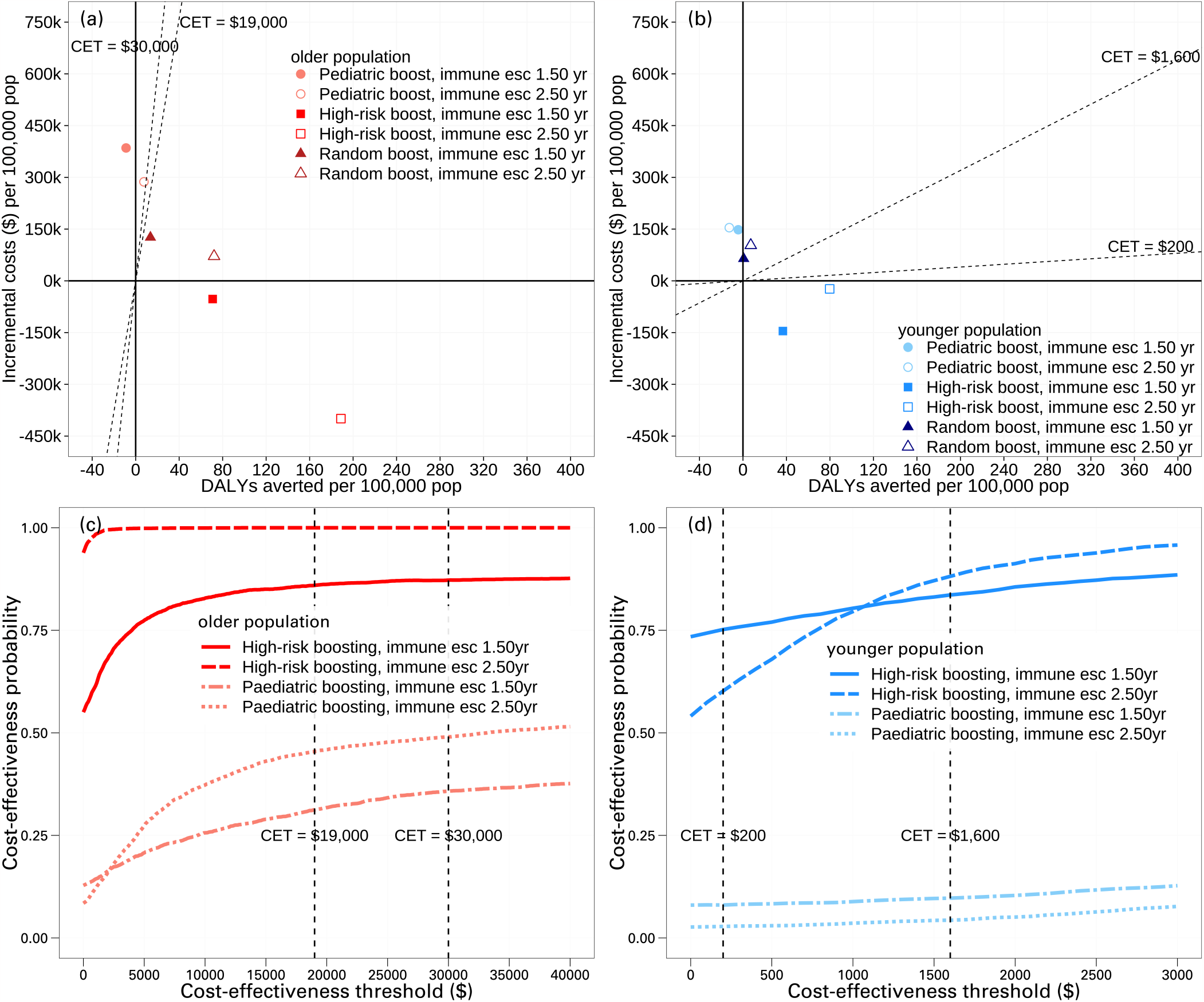
Cost-effectiveness results for different vaccination strategies in high transmission high vaccination coverage settings, for older (Group A) and younger (Group B) demographics with early (1.50 year) and late (2.50 year) seeding of an immune evading variant. Top figures represent cost-effectiveness planes for **(a)** older population and **(b)** younger population. Bottom figures show cost-effectiveness acceptability curves considering stochastic uncertainty and economic parameter uncertainty for the most cost-effective high-risk strategies for **(c)** older population and **(d)** younger population. The boosting strategies considered here are: pediatric boosting (ages 5-15), high-risk boosting (65+ in the older population, 55+ in the younger population) and random boosting at 2 years. High risk boosting is the most cost-effective strategy and dominates pediatric and random boosting strategies.

High risk boosting *may* be cost-effective or even cost-saving in younger MIC populations, depending on country-level willingness to pay (WTP) per DALY averted threshold, or CET, and other key model inputs, which we see in Figure 3(right side). These results are driven primarily by home-based care cost inputs, and vaccine delivery costs, which remain highly uncertain in these settings (Fig. E6 in the appendix). In sensitivity analyses investigating no home-based care costs in these settings, we find elder boosting strategies may remain cost-effective when vaccine is donated and for countries with higher CETs (Figure E7 in the appendix).

The relative benefits of boosting, especially high-risk boosting, hold across all the scenarios considered in Figures 2, 3 and Table 2. However, the absolute benefits are greater in the older population.

Note that we assume high transmission potential, which leads to high seroprevalence. The relative impact of vaccination in the low seroprevalence scenario is similar (appendix Section E.1).

#### 3.1.2. Boosting frequency

The timing and frequency of boosting, relative to emergence of the immune escape variant, influenced the impact of vaccination on the number of infections (Figure 4). Half-yearly boosting consistently achieves fewer severe outcomes when compared to boosting once (see appendix Table E3).

**Figure 4:**
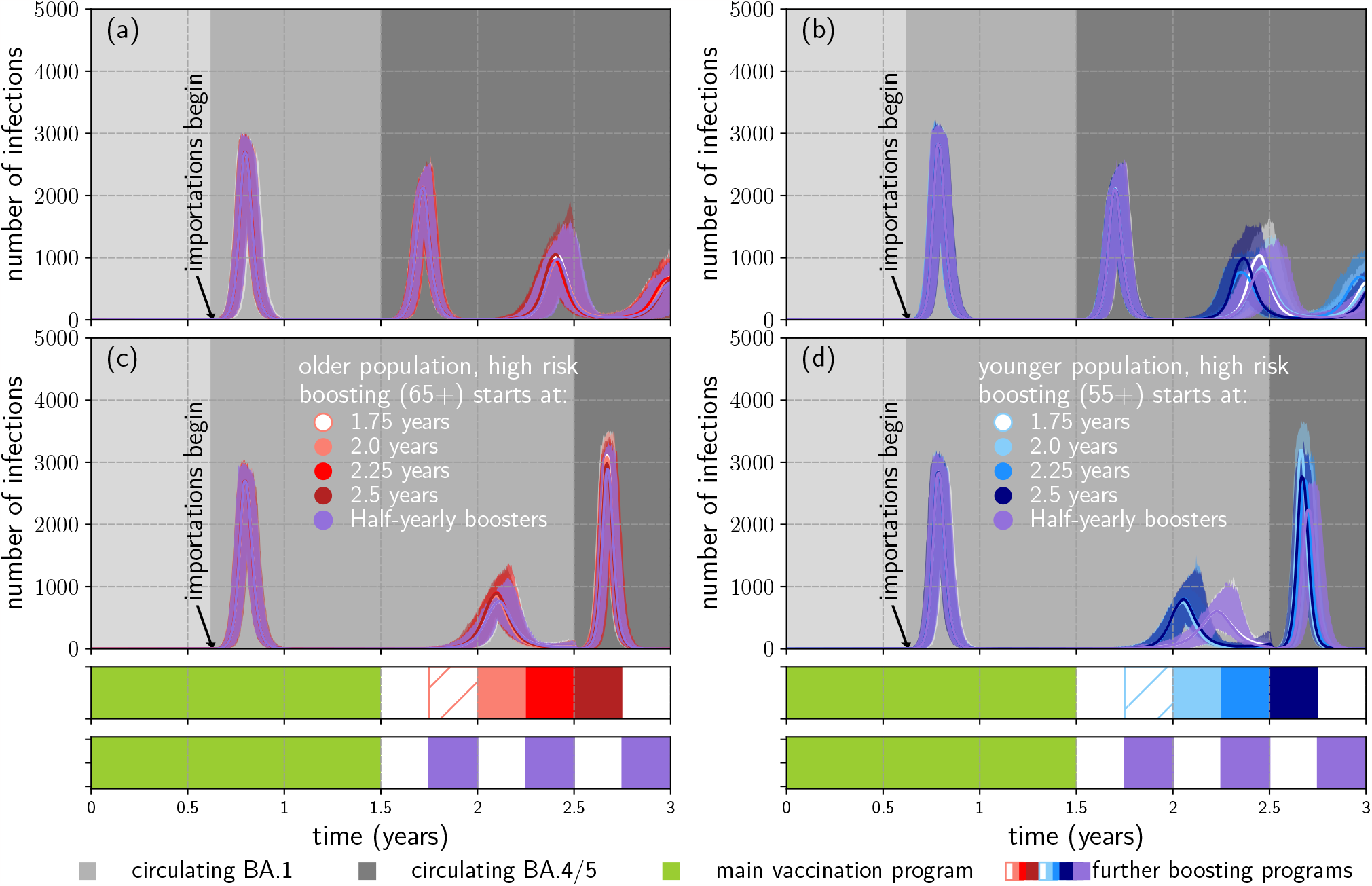
Outbreaks in high transmission settings with high vaccination coverage, for older and younger demographics, comparing boosting once at a range of times and frequent boosting. (a) older population with early immune escape (1.5 years); (b) younger population with early immune escape (1.5 years); (c) older population with late immune escape (2.5 years); (d) younger population with late immune escape (2.5 years). Scenarios **(a)-(d)** are run with high-risk boosting (65+ in the older population, 55+ in the younger population) rolled out at either 1.75 years, 2.0 years, 2.25 years, 2.5 years, or half-yearly starting from 1.75 years. Scenarios are presented with lines representing pointwise medians from 1000 simulations and shaded regions representing the minimum and maximum from the simulations. The timing and frequency of boosting, relative to emergence of the immune escape variant, influences the impact of vaccination on the subsequent outbreaks.

The cost-effectiveness of high-risk boosting varies according to immune escape timing, but generally is very cost-effective or cost-saving in the older (HIC) population and in younger (MIC) populations. Half-yearly boosting remains highly cost-effective in older populations, but more expensive than boosting only once (see appendix Figure E8).

However, half-yearly boosting is unlikely to be cost-effective for younger (MIC) countries with high vaccine coverage (∼ 80%) unless vaccines are donated. These results were driven by home-based care costs (lower costs indicate half-yearly boosting is unlikely to be cost-effective) and vaccine program costs (lower costs, for example, donated vaccine, would mean half-yearly boosting may be very cost-effective or cost-saving) (see appendix Figure E9).

#### 3.1.3. Age cut-off for cost-effective boosting

We systematically expanded booster eligibility to younger age groups (Figure 5). The difference in health outcomes between boosting 45+ and younger age groups is minimal (see appendix Table E4), but increasing the age cohorts in the program leads to higher costs and thus lower cost effectiveness. Boosting 65+ and 55+ is likely highly cost-effective or cost-saving. Boosting 45+ is also likely highly cost-effective.

**Figure 5:**
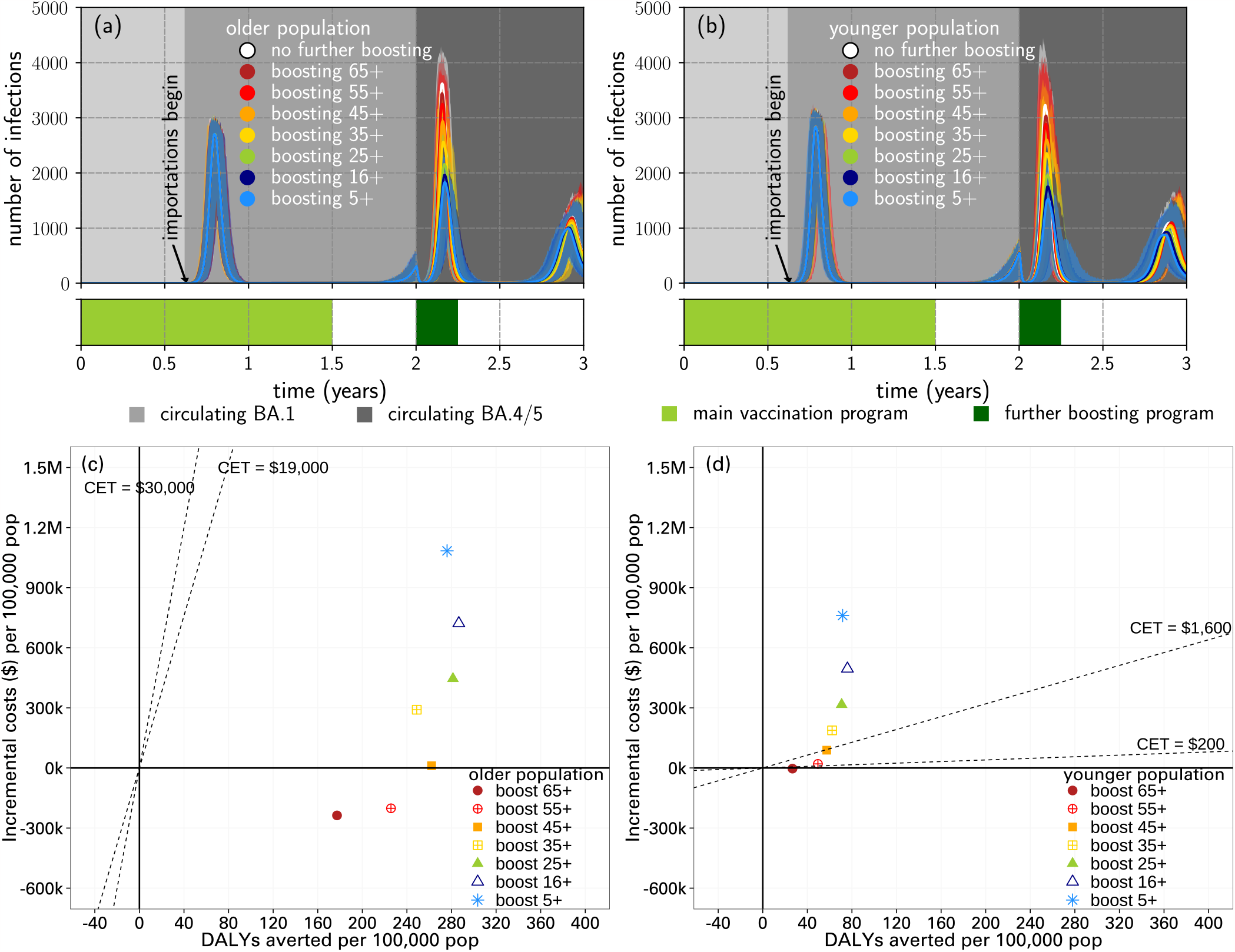
Outbreaks and cost-effectiveness analyses in the high transmission high vaccination coverage setting, for older and younger demographics, comparing the impact of lowering the age cut-off for high risk boosting. **(a)** epidemic waves in the older population; **(b)** epidemic waves in the younger population; **(c)** cost-effectiveness analysis in the older population; **d** cost-effectiveness analysis in the younger population. All scenarios here had an immune escape variant seeded at 2 years, with boosting at 2 years. The solid lines in (a) and (b) represent the pointwise median infections from 1000 simulations and the shaded regions represent the pointwise maximum and minimum infections. Boosting 65+ and 55+ is likely to be cost-effective or cost-saving.

### 3.2. Low-medium vaccination coverage scenarios

#### 3.2.1. Comparing primary and booster strategies

We consider the trade-off between new primary vaccination and high risk boosting strategies. We find that high risk boosting strategies perform better, reducing the size of epidemic peaks (Figure 6 (a), (b)).

**Figure 6:**
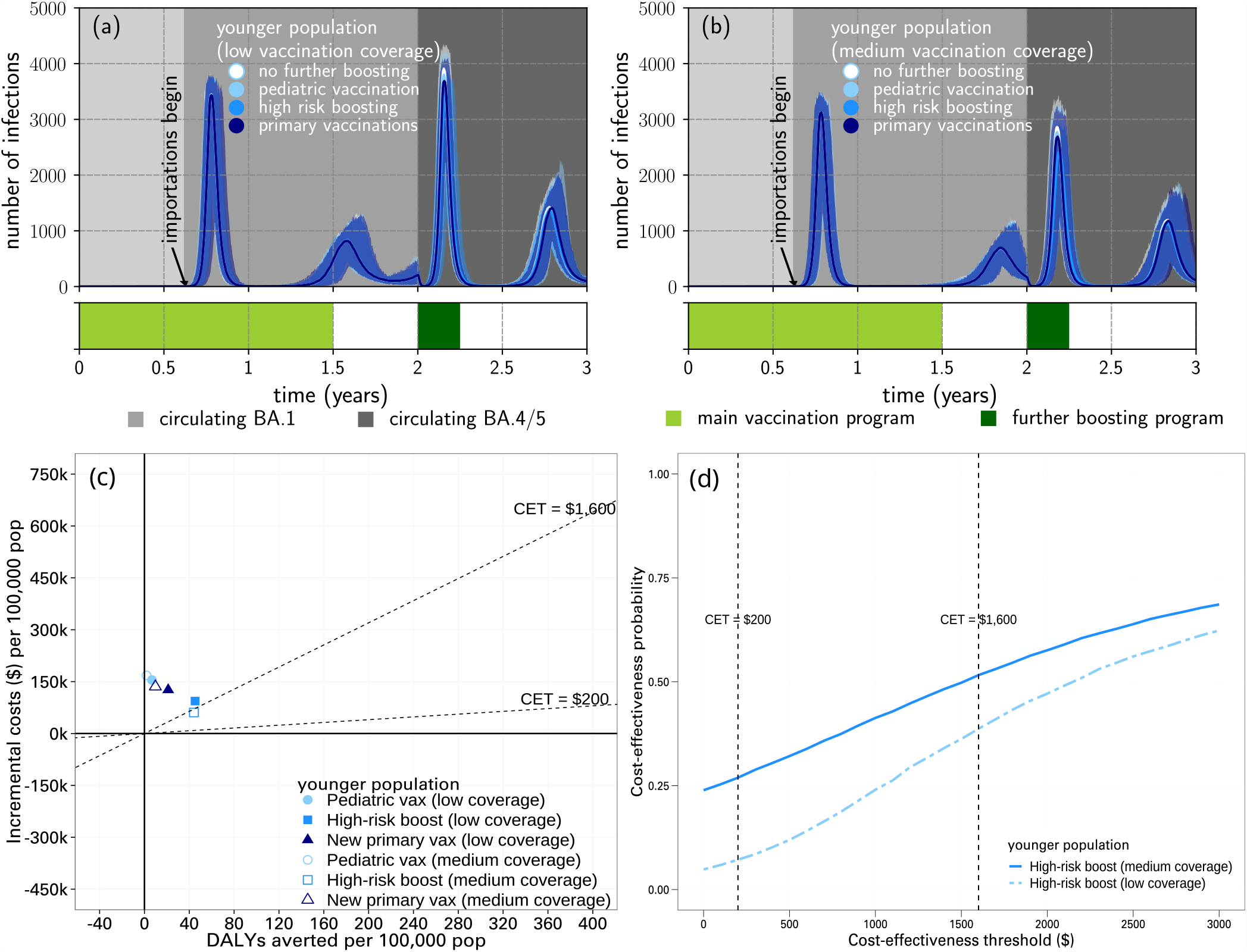
Outbreaks and cost-effectiveness analyses in the high transmission setting, with low and medium vaccination coverage for younger demographics. **(a)** epidemic curves for younger population with low vaccination coverage (initial coverage 20%); **(b)** for medium coverage (initial coverage 50%); **(c)** cost-effective analyses for low and medium vaccination coverage scenarios; **(d)** cost-effectiveness acceptability curves considering stochastic uncertainty and economic parameter uncertainty. All scenarios had an immune escape variant seeded at 2 years. These scenarios are run with vaccination strategies of pediatric vaccination (ages 5-15), high risk boosting (65+ first), and new primary vaccinations (allocated randomly among ages 5+) at 2 years. The solid lines in (a) and (b) represent the pointwise median infections from 1000 simulations and the shaded regions represent the pointwise maximum and minimum infections. Pediatric and new primary vaccinations are unlikely to be cost-effective.

New primary pediatric vaccination and new primary vaccination strategies are unlikely to be cost-effective (Figure 6(c)). High-risk boosting strategies may be cost-effective for younger (MIC) countries with high WTP thresholds or CETs (Figure 6(d)), but this depends on unit cost inputs, which are driven primarily by home-based care and vaccine program costs.

#### 3.2.2. Impact of bivalent boosting

We found a modest benefit in bivalent boosters over monovalent boosters on the dynamics of infections, with very minor differences in infection peak height (Figure E15 in the appendix) and also minor differences in the number of deaths between vaccine types(Table E6 in the appendix).

Given that the modelled benefits of bivalent boosters is minor compared with monovalent formulations, we anticipate that bivalent vaccines in this context will not be cost-effective unless willingness to pay is high. As such, we have not subjected these scenarios to formal cost-effective analyses.

## 4. Discussion

Our modelling approach allowed the exploration of COVID-19 booster dose impacts in diverse population settings. We assumed differing levels of prior vaccination delivery and infection experience, linked to income group level characteristics and vaccine program and healthcare costs. Given the age-dependency of severe disease risk, we concluded that elder-targeted strategies are most likely to be cost-effective (or even cost-saving) across a broad range of uncertainties. Notably, we consistently found pediatric programs (primary series or boosting) are not cost-effective. Absolute harms averted by vaccination are influenced by: age and risk profile of the population, prior immune landscape (infection exposure history, vaccination rollout), and timing of emergence of an immune escape variant in relation to booster delivery. Half-yearly ‘high risk’ booster programs are more expensive but may be cost effective in older, high income populations. However, this finding is much more uncertain in populations with younger demographics (representing upper- and lower-MIC), depending on the costs of community-based care and vaccine implementation.

A strength of our study was the modelling flexibility, enabling distinct configuration of the various interweaving elements. We used the immunity model to implement assumptions relevant to two exemplar Omicron variants, against which the effectiveness of ancestral and bivalent vaccines were explored. By introducing variants at different time points, we systematically evaluated the importance of epidemic timing in relation to plausible immune escape scenarios. Separate representation of clinical pathways and cost-effectiveness analysis further enabled adaptation to different settings. Health sector costs were exemplified by ‘averages’ based on regional data, with sensitivity analyses highlighting local drivers that will be influential for decision making depending on context.

As with all models, multiple simplifying assumptions were made for the purpose of tractability that do not necessarily reflect reality. We assumed circulation of a single dominant SARS-CoV-2 variant, to interrogate the ‘worst case’ scenario of a step change in vaccine effectiveness at a single point in time. The present state of multiple lineage co-circulation with variable immunological cross-reactivity and breadth likely lessens such impacts.^30^ We could have explored more optimistic values for bivalent vaccine effectiveness, which may lead to greater estimated benefits.^31^ Neither social restrictions nor antiviral agents were included, given that neither are presently being widely applied at the population level globally.

Long COVID was not included as a potential outcome of infection, as there remains limited quantitative data of this clinical burden. The anticipated costs of therapeutic pathways are yet to be determined pending identification of those which are most likely to be effective for various syndromic presentations.

A more significant limitation of our work in relation to health system costs is the assumption that the modelled adult-targeted vaccination coverage is achievable and can be costed across all country settings considered. In reality, most LMICs do not have adult immunisation programs in place, as highlighted by the COVID-19 pandemic.This deficiency is a global public health priority. We also do not consider indirect costs to society, such as productivity losses, associated with illness or death. A societal perspective would increase the cost-effectiveness of elder-targeted vaccination.

Our study considers a diversity of demographics and hybrid immunity histories, with lessons offered for countries that may fall between the “older” and ‘younger” exemplar groupings. We note that all our considered populations had some level of background vaccination (22%) by the end of the first Omicron wave; our work complements other studies^32;33^ that have considered the zero past vaccination setting with prior natural immunity in low and middle income countries.

As the world transitions towards COVID-19 endemicity, our study demonstrates the ongoing value of COVID-19 booster doses targeted towards older age groups at risk of severe outcomes in a range of demographics with different hybrid immunity histories. This approach is cost-effective or even cost-saving across multiple country settings. However, this assumes that adult immunisation programs are in place and do not impact on delivery of other services. Pediatric vaccination may be more readily implementable within existing health systems but was not cost-effective in any of the scenarios explored. These results were presented to the Advisory Committee on Immunization and Vaccines-related Implementation Research (IVIR-AC),^8^ with our work being subsequently cited as part of the WHO updated COVID-19 vaccination guidance for March 2023.^9^

SARS-CoV-2 reporting is being subsumed into broader respiratory pathogen surveillance systems. The WHO continues to encourage vigilance for identification of new variants with heightened transmissibility or pathogenic potential. A downward age shift in disease severity would require revision of current strategies. Our framework is sufficiently flexible to incorporate emerging evidence of virus and vaccine characteristics and can be configured to specific country settings. More broadly, it provides a template for the use of modelling to evaluate strategies for the control of any emerging or epidemic infectious disease.

## Data Availability

All data produced in the present study are available upon reasonable request to the authors. Code is available in the following GitHub repositories: https://github.com/goldingn/neuts2efficacy/ for the immunological model; https://github.com/spectrum-spark/covid_singlestrain_scenarios/tree/singlestrain-paper for the population transmission and clinical pathway models; and https://github.com/spectrum-spark/covid-CEA/ for the cost-effectiveness analysis.

https://github.com/spectrum-spark/covid_singlestrain_scenarios/tree/singlestrain-paper

https://github.com/spectrum-spark/covid-CEA/

## 5. Data sharing

Code is available in the following GitHub repositories: https://github.com/goldingn/neuts2efficacy/ for the immunological model; https://github.com/spectrum-spark/covid_singlestrain_scenarios/tree/singlestrain-paper for the population transmission and clinical pathway models; and https://github.com/spectrum-spark/covid-CEA/ for the cost-effectiveness analysis.

## 6. Acknowledgements

We would like to acknowledge contributions from Mackenzie Bourke, Alexandra Hogan, Nick Golding and Freya Shearer.

This work was supported by the World Health Organization.

In addition, we would like to acknowledge the support from the Australian Government Department of Foreign Affairs and Trade Indo-Pacific Centre for Health Security (Supporting Preparedness in the Asia-Pacific Region through Knowledge) and the Australian National Health and Medical Research Council (NHMRC) SPECTRUM CRE (GNT1170960).

ABH and IM are funded by NHMRC Investigator Grants (2021/GNT2009278 and 2022/GNT2016726 respectively). IM is also supported by NHMRC Principal Research Fellowship (GNT1155075).

## CRediT authorship contribution statement

**Thao P. Le:** Data curation, Formal Analysis, Investigation, Visualization, Writing – original draft, Writing – review & editing. **Eamon Conway:** Data curation, Methodology, Software, Formal Analysis, Writing – original draft, Writing – review & editing. **Edifofon Akpan:** Data curation, Formal Analysis, Investigation, Visualization, Writing – review & editing. **Isobel Abell:** Methodology, Writing – original draft, Writing – review & editing. **Patrick Abraham:** Data curation, Formal Analysis, Investigation, Visualization, Writing – original draft, Writing – review & editing. **Christopher M. Baker:** Conceptualization, Methodology, Supervision, Writing – original draft, Writing – review & editing. **Patricia T. Campbell:** Formal Analysis, Writing – original draft, Writing – review & editing. **Deborah Cromer:** Data curation, Methodology, Writing – original draft, Writing – review & editing. **Michael J. Lydeamore:** Methodology, Resources, Writing – original draft, Writing – review & editing. **Yasmine McDonough:** Validation, Writing – original draft, Writing – review & editing. **Ivo Mueller:** Conceptualization, Supervision, Writing – review & editing. **Gerard Ryan:** Data curation, Methodology, Writing – original draft, Writing – review & editing. **Camelia Walker:** Data curation, Methodology, Software, Writing – original draft, Writing – review & editing. **Yingying Wang:** Data curation, Formal Analysis, Investigation, Visualization, Writing – review & editing. **Natalie Carvalho:** Formal Analysis, Methodology, Supervision, Writing – review & editing. **Jodie McVernon:** Conceptualization, Funding acquisition, Supervision, Writing – original draft, Writing – review & editing.

