## Appendix for "Cost-effective boosting allocations in the post-Omicron era of COVID-19 management"

[1]School of Mathematics and Statistics, The University of Melbourne, Melbourne, Victoria, Australia
[2]Melbourne Centre for Data Science, The University of Melbourne, Melbourne, Victoria, Australia
[3]Centre of Excellence for Biosecurity Risk Analysis, The University of Melbourne, Melbourne, Victoria, Australia
[4]Population Health & Immunity Division, Walter and Eliza Hall Institute of Medical Research, Parkville, Victoria, Australia
[5]Melbourne School of Population and Global Health, The University of Melbourne, Victoria, Australia
[6]Department of Infectious Diseases, The University of Melbourne at the Peter Doherty Institute for Infection and Immunity, , Melbourne, Victoria, Australia
[7]Kirby Institute, University of New South Wales, Sydney, New South Wales, Australia
[8]Department of Econometrics and Business Statistics, Monash University, Melbourne, Victoria, Australia
[9]Department of Medical Biology, University of Melbourne, Melbourne, Victoria, Australia
[10]Telethon Kids Institute, Nedlands, Western Australia, Australia
[11]Victorian Infectious Diseases Reference Laboratory Epidemiology Unit at the Peter Doherty Institute for Infection and Immunity, The Royal Melbourne Hospital, Melbourne, Victoria, Australia
[‡]These authors should be considered as joint senior author.

E.5 Low-medium vaccination coverage: impact of bivalent boosting - extended results . . 58

References 61

### A Immunological model

Within both the transmission model (Section B) and the clinical outcomes model (Section C) we directly modelling each individual's neutralising antibody titre. Khoury and colleagues [1, 2] developed a model of correlates of protection relating an individual's neutralising antibody titre to their protection against sympotomatic and severe disease outcomes. Here we use Golding and colleagues' [3] implementation of that model, which used the relationship between neutralising antibodies efficacy of protection along with data on efficacy and time since vaccination to extend the work to estimate the relationship between neutralising antibody titres and protection against all outcomes of interest: infection, symptomatic disease, onward transmission given breakthrough infection, hospitalisation and death. Within our model each individual is assigned their own neutralising antibody titre, which results in inter-individual variation leading to varying distributions of protection throughout the community, thus impacting the resulting dynamics. In the Australian context, at the time Delta and Omicron outbreaks occurred almost all immunity was vaccine-derived, therefore the model is initialised by considering neutralisating antibody titre from vaccination alone.

#### A.1 Neutralising antibody titres

An individual's neutralising antibody titre can be increased to different degrees by different exposures to the virus. The processes that we consider are: (i) the first, second or booster dose of a vaccine or (ii) infection occurring in either a unvaccinated or vaccinated individual. Note that for simplicity we assume that infection prior to or following vaccination results in the same titre of neutralising antibody. Immune responses are stratified by the type of vaccine product, AstraZeneca (AZ) or mRNA vaccine (Pfizer or Moderna), that the individual has received based on supply and distribution data.

At the time of an exposure, we sample the neutralising antibody titre acquired, $a_i^0$ , from

$$\log_{10}(a_i^0) \sim \mathcal{N}(\mu_j^x, \sigma^2), \tag{1}$$

where $a_i^0$ is the neutralising antibody titre of individual $i$ after exposure, $\mu_j^x$ is the mean neutralising antibody titre against strain $x$ in the population after exposure process $j$ and $\sigma^2$ is the variance of neutralising antibodies across the population.

The mean neutralising antibody titre, $\mu_j^x$, is set using logical rules based upon the infection and vaccination history of the individual. As in [1], in our work it is assumed that an unvaccinated individual will have an average neutralising antibody titre of 0.0 on the $\log_{10}$-scale after exposure. This is our baseline measurement and is used to calibrate across multiple neutralising antibody studies. The mean vaccine induced antibody response for an individuals with no prior exposure to COVID-19 is estimated such that,

$$\mu_j^x = \mu_j^0 + \log_{10}(f_x), \tag{2}$$

where $\mu_j^0$ is the mean level of neutralising antibody titre for vaccine $j$ against a base strain of COVID-19 (for us this is Delta) and $f(x)$ is the fold change in neutralising antibody titre between the base strain and strain $x$. To account for the effect of exposure to COVID-19 prior or post vaccination, we use an altered form of Eqn. 2. For brevity, we have used Table A1 to list the equations used to obtain $\mu_j^x$ with infection.[1] The value of the parameters used in our subsequent simulations are listed in Table B1.

**Table A1:** The relationships assumed within our immunological model for neutralising antibody titre for individuals that have been exposed to COVID-19. Here we have used the extended subscript with an E to represent prior or current exposure to the circulating strain of COVID-19.

| Process | Average titre formula |
|---|---|
| Unvaccinated ($U \cap E$) | $\mu_{U \cap E}^x = \mu_U^0$ |
| AZ dose 1 ($AZ1 \cap E$) | $\mu_{AZ1 \cap E}^x = \mu_{P2}^0$ |
| AZ dose 2 ($AZ2 \cap E$) | $\mu_{AZ2 \cap E}^x = \mu_B^0$ |
| Pfizer dose 1 ($P1 \cap E$) | $\mu_{P1 \cap E}^x = \mu_{P2}^0$ |
| Pfizer dose 2 ($P2 \cap E$) | $\mu_{P2 \cap E}^x = \mu_B^0$ |
| mRNA booster ($B \cap E$) | $\mu_{B \cap E}^x = \mu_B^0$ |

It is assumed that an individual's titre of neutralising antibodies will decay after boosting. This decay is assumed to be exponential, therefore,

$$\log_{10}(a_i) = \log_{10}(a_i^0) - \frac{k_a}{\log(10.0)} t, \quad (3)$$

where $a_i$ is the time dependent neutralising antibody titre of individual $i$, $k_a$ is the decay rate of neutralising antibodies and $t$ is the time from the last boosting process (to limit the computational cost of constantly converting neutralising antibodies, all equations are expressed in terms of $\log_{10}(a_i)$ in our work).

To convert the neutralising antibody titre of an individual to their protection against any disease outcome, $\rho_\alpha$, we use

$$\rho_\alpha = \frac{1}{1 + \exp(-k(\log_{10}(\alpha_i) - c_\alpha))}, \quad (4)$$

where $k$ governs the steepness of the logistic curve (logistic growth rate), and $c_\alpha$ defines the midpoint of the logistic function for disease outcome $\alpha$.

The immunological model interacts with the transmission model by altering the probability that an individual develops symptoms, $q_i$, their rate of onward transmission given breakthrough infection, $\tau_i$, and the contact's level of susceptibility, $\xi_j$.

The susceptibility of contact $j$ is,

$$\xi_j = (1 - \rho_\xi)\xi_i^0, \quad (5)$$

[1]These formulae are updated as information continues to evolve.

where $\rho_\xi$ is the protection against infection and $\xi_i^0$ is the susceptibility of the $i$th individual if they were completely COVID naive. The probability that individual $i$ develops symptoms is governed by,

$$q_i = \frac{1-\rho_q}{1-\rho_\xi} q_i^0, \tag{6}$$

where $\rho_q$ is the protection against symptomatic infection and $q_i^0$ is the probability of symptomatic infection for individual $i$ if they were completely COVID naive (zero neutralising antibody titre).

To model the onward transmission rate more care must be taken. It is assumed in our model that asymptomatic individuals are 50% less likely to infect their contacts when compared to their symptomatic counterpart. However, this reduction in transmission due to asymptomatic infections is not accounted for in the clinical trial data used to calibrate the protection against onward transmission. To avoid double counting the effect of the neutralising antibodies we alter the functional form for the rate of onward transmission to,

$$\tau_i = \frac{s(1-\rho_\tau)(1+q_i^0)}{1+q_i} \beta_i, \tag{7}$$

where $s$ is either 0.5 or 1 depending upon whether the individual is asymptomatic or symptomatic respectively, $\rho_\tau$ is the protection against onward transmission and $\beta_i$ is the baseline (zero neutralising antibody titre) infectiousness of the infector. Note that $\beta_i$ depends upon the age of the individual and the expected transmission potential of the population.

To reduce the computational cost of updating the immunological component of the transmission model for each individual at every timestep, we only solve the immunological component of the IBM when we require the protection against an outcome of interest. This is done by storing the time of last boost of neutralising antibodies and the titre that the individual was boosted to. When required, we update the individual's neutralising antibody titre to the current timestep using this stored information.

#### A.2 Clinical outcomes

The clinical outcomes model uses the transmission model as an intermediary between the immunological response of each infected individual and their corresponding clinical outcome. This is done by outputting each infected individual's neutralising antibody titre at the point of exposure, a symptom indicator and their time of symptom onset for use within the clinical pathways model.

The immunological model determines the probability of hospitalisation, ICU requirement and death based on observed relationships between neutralising antibody titres and clinical endpoint outcomes from efficacy studies. For a symptomatic individual $i$, the probability of hospitalisation is given by

$$p_{H|I}^i = \frac{OR(p_{H|E}^0, \rho_h(\alpha_i))}{q_i}, \tag{8}$$

where $p_{H|E}^i$ is the baseline probability of hospitalisation given infection, $\rho_h(\alpha_i)$ is the protection against hospitalisation, $\alpha_i$ is individual $i$'s neutralising antibody titre at the point of exposure, and

$$OR(p, r) = \frac{\frac{rp}{p-1}}{1 + \frac{rp}{1-p}}, \tag{9}$$

is the function that uses odds ratio $r$ and baseline probability $p$ to compute an adjusted probability.

If individual $i$ is hospitalised, the probabilities governing which hospital pathway is chosen are altered such that,

$$p^i_{ICU|H} = \frac{OR(p^0_{ICU|E}, \rho_h(\alpha_i))}{p^i_{H|I} q_i}, \tag{10}$$

and,

$$p^i_{H_D|ICU^c} = \frac{OR(p^0_{H_D|E}, \rho_D(\alpha_i))}{(1 - p^i_{ICU|H}) p^i_{H|I} q_i}, \tag{11}$$

where $p^0_{ICU|E}$ is the baseline probability of requiring the ICU given infection, $p^0_{H_D|E}$ is the probability of death on ward (without visiting ICU) given infection and $\rho_D(\alpha_i)$ is the protection against death given infection.

If individual $i$ is in the ICU, then their probabilities of death in the ICU, $p^i_{ICU_D|ICU}$, and death on the ward given they left ICU without dying, $p^i_{W_D|ICU^c_D}$, are altered such that,

$$p^i_{ICU_D|ICU} = \frac{OR(p^0_{ICU_D|E}, \rho_D(\alpha_i))}{p^i_{ICU|H} p^i_{H|I} q_i}, \tag{12}$$

and,

$$p^i_{W_D|ICU^c_D} = \frac{OR(p^0_{W_D|E}, \rho_D(\alpha_i))}{(1 - p^i_{ICU_D|ICU}) p^i_{ICU|H} p^i_{H|I} q_i}, \tag{13}$$

where $p^0_{ICU_D|E}$ is the baseline probability of dying in the ICU and $p^0_{W_D|E}$ is the baseline probability of dying in the ward after returning from the ICU. Note that we assume no difference between the protection from hospitalisation given infection and the protection from ICU given infection here.

To determine all parameters in Eqn. (3) and Eqn. (4), we use a re-implementation of [1] and [2] in a Bayesian framework [3]. This allows us to calibrate the level of protection, which is analogous to vaccine efficacy for individuals with no exposure to COVID-19, to observed clinical data. The model fit in Golding (2022) [3] takes in a range of data relating neutralising antibody levels to efficacy, and estimates of vaccine efficacies from a range of studies to estimate efficacy over time against the Delta variant. To estimate the efficacies against the Omicron variant, Golding and colleagues estimate an 'escape' parameter for the Omicron variant relative to the Delta variant. This was done by using the relative rates of infection in Danish households between Omicron and Delta to estimate the relative $R_0$ between the variants, and early evidence of vaccine efficacies against Omicron from the UK to understand the level of vaccine escape. This was then combined with the information fit on the Delta variant to model waning over time for both the Delta and Omicron variants.

#### B Population transmission model

To model the transmission of COVID-19 throughout the population of interest we have extended a previous individual-based model [4] to account for loss of protection over time since vaccination. Waning of immunity is accounted for by modelling the boosting and decay of each individual's neutralising antibody titre. This approach overcomes limitations of the typical SIRS model, where waning is modelled as an exponential distribution (or more generally as a phase type distribution), and enables us to investigate the possible impacts of variants of concern using the expected fold change in neutralising antibody titre.

Each individual within the simulation is constructed using a known age, their corresponding age bracket within the contact matrix, a list of their vaccination history and the decay rate of their neutralising antibodies. Note that each individual's entire vaccination history must be known *a priori* and is an input into the transmission model. This allows us to incorporate known vaccination information and can quickly switch out different vaccination scenarios.

At the start of the simulation, each individual is assumed to be susceptible and to have zero neutralising antibodies. Once an individual is constructed, they are dynamically stored within a vector. As the simulation progresses, the individual is vaccinated according to their inputted vaccination history, and can become sick and spread diease, as follows:

The spread of infection is modelled by directly simulating contact between infectious and susceptible individuals. For an infectious individual $i$ in age bracket $n$, we sample the number of contacts that they make in age bracket $m$ from a negative binomial (NB) distribution, such that,

$$C_m \sim \text{NB}\left(r\delta t, \frac{\Lambda_{n,m}}{r + \Lambda_{n,m}}\right), \tag{14}$$

where $C_m$ is the number of contacts made this timestep by the individual in age bracket $m$, $r$ is the overdispersion parameter (in terms of contacts per day), $\delta_t$ is the size of the current time step (in units of days), and $\Lambda_{n,m}$ is the mean number of daily contacts between age bracket $n$ and $m$. Note that we have used the definition of the negative binomial distribution where $\text{NB}(r, p)$ corresponds to probability density function $f(k) = \binom{k+r-1}{r} p^k (1-p)^r$. We then sample $C_m$ contacts uniformly from individuals within the $m$th age bracket. If contact $j$ is susceptible we determine if infection occurred using,

$$I \sim \text{Bernoulli}(\tau_i \xi_j), \tag{15}$$

where $I$ is an indicator variable for successful infection, $\tau_i$ is the infectiousness of the infectious individual $i$, and $\xi_j$ is the susceptibility of the contact $j$. We note that $\tau_i$ depends upon the transmission potential of the population of interest [5], and both $\tau_i$ and $\xi_j$ depend upon the underlying immunological model (Section A) and the heterogeneous characteristics of the individual of interest (for example, $\tau_i$ will be altered depending on the symptom status of the infectious individual and both $\tau_i$ and $\xi_j$ will depend upon age). This process of generating contacts and the resulting infections is repeated over all age brackets.

At the point of infection, whether an individual will be asymptomatic or symptomatic is sampled from

$$Q \sim \text{Bernoulli}(q_i), \tag{16}$$

where $Q$ is an indicator variable for symptomatic infection and $q_i$ is the probability that individual $i$ will be symptomatic, which depends on the age and neutralising antibody titre of individual $i$. We also sample the time that the newly exposed individual becomes infectious, the time for the onset of symptoms and the time that the individual will recover.

When the infectious individual recovers, we store the individual's age, time of symptom onset, neutralising antibody titre at exposure, symptom indicator ($Q$), the number of individuals they infected, their vaccine status (what was the latest vaccine they received) at exposure, the time they were isolated from the community, and the number of times that they have been infected. This generates a line list of infections that is used to model clinical outcomes (Section C).

#### B.1 Scenario and input parameters

In our scenarios, we vary population type, immune escape, transmission potential (TP), vaccine coverage and boosting strategy. The broad timeline of the scenarios/simulations is shown in Figure B1.

Each scenario was run 1000 times, as we found that results comfortably converged by that point.

##### B.1.1 Population types

We considered "older" and "younger" populations, separating them based on *old-age dependency ratio*, or $OADR$, which is defined as

$$OADR = \frac{\text{population aged 65 and over}}{\text{population aged 20–64}} \times 100.$$

So, our "older" populations have an $OADR \geq 15$ , while our "younger" populations have an $OADR \leq 12$. In practice, this means:

- **"Older" population** is representative of high-income countries (HIC) in the WPR. The "older" population distribution is averaged from China, Hong Kong SAR, Macao SAR, Japan, Republic of Korea, Singapore, Australia, New Zealand, New Caledonia, Guam, and French Polynesia;
- **"Younger" population** is representative of mostly lower-middle income and some upper-middle income countries (MIC) in the WPR. The "younger" population distribution is averaged from Mongolia, Brunei Darussalam, Cambodia, Lao People's Democratic Republic, Philippines, Fiji, Papua New Guinea, Solomon Islands, Vanuatu, Kiribati, Micronesia (Fed. States of), Samoa, and Tonga.

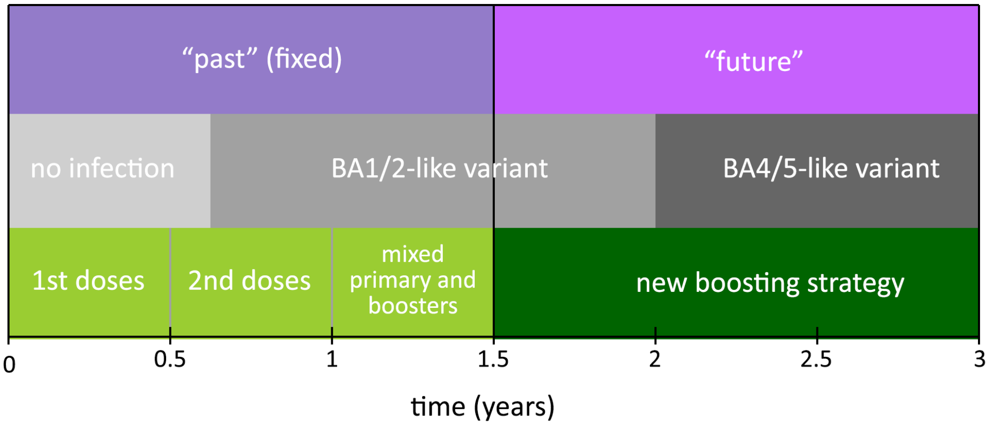

**Figure B1:** Broad timeline of the vaccination schedule and circulating variant. We consider the first 1.5 years to be the "past", and we are focused on multiple different strategies and scenarios in the "future" between 1.5—3 years. Note that we vary the precise timing of the new boosting strategy, and the time the BA4/5-like variant emerges in different scenarios.

We acquired 2021 population data from the United Nations population database [6].

The older and younger population distributions are displayed in Figures B2(a) and (b) respectively. The older population demographic has more of the population aged around 20–64 years, while the younger population demographic skews downward and peaks at the 5–11 age group. For the simulations, we generated 100,000-person populations based on these distributions.

The older and younger populations in our model also have a different contact matrix describing social mixing (and thus infection spread) in the model (Figure B3).

To derive some exemplar contact matrices, we used aggregated contact matrices from "older" ($OADR \geq 15$) and "younger" ($OADR \leq 12$) countries that could be found on http://www.socialcontactdata.org/ [8, 9, 10, 11, 12, 13, 14, 15, 16, 17, 18]: for "older" countries we included Belgium, Finland, France, Germany, Hong Kong, Italy, Luxembourg, Poland; and "younger" included Vietnam and Zimbabwe. These countries fell into the appropriate *OADR* values and had contact matrix values for all the age-groups we required.

##### B.1.2 Transmission potential

Transmission potential (TP) reflects different populations' intrinsic transmission, which is dependent on a variety of factors such as demographics, weather/climate, housing, population density etc. In general, populations with high TP have a high past attack rate and populations with low TP have a low past attack rate.

We consider scenarios with:

- **High TP** (with mean attack rates around 80%–100% in the first 1.5 years depending on vaccination coverage); and

- **Low TP** (with mean attack rates around 15%–45% in the first 1.5 years depending on vaccination coverage).

###### B.1.3 Variants and immune escape

All scenarios start with a BA1/2-like variant, with an introduced immune escape variant (BA4/5-like) at a later time. When an immune escape variant emerges, all prior neutralising antibodies are suppressed relative to the new variant, representing the immune escape. The new circulating disease also has a higher transmission potential than the prior variant.

We consider immune escape variant introduction at either:

- **1.5 years**;
- **2.0 years**; or
- **2.5 years**

after the start of the main vaccination program.

###### B.1.4 Vaccination

**Vaccination coverage**

We consider three initial vaccination coverage levels, which are also depicted in Figure B2(c), (d) and (e) (for the younger population):

- **High coverage**: 80% primary vaccine coverage after the first year (88% primary vaccine coverage by 1.5 years) in both "older" and "younger" populations;
- **Medium coverage**: 50% primary vaccine coverage (55% primary vaccine coverage by 1.5 years) in the "younger" population only; and
- **Low coverage**: 20% primary vaccine coverage (22% primary vaccine coverage by 1.5 years) in the "younger" population only.

Note that low vaccine coverage applies to a subset of "younger" populations where this scenario is relevant (such as Papua New Guinea and the Solomon Islands) — this is because countries with "older" populations tend to have high vaccination coverages, and exemplar low-vaccination coverage populations have younger population distributions.

**Vaccination schedule in 0–1.5 years**

The broad schedule in the first 1.5 years of our simulation is shown in Figure B1. Either 20%, 50%, or 80% total vaccination coverage (complete primary doses) is achieved by the end of the first year, with first doses given out in the first 6 months, and second doses given out in the second 6 months.

During time 1–1.5 years, 80% of already vaccinated individuals are given boosters, while the remaining number of doses is given as primary course vaccinations.

The vaccine allocation broadly follows WHO guidelines, which recommends prioritising the vaccination of older and higher-risk groups. At the lower vaccination rate of 20%, we first allocate doses such that 80% of the 65+ age group are fully vaccinated by the end of the first two stages. At the higher vaccination rates of 50% and 80%, we allocate initial doses such that 95% of the 65+ age group are fully vaccinated by the end of the first two stages. The remaining available doses in the first two stages are then equally (proportionately) allocated to the 5–64 age groups.

###### Vaccination schedule in 1.5–3 years

Exploring different vaccination timings and targeting in the 1.5–3 year period is the main investigation of this paper, so we defer closer examination of these schedules to sections B.1.5, B.1.6, B.1.7.

###### Vaccine type

We assume that all primary doses are monovalent ChAdOx1 nCoV-19 (AstraZeneca) and all booster doses are monovalent BNT162b2 (Pfizer/BioNTech), unless otherwise noted (i.e. in the section comparing bivalent vaccination with monovalent vaccination).

##### B.1.5 High coverage boosting strategies

For the high coverage vaccination scenarios, we primarily consider three boosting strategies:

- **Pediatric boosting**;
- **High risk boosting**; and
- **Random boosting**.

We fix the number of vaccine doses (11,000) in these scenarios, so that we can focus on the impact of vaccine allocation. 11,000 doses is enough doses to boost approximately 80% of the 65+ age group in the older population (55+ age group in the younger population), or to boost approximately 80% of individuals aged 5–15 in the older population. Figure B4 shows the vaccine allocation across the populations for each boosting strategy.

Alongside the age-based boosting strategies, we also consider how timing affects the results. We consider two options, with boosters starting at either a fixed point (e.g. at 2 years) or delivered routinely (e.g. every 6 months). Each instance of a booster program takes approximately 3 months to administer.

##### B.1.6 Low coverage vaccination and boosting strategies

Here we consider scenarios for younger populations with low or medium vaccination coverage. We have three different vaccination/boosting strategies targeting different groups:

- New pediatric primary vaccination,
- High risk boosting (older first),
- New general (random) primary vaccinations.

In the low and medium vaccination coverage scenarios, there is typically *insufficient* numbers of vaccinated individuals to be given a booster dose (all 11,000 of them). Because there is a large number of individuals still unvaccinated, we consider two new primary vaccination programs, one to children, and one to the general population. There are still 11,000 total doses, meaning that 5,500 individuals will be fully vaccinated in these new vaccination programs.

##### B.1.7 Boosting strategies for the age-cutoff-investigation

In the previous strategies, we typically defined "high risk boosting" as boosting to the 65+ age groups within the older population, and to the 55+ age groups within the younger population. Here we consider what would happen if we systematically reduce this age threshold (i.e., increasing booster eligibility to increasingly younger age groups), to test the limits of cost effectiveness of coverage. In order to have high booster coverage, we need high primary vaccination coverage. As such, we only consider the high-coverage (80%) scenarios, in both older and younger population settings.

We considered seven boosting strategies:

- **65+ years boosting**,
- **55+ years boosting**,
- **45+ years boosting**,
- **35+ years boosting**,
- **25+ years boosting**,
- **16+ years boosting**,
- **5+ boosting**.

There is a fixed percentage of booster uptake: 80% of eligible individuals (who have had a primary series vaccination) receive a booster. This means that the number of booster doses given out increases as the eligibility age decreases. Since the younger population and older population have different age distributions, the number of doses changing per 'step' will be different between the two.

Note that the 65+ years scenario that we test here is not the same as the prior "further boosting high risk" scenarios in the rest of the report, where a fixed number of doses (11,000) were administered. Here, we administer sufficient doses to achieve 80% booster uptake, which is not exactly equal to 11,000.

##### B.1.8 Bivalent boosting

Bivalent boosting is expected to have a greater impact in populations with low levels of vaccination [19]. Hence we only explored low- and medium-coverage vaccination settings in younger populations. Khoury et al [19] found that bivalent vaccines, on average, produced 1.61-fold higher titers than monovalent vaccines. We implement bivalent boosters within the model by using this average multiplier (1.61) on top of the Pfizer booster neutralisation values for any boosters given out during the 1.5–3 year stage as part of high-risk boosting.

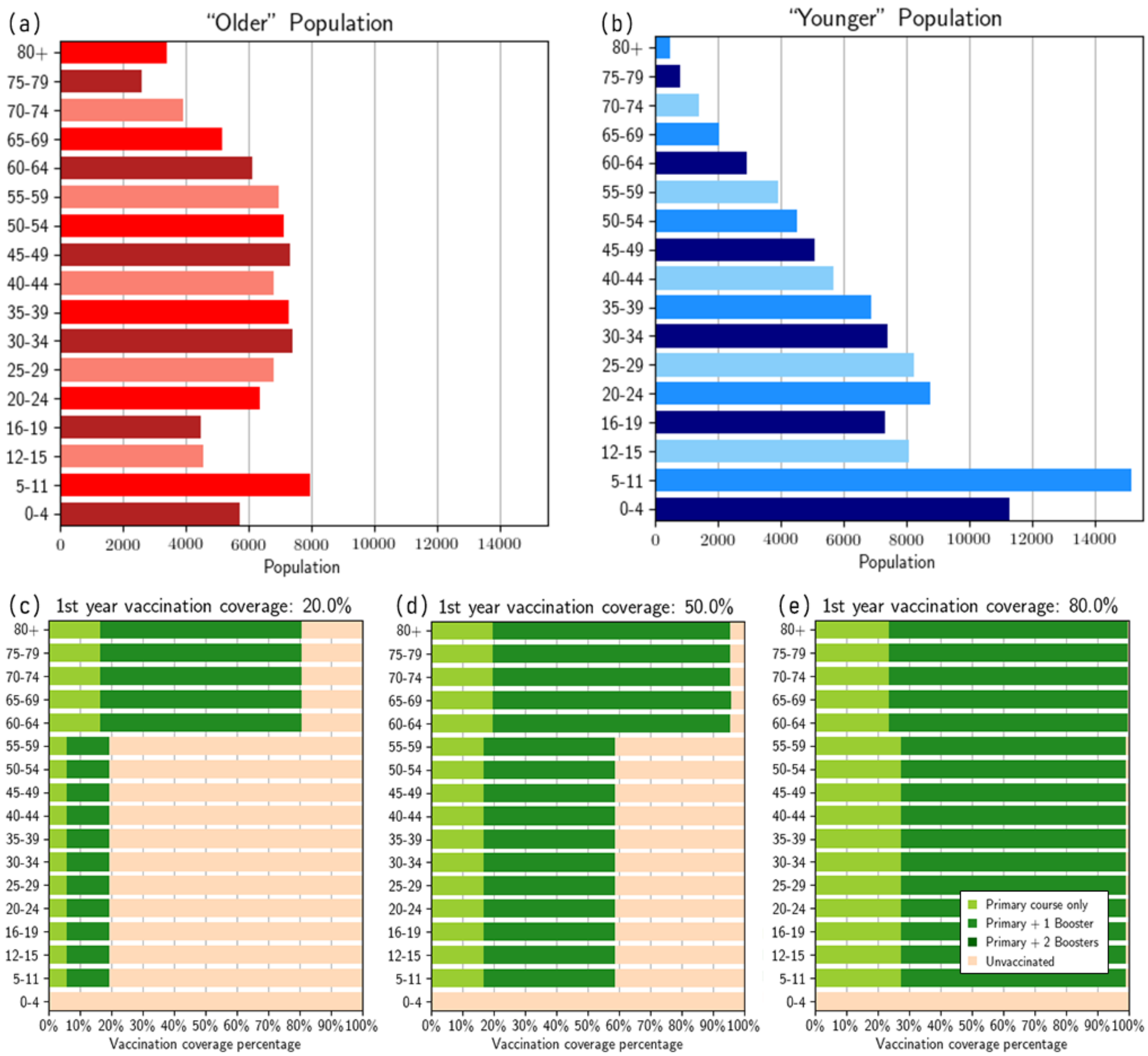

**Figure B2:** Population demographics and initial vaccination coverages. (a) "older" population distribution; (b) "younger" population distribution; (c) low initial vaccination coverage* after 1.5 years, (d) medium initial vaccination coverage* after 1.5 years, (e) high initial vaccination coverage* after 1.5 years. *Note: proportions are for the younger population, but it is very similar for the older population. Adapted from Figure 3 from [7].

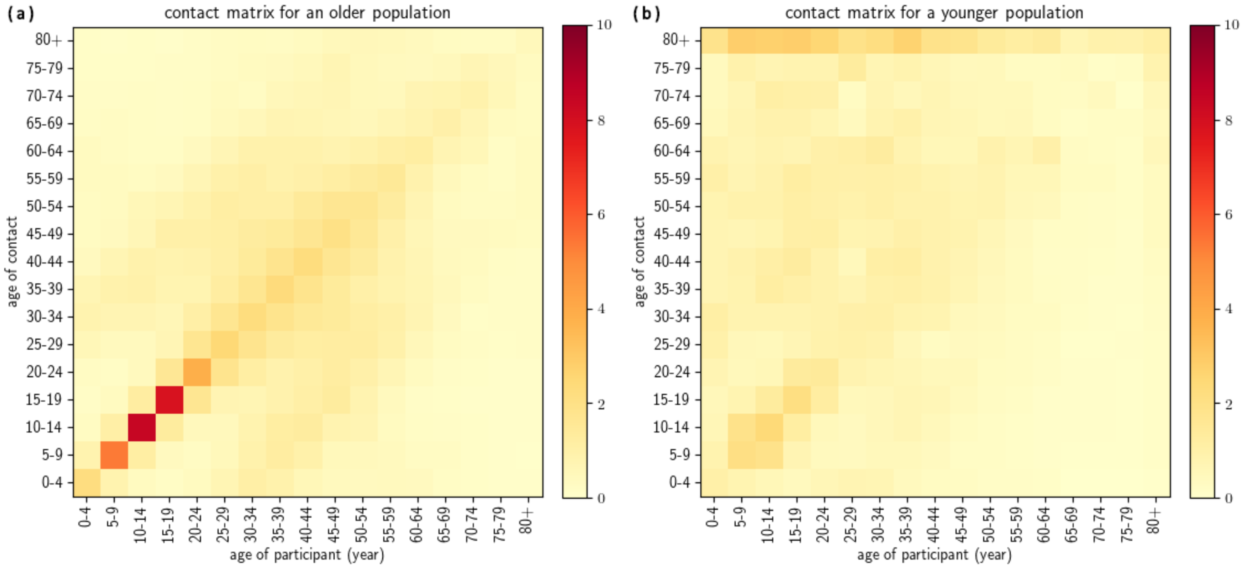

**Figure B3:** Contact matrices used for: (a) “older” population, (b) “younger” population. Adapted from Figure 8 from [7].

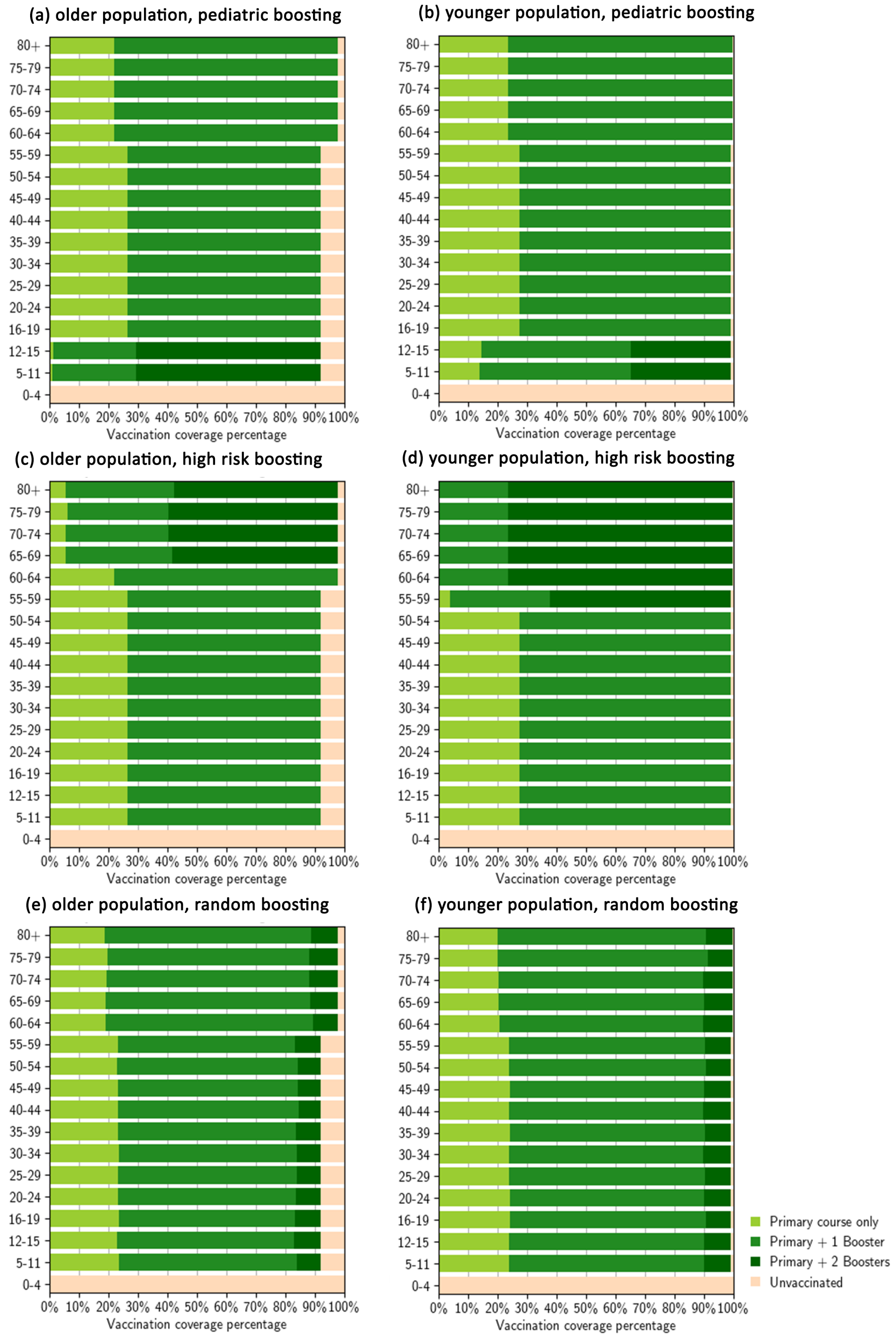

**Figure B4:** Vaccination coverage distribution after different high-coverage boosting allocations in the older and younger populations.

**Table B1:** Estimated parameter values from the immunological model used in the simulations. Note that the age brackets for probabilities of symptomatic infection (prob_symptoms), relative infectiousness once infectious (relative_infectiousness), and susceptibility to becoming infected upon contact with an infected individual correspond to [0, 5, 10, 15, 20, 25, 30, 35,40, 45, 50, 55, 60, 65, 70, 75, 80]. "$c_{50}$" corresponds to the midpoint of the logistic function for a particular disease outcome (hospitalisation, death etc...). Source: [3, 20].

| Parameter: description | Value |
|---|---|
| $c_h$: $c_{50}$ of hospitalisation | -1.2161786725147814 |
| $c_d$: $c_{50}$ of death | -1.1753151405677293 |
| $c_\xi$: $c_{50}$ of acquisition | -0.47195962651907175 |
| $c_\tau$: $c_{50}$ of transmission | 0.02953683449343693 |
| $c_q$: $c_{50}$ of symptoms | -0.6442020773907903 |
| $\mu^0_{\text{AZ1}}$: $\log_{10}$ of the mean neutralising antibody titre after the first dose of AstraZeneca (no infection) | -0.5299522095755013 |
| $\mu^0_{\text{AZ2}}$: $\log_{10}$ of the mean neutralising antibody titre after the second dose of AstraZeneca (no infection) | -0.12031713312180076 |
| $\mu^0_{\text{P1}}$: $\log_{10}$ of the mean neutralising antibody titre after the first dose of Pfizer (no infection) | -0.23154132545543396 |
| $\mu^0_{\text{P2}}$: $\log_{10}$ of the mean neutralising antibody titre after the second dose of Pfizer (no infection); also the mean titre after one AZ dose and one infection | 0.1540166902000597 |
| $\mu^0_{\text{B}}$: $\log_{10}$ of the mean neutralising antibody titre after the first mRNA booster dose (without infection); also the mean titre after two AZ doses and one infection | 0.3225538899068383 |
| log10_mean_neut_bivalent_booster | 0.52937976593 |
| $\mu^0_U$: $\log_{10}$ of the mean neutralising antibody titre after infection whilst unvaccinated | 0.0 |
| $\log(k)$: governs the logistic curve steepness relating antibodies to protection against disease outcome | 1.686059432639791 |
| $k_a$: decay rate of neutralising antibodies | 0.008235096361537353 |
| $\log_{10}(f_{\text{Delta}})$: $\log_{10}$ of the fold change in neutralising antibody titre between Delta and the baseline, Delta | 0.0 |
| $\log_{10}(f_{\text{Omicron}})$: $\log_{10}$ of the fold change in neutralising antibody titre between Delta and Omicron (BA1-like) | -0.6923808174384031 |
| $\sigma$: standard deviation of the $\log_{10}$ of neutralising antibodies across the population | 0.4647092 |
| probability of symptoms across age groups | [0.29, 0.29, 0.21, 0.21, 0.27, 0.27, 0.33, 0.33, 0.4, 0.4, 0.49, 0.49, 0.63, 0.63, 0.69, 0.69, 0.69] |
| relative infectiousness across age groups | [ 0.79915, 0.687631, 0.675465, 0.756167, 0.918134, 0.965157, 0.947093,0.932175, 0.933528, 0.939668, 0.953521, 0.981757, 1.0, 0.998492, 0.989752, 0.973762, 0.943804 ] |
| susceptibility across age groups | [ 0.301016, 0.367215, 0.432601, 0.527461, 0.764291, 0.92387, 0.982666, 0.974271, 0.93188, 0.914635, 0.928871, 0.962179, 1.0, 0.972011, 0.882005, 0.823837, 0.802185 ] |
| $\log_{10}(f_{\text{Omicron-escape}})$: $\log_{10}$ of the fold change in neutralising antibody titre between the original Omicron variant (BA1-like) and the BA4/BA5-like immune escape variant | -1.18049745646 |
| $R_0$ ratio between the original Omicron variant (BA1-like) and the BA4/BA5-like immune escape variant | 1.3 |

### C Clinical pathways

The clinical outcomes model is based on the clinical model from [21] and is an extension of the work done in [4] and [22]. This model extends on previous work by restructuring the existing model as a continuous-time, stochastic, agent-based model, where the neutralising antibody titre and age of the individual determines their transition probabilities. Furthermore, an emergency department (ED) compartment has been added as [23] noted that limited ED consult capacity can cause a bottleneck that prevents admission to hospital. The full compartmental structure of the clinical outcomes model is depicted in Figure C1.

All parameters governing the pathway each individual takes through the health system are altered depending upon their individual neutralising antibody titre. The evaluation of these transition probabilities are explained in full in the immunological model (Section A).

For each symptomatic individual in the line-list outputted from the transmission model (Section B), we determine if they are hospitalised by sampling,

$$H \sim \text{Bernoulli}(p^i_{H|I}), \tag{17}$$

where $H$ is an indicator variable for hospitalisation and $p^i_{H|I}$ is the probability that individual $i$ is hospitalised given symptomatic infection. If individual requires hospitalisation they will present to the ED, where they may not be seen due to capacity limitations. ED consult capacity is modelled by admitting only the first $C_{ED}$ presentations to ED each day. If individual $i$ is not seen, they will present again to the ED with probability $1 - p_{L|ED}$ after $\tau_{L|ED}$ days sampled from,

$$\tau_{L|ED} \sim \text{Gamma}(\kappa_{L|ED}, \theta_{L|ED}), \tag{18}$$

where $p_{L|ED}$ is the probability that an individual does not present again to the ED and $\kappa_{L|ED}$ and $\theta_{L|ED}$ are the shape and rate parameters of the gamma distribution respectively. For individuals that do not return to ED and are therefore not admitted to hospital, their age, neutralisation titre upon exposure and number of presentations to ED are recorded such that these can be used to understand possible excess mortality due to ED capacity limits. If individual $i$ is admitted to hospital we determine what hospital pathway they will follow.

There are three initial pathways for hospitalised individual $i$. Individual $i$ will either recover and be discharged from a ward bed, die in a ward bed, or move to an ICU bed; as the three pathways have different length of stay distributions they modelled as three separate compartments $H_R$, $H_D$ and $ICU_{pre}$. To determine which pathway individual $i$ will follow, we sample from,

$$X_h \sim \text{Categorical}(\mathbf{p}^i_1), \tag{19}$$

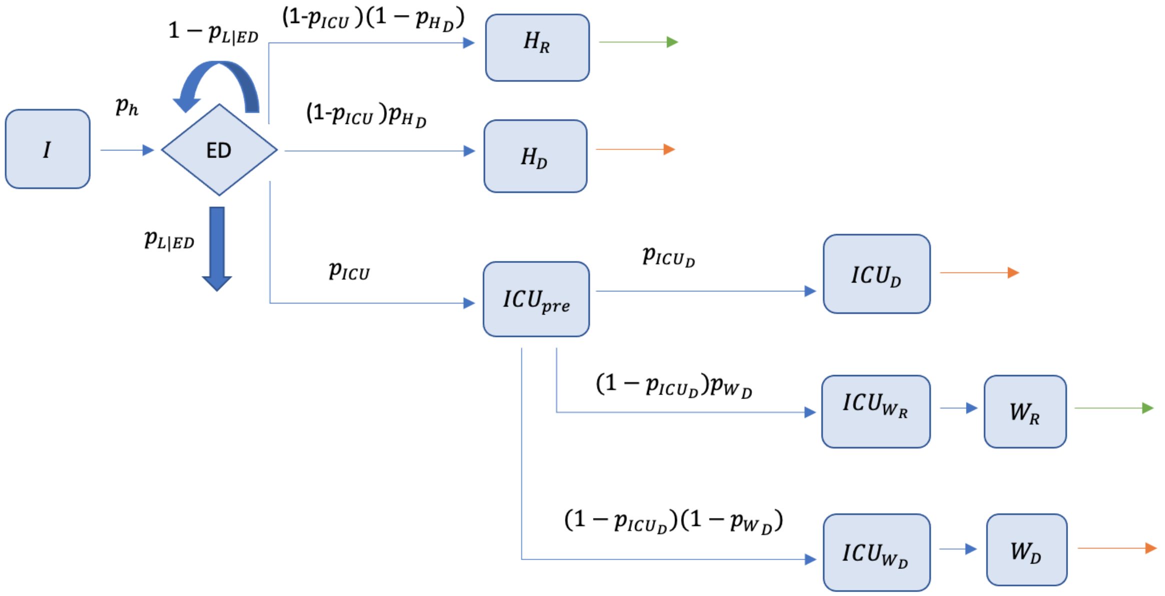

**Figure C1:** Depiction of the compartmental structure of the clinical pathways model.

where $X_h$ is the sampled hospital pathway,

$$\mathbf{p}_1^i = \left[p^i_{ICU|H}, (1 - p^i_{ICU|H}p^i_{H_D|ICU^c}), (1 - p^i_{ICU})(1 - p^i_{H_D|ICU^c})\right] \tag{20}$$

is a vector containing the probability of transitioning into $ICU_{pre}$, $H_D$, or $H_R$ respectively, $p^i_{ICU|H}$ is the probability that individual $i$ is admitted to ICU given they are hospitalised and is the probability that individual dies on ward given that they are in hospitalised and are not going to ICU. If individual $i$ requires the ICU, they follow a further ICU pathway to determine their final outcome.

The pathway through the ICU also consists of three different components. Within the ICU pathway an individual will either die in the ICU ($ICU_D$), die in a ward bed after leaving the ICU ($ICU_{WD}$), or recover and be discharged from a ward bed after leaving the ICU ($ICU_{WR}$). We sample which pathway is taken within the ICU from,

$$X_{ICU} \sim \text{Categorical}(\mathbf{p}_2^i), \tag{21}$$

where $X_{ICU}$ is the sampled ICU pathway,

$$\mathbf{p}_2^i = \left[p^i_{ICU_D|ICU}, (1 - p^i_{ICU_D|ICU}p^i_{W_D|ICU^c_D}), (1 - p^i_{ICU_D|ICU})(1 - p^i_{W_D|ICU^c_D})\right] \tag{22}$$

is a vector containing the probability of transitioning into the $ICU_D$, $ICU_{WD}$ or $ICU_{WR}$ compartment respectively, $p^i_{ICU_D|ICU}$ is the probability that individual $i$ dies in the ICU given they were admitted to ICU and $p^i_{WD|ICU^c_D}$ is the probability that individual of dies in a ward bed after leaving ICU without dying. For an individual that transitions into $ICU_{WR}$ or $ICU_{WD}$, they will move into a further ward compartment, $W_R$ or $W_D$, where they will either recover or die respectively.

Finally, the length of stay for individual $i$ in each compartment is sampled such that,

$$\tau_c \sim \text{Gamma}(\kappa_c, \theta_c), \quad (23)$$

where $\tau_c$ is the time spent in compartment $c$, and $\kappa_c$ and $\theta_c$ are the shape and rate parameter for compartment $c$ respectively. Uncertainty is incorporated by sampling rate and shape parameters from the posterior estimated for the Australian Delta wave [24].

By generating a clinical timeline for every symptomatic individual, we can calculate hospital admissions, ICU occupancy, ward occupancy and deaths by age in continuous-time. Furthermore, by explicitly incorporating the effects of neutralising antibodies on protection against each outcome, we are able to account for individual level immune responses. Note that we assume that neutralising antibody titre levels do not change the distribution of time spent in any compartment.

### D Cost-effectiveness analysis

#### D.1 CEA model overview

The cost-effectiveness model uses as inputs the outputs from the clinical pathways model linked to the mechanical agent-based model (ABM) that in turn, is linked to the infection transmission/dynamics model. A population of 100,000 is run through the models, over a three-year period. The first 1.5 years represents the main vaccination program (also referred to as prior or primary vaccination). The second 1.5 years represents the different boosting programs that are considered for the cost-effectiveness analysis, representing different emergence times, transmissibility, boosting times, and boosting frequency. The ABM and clinical pathways models provide scenario-specific mean estimates of vaccination doses delivered per 100,000 people, COVID-19 infections (all, symptomatic, hospital admissions and total hospital bed days occupied, intensive care unit (ICU) admissions and total ICU beds occupied), and COVID-19 related deaths by 10-year age groups.

The cost-effectiveness analysis has been conducted from the healthcare system perspective, including direct medical costs only. The main categories of costs included are (1) programmatic costs related to the vaccination intervention, including vaccine dose costs, wastage, and delivery costs; and (2) disease management costs at home, in outpatient and inpatient settings for symptomatic COVID-19 related illness. COVID-19 testing costs were not included. While these costs have been estimated to be substantial [25], they remain highly uncertain. In any resurgence, we estimate a much broader use of rapid antigen testing than polymerase chain reaction (PCR) for case ascertainment; thus, the historical use of testing strategies cannot inform future testing use. Furthermore, PCR capacity varies dramatically by country, and the use of different types of tests will likely vary by case numbers.

#### D.2 Defining exemplar country contexts for cost-effectiveness analysis

All countries in the WPR started COVID-19 vaccination programs in 2021. While countries had different vaccination strategies, in general first doses were assigned to frontline workers, at risk adults and the elderly, followed by the remaining adult population. Programs were expanded to include children aged 12 and above starting in early 2022. Most countries further expanded their vaccine policy to include children 5 years and older in mid-2022.

According to WHO data, 2-dose vaccine coverage varies significantly throughout the Western Pacific (Tables D1 and D2). High income, ‘older’ demographic countries tend to have higher vaccination coverage, ranging from 64.5% in New Caledonia to 87.4% in Singapore with a median of 84.5% in New Zealand (as a proportion of total population as of 22/12/22). Lower-middle and upper-middle income countries with younger demographics displayed a much wider range of vaccination coverage ranging from 3.6% in Papua New Guinea (PNG) to 101.9% in Brunei with a median coverage 67.4% in The Philippines. Booster coverage displayed a similar pattern.

In alignment with the two ABM populations, representing differing demographics within the WPR, we consider three key groupings of 'exemplar' countries in terms of: (1) demography (typical 'older' versus 'younger' population demographics); (2) health systems capacity and prior primary COVID-19 vaccine coverage rates (strong health systems and high prior primary vaccine coverage versus relatively weaker health systems and lower prior primary vaccine coverage); (3) income group level (high income versus upper-middle and lower-middle income); and (4) vaccination delivery unit costs and disease management costs.

The representative 'exemplar' countries groupings are as follows, with full details provided below (Table D1):

- **Group A**: High income country (HIC), 'older' population with strong health systems capacity and high ( 80%) prior primary vaccine coverage. High unit costs for vaccine delivery and disease management. (Countries in this group include Japan, Australia, Republic of Korea, and Hong Kong, and are representative of other high income countries in the WPR such as New Zealand)
- **Group B**: Upper- and lower- middle income country (MIC), 'younger' population with varying levels health systems capacity and prior primary vaccine coverage ( 80% and 50%). Low-to-high unit costs for vaccine delivery (depending on geography and population size) and low-to-middle unit costs for disease management. (Countries in this group include Fiji, Samoa, Tonga, Mongolia, Cambodia, Philippines, Lao, Vanuatu, Kiribati, Micronesia, PNG, and Solomon Islands)
- **Group C**: Lower-middle income country, younger population with weaker health systems capacity and low ( 20%) prior primary vaccine coverage. Low unit costs for vaccine delivery and disease management. (Countries in this group, a subgroup of Group B, include PNG and Solomon Islands)

Some WPR countries are not included in these representative 'exemplar' country groupings (for example, those with demographics that classify as neither 'older' nor 'younger', or those with demographics that match to 'older' or 'younger' categorisation, but per-capita income level does not). The implications for these countries would need to be considered in light of the findings for **Groups A** and **B**.

**Table D1:** Countries and areas in WHO Western Pacific Region (WPR) and characteristics.

| Country[2] | Income classification[3] | Younger/ Older | Population size[4] | 2 dose vaccination coverage[5] | Booster coverage[5] |
|---|---|---|---|---|---|

[2]Abbreviations: PDR, People's Democratic Republic; SAR, Special Administrative Region; USA, United States of America

[3]Income Classification data sourced from World Bank 22/12/22

[4]Population data from Our World in Data 22/12/22

[5]Data retrieved from WHO Coronavirus (COVID-19) Dashboard 22/12/22. Data on 2-dose coverage and booster coverage are estimated based on number of doses administered and total population size

| | | | | | |
|---|---|---|---|---|---|
| American Samoa | Upper middle | - | 45,035 | 75.05% | 43.77% |
| Australia | High | Older | 25,688,079 | 84.92% | 56.06% |
| Brunei Darussalam | High | Younger | 445,373 | 101.93% | 77.49% |
| Cambodia | Lower middle | Younger | 16,589,023 | 87.33% | 62.21% |
| China | Upper middle | Older | 1,412,360,000 | 86.82% | 54.7% |
| Cook Islands | - | - | 17,604* | 83.56% | 30.38% |
| Fiji | Upper middle | Younger | 924,610 | 71.33% | 18.78% |
| French Polynesia | High | Older | 304,032 | 66.23% | 39.95% |
| Guam (USA) | High | Older | 170,534 | 83.55% | 42.74% |
| Hong Kong | High | Older | 7,413,100 | - | - |
| Japan | High | Older | 125,681,593 | 81.43% | 66.67% |
| Kiribati | Lower middle | Younger | 128,874 | 61.86% | 19.61% |
| Lao PDR | Lower middle | Younger | 7,425,057 | 74.54% | 27.86% |
| Macao SAR | High | Older | 686,607 | | |
| Malaysia | Upper middle | - | 33,573,874 | 85.06% | 50.28% |
| Marshall Islands | Upper middle | - | 42,050 | 61.89% | 36.41% |
| Micronesia, Federates States of | Lower middle | Younger | 113,131 | 57.53% | 26.55% |
| Mongolia | Lower middle | Younger | 3,347,782 | 66.64% | 32.25% |
| Nauru | High | - | 12,511 | 79.24% | 46.75% |
| New Caledonia | High | Older | 271,030 | 64.49% | 32.96% |
| New Zealand | High | Older | 5,122,600 | 84.87% | 56.33% |
| Niue | - | - | 1,653* | 100.99% | 75.65% |
| Northern Mariana Islands Commonwealth of the USA | High | - | 49,481 | 78.2% | 42.11% |
| Palau | Upper middle | - | 18,024 | 101.07% | 71.11% |
| Papua New Guinea | Lower middle | Younger | 9,949,437 | 3.46% | 0.36% |
| Philippines | Lower middle | Younger | 113,880,328 | 67.42% | 19.38% |
| Pitcairn Island | - | - | 50* | 74% | 46% |
| Republic of Korea | High | Older | 51,744,876 | 87.17% | 65.63% |
| Samoa | Lower middle | Younger | 218,764 | 89.54% | 39.85% |
| Singapore | High | Older | 5,453,566 | 87.4% | 77.34% |
| Solomon Islands | Lower middle | Younger | 707,851 | 31.68% | 2.54% |

| | | | | | |
|---|---|---|---|---|---|
| Tokelau | - | - | 1,399* | 163.19% | 71.7% |
| Tonga | Upper middle | Younger | 106,017 | 72.72% | 36.24% |
| Tuvalu | Upper middle | - | 11,204 | 79.05% | 46.88% |
| Vanuatu | Lower middle | Younger | 319,137 | 42.75% | 5.39% |
| Vietnam | Lower middle | - | 97,468,029 | 87.89% | 59% |
| Wallis and Futuna | - | - | 10,749* | 62.15% | 28.46% |

In Table D1, Group A is marked in green, representing older populations with high income, strong health systems capacity and high prior primary coverage. Meanwhile, Group B, marked in yellow, represent younger populations with upper- and lower-middle incomes, middle-to-strong health systems capacity and middle-to-high prior primary vaccine coverage. Finally, Group C are marked in blue, represent younger populations with lower-middle income, weak health systems capacity and lower prior primary vaccine coverage. There are several countries that are not highlighted—these were not included in these representative 'exemplar' country groupings. The implications for these countries would need to be considered in light of the findings for Groups A and B, as they would likely sit somewhere in between these groupings.

**Table D2:** Median vaccination coverage by age demographic in the Western Pacific Region.

| Age Demographic | Median 2-dose coverage[6] | IQR | Range |
|---|---|---|---|
| Younger | 67.42% | 57.53% – 74.54% | 3.46% – 101.93% |
| Older | 84.87% | 81.43% – 86.82% | 64.49% – 87.4% |

#### D.3 Resource use and costs

Inputs and data sources for estimating costs of COVID-19 vaccination and disease management are presented in Table D4.

##### D.3.1 COVID-19 vaccine dose cost

COVID-19 vaccine price data were retrieved from the WHO COVID-19 vaccine price report [26]. This report summarizes vaccine dose price data based on the WHO MI4A COVID-19 Vaccine Purchase Database [27], which includes vaccine purchase data from public sources and data reported by countries through the WHO/UNICEF Joint Reporting Form (eJRF). Countries' names are not available in the dataset; however, the WHO region and income level are provided. Few countries in the Western Pacific Region (WPRO) had available price data, so our study has used global pricing data, by income group. Though AstraZeneca is no longer a preferred vaccine, we have included it in the economic model as

[6] as % of total population, as of 22/12/22

it was used widely in the Western Pacific region in 2021 and has the lowest price per dose across all vaccines.

The vaccine dose price used in the base case was the average price per dose for all vaccines: Pfizer BioN-Tech (Comirnaty), Moderna (mRNA-1273), Janssen (Ad26.COV 2-S), and AstraZeneca (Vaxzevria) by income group. For groups A and C, the ranges were the minimum and maximum prices for high- and lower-middle-income countries respectively. Group B comprises a mix of lower- and upper-middle income countries, so the range was obtained from the minimum and maximum vaccine prices of lower- and upper-middle countries combined (Table D4 and Table D3).

**Table D3:** Average price per vaccine (per dose, US$), by income group from WHO vaccine price report. Data from public sources and as reported by countries to WHO, up to March 2022.

| Income group | All vaccines | Pfizer | Moderna | Janssen | AstraZeneca |
|---|---|---|---|---|---|
| High income | $14.2 | $20.7 | $25.2 | $9.7 | $3.9 |
| Upper-middle income | $10.3 | $12.5 | $10.0 | $7.5 | $4.0 |
| Lower-middle income | $7.8 | $10.0 | $10.0 | $7.5 | $4.1 |

**Table D4:** Inputs for estimating COVID-19 vaccination and disease management costs.

| Parameter | Base case (range) | Source of data and rationale |
|---|---|---|
| *Cost per dose of vaccine ($)* | | |
| Group A (all high income) | 14.2 (3.9–25.2) | Data from WHO COVID-19 vaccine price report [26]. Base case is mean by income group across vaccines; range is the minimum and maximum by vaccine type. |
| Group B (all middle income) | 7.8 (4.0–12.5) | As for Group A |
| Group C (middle income, low cov) | 7.8 (3.9–25.2) | As for Group A |
| *Delivery cost per dose ($)* | | |
| Group A (80% coverage) | 23.1 (11.1–33.6) | Data from government reports [28, 29] or studies [30, 31]. Base case is average across Group A countries with cost data; range is minimum and maximum of costs. |
| Group B (50 or 80% coverage) | 9.8 (0.7–19.1) | Data from UNICEF reports [32, 33]. Base case assumes the cost at 70% coverage; range is minimum and maximum of costs |
| Group C (20% coverage) | 7.7 (2.5–10.5) | Data from UNICEF reports [32, 33]. Base case assumes double the cost at 70% coverage; range is minimum and maximum of costs |

| | | |
|---|---|---|
| *Proportion of doses wasted* | | |
| Group A (all high income) | 10% (0–20) | Data from UNICEF reports [32, 33]. Assumes range from no dose wasted to double the proportion of doses wasted. |
| Group B (all middle income) | 10% (0–20) | As for Group A |
| Group C (middle income, low cov) | 10% (0–20) | As for Group A |
| *Cost for non-hospitalized case ($)*[7] | | |
| Group A (all high income) | 75 (53–115) | Data from medical fee schedules [34, 35, 36, 37], studies [38], or WHO-CHOICE [39]. Base case is average of available cost data; usual range estimation above. |
| Group B (all middle income) | 52 (21–131) | Data from Torres-Rueda, et al. [25] |
| Group C (middle income, low cov) | 31 (28–35) | Data from Torres-Rueda, et al. [25] |
| *Cost for hospitalization without ICU, per day($)* | | |
| Group A (all high income) | 351 (209–657) | Data from medical fee schedules [34, 35, 36, 37], studies [38], or WHO-CHOICE [39]. Base case is average of available cost data; usual range estimation above. |
| Group B (all middle income) | 52 (34–99) | Data from Torres-Rueda, et al. [25] |
| Group C (middle income, low cov) | 41 (40–42) | Data from Torres-Rueda, et al. [25] |
| *Cost for hospitalization needing ICU, per day($)*[8] | | |
| Group A (all high income) | 2594 (825–4284) | Data from medical fee schedules [34, 35, 36, 37], studies [38], or WHO-CHOICE [39]. Base case is average of available cost data; usual range estimation above. |
| Group B (all middle income) | 543 (295–1273) | Data from Torres-Rueda, et al. [25] |

[7] Cost for non-hospitalized case includes two visits to a clinic

[8] Costs of hospitalized critical case from original report included both ICU and non-ICU bed days and thus have been inflated by 20% to represent the cost of an ICU bed day alone.

| | | |
|---|---|---|
| Group C (middle income, low cov) | 347 (341–353) | Data from Torres-Rueda, et al. [25] |
| *Cost per COVID-related death ($)* | | |
| Body bag | 65 (fixed) | Data from Torres-Rueda, et al. [25] |

###### D.3.2 COVID-19 vaccine delivery cost

The cost of COVID-19 vaccine delivery remains uncertain. Delivery costs will vary by vaccine type (including cold chain requirements), country health systems capacity, delivery mechanism and target population and coverage level. In most scenarios explored, a set number of doses (11,000) are delivered to a population of 100,000, equivalent to a coverage of 11% of the total population. While the different scenarios explored (high risk versus random versus paediatric boosting) likely have different delivery costs associated with them, given the underlying uncertainty, we assume the same delivery costs across all scenarios. Delivery costs are generally assumed to be U-shaped, decreasing as coverage increases due to shared fixed costs across a larger population being vaccinated, and increasing at very high coverage levels due to difficulties in vaccinating hard-to-reach populations. For Groups A and B countries, given high to moderate prior vaccination coverage, we assume booster doses would be delivered at the same unit cost per dose as the primary doses. We assume across all scenarios that delivery of booster doses even at low coverage levels, would incur the same cost of vaccine delivery as the primary doses.

**Group A ('older' population, high income countries)**

There are no consistent estimates of vaccine delivery costs for high-income countries. We sought to estimate or find COVID-19 vaccination delivery costs for a select number of countries in Group A where data were available, to use as inputs for the modelling. Delivery costs per dose for Hong Kong and Korea were taken from previous publications with assumed COVID-19 vaccination coverage rates of 72% and 80%, respectively [30, 31]. Delivery costs per dose for Japan were taken from the Japanese Government's National Treasury's burden for the vaccination measures against the COVID-19 report, at an unspecified coverage rate [28]. Delivery costs for Australia were calculated by dividing the Australian government's reported funding for COVID-19 vaccine distribution and administration in 2020–2022 by the total doses administrated up to mid-2022 (about 80% coverage) [29]. These unit delivery costs were used for the 80% and 50% coverage scenarios and multiplied by two to estimate the delivery costs at 20% coverage. We use the average cost across all estimates obtained for the base case delivery cost, and the minimum and maximum delivery cost estimates as upper and lower-bound ranges.

##### Group B and C ('younger' population, middle-income countries)

The COVID-19 vaccine delivery cost estimates used in modelling the 'younger' demographic populations were based on two recent UNICEF reports that provided estimates for low- and middle-income countries (LMICs) [32, 33]. These delivery costs refer to the costs associated with delivering vaccines to target populations exclusive of vaccine purchase costs. The costs estimated in both reports are financial costs, including (1) variable costs (e.g., cold chain equipment, per diem for outreach, personal protective equipment, vaccine transport, and management, etc.) and (2) fixed costs (i.e., handwash station, training, planning and coordination, social mobilization, pharmacovigilance, behavioural and social data collection). For this study, we assumed the economic costs required for a cost-effectiveness analysis, would be similar to the financial costs, and therefore used these estimates in the base case.

In the latest UNICEF report, COVID-19 vaccine delivery costs were estimated for countries achieving a 70% of total population coverage (equivalent to 92% coverage rate in population ≥12 years of age) in four different scenarios (leveraging fixed delivery sites, balancing human resource protection, protecting human resources partially, protecting human resources fully) [33]. In the earlier report, which focussed on achieving 20% coverage, the estimation was based on the leveraging fixed delivery sites scenario only. For consistency, we have chosen the delivery costs under this scenario as the base case, which assumed 10% of the available workforce allocated to delivery, 85% fixed site delivery, and 15% outreach delivery. The fixed-outreach proportion was close to the data for the Western Pacific region in 85 National Deployment and Vaccination Plans (86%–14%) [33]. In the leveraging scenario from the earlier UNICEF report, the average cost per dose delivery at 20% coverage was approximately double that of achieving 70% coverage, as fewer people shared fixed costs. Due to the lack of country-specific estimates at 20% coverage, we also assumed that the delivery costs at 20% coverage were double those at 70% coverage. We also assumed that the 50% and 85% coverage scenarios had the same unit delivery costs as the 70% coverage scenario. Delivery cost estimates used in the model are provided in Table D5.

**Table D5:** Vaccination delivery cost estimates (2020 USD) by country and initial primary vaccination coverage.

| Country | Group | 20% Coverage | 50% Coverage[9] | 70% Coverage |
|---|---|---|---|---|
| Australia | A | - | - | $22.3[10] |
| Japan | A | - | - | $33.60 |
| Korea, Rep. | A | - | - | $11.10 |
| Hong Kong | A | - | - | $25.30 |
| Fiji | B | - | * | $10.50 |
| Samoa | B | - | * | $17.80 |

[9] Entries marked with * are assumed to be the same as the 70% coverage

[10] Australia 2020-2022 vaccine program budget (including vaccine program implementation, administration and distribution) divided by total administrated doses up to mid-2022 = 1876.7 million /57.92 million ∼ 32.4 AUD ∼ 22.3 USD. Budget data from Minister for Finance of the Commonwealth of Australia; doses administration data from Our World in data 01/07/2022

| | | | | |
|---|---|---|---|---|
| Tonga | B | - | * | $19.10 |
| Mongolia | B | - | * | $13.40 |
| Cambodia | B | - | * | Not available[11] |
| Lao PDR | B | - | * | $1.30 |
| Philippines | B | - | * | $0.70 |
| Vanuatu | B | - | * | $5.70 |
| Kiribati | B | - | * | $29.3[12] |
| Micronesia, Fed. Sts. | B | - | * | $23.1[12] |
| Papua New Guinea | B & C | $5.0[13] | - | $2.50 |
| Solomon Islands | B & C | $10.5[13] | - | $5.20 |

##### D.3.3 COVID-19 treatment cost

###### Group A ('older' population, high income countries)

Detailed cost estimates for management of COVID-19 infections were estimated for a small number of countries in Group A, where these were readily available. Based on costing methods by Torres-Rueda et al. [25] as described below and used for middle-income countries (Groups B and C), we used the Australian medical fee schedules and publicly available government data to calculate the three types of case management costs [35, 37]. We used the same method to estimate the case management costs in Japan by applying the Japanese medical fee schedule [34]. Of note, in the home-based cases in HICs, we have excluded the home-based bed-day cost due to lack of detailed costing method in the reference article. Also, in hospital-based critical cases, we dropped the general ward bed-day input and changed the number of units per input for ICU bed-day from 0.66 to 1. Malaria testing was included in all LMICs hospital-based cases, but we removed it from the costs for HICs, given that HICs are predominantly low-prevalence malaria regions where this testing may not be a routine admission test. The costs of inpatient cases in Hong Kong were taken from a cost-effectiveness study of the COVID-19 vaccine in Hong Kong, with the source being the public charges for non-eligible persons [31, 36]. The costs of hospitalisation cases in Korea were obtained from a COVID-19 cost-effectiveness analysis in Korea, which employed the cost estimations by Korea Disease Control and Prevention Agency [38]. The outpatient costs for home-based care in Korea were based on WHO CHOICE unit costs [39], which we adjusted for inflation and currency conversion.The cost per COVID-19 related death only includes the cost of a body bag based on the study by Torres-Rueda et al. [25], and thus is likely to be underestimated.

[11] Cambodia delivery costs were not available in UNICEF report.

[12] Costs of delivery for Micronesia and Kiribati were excluded from the mean delivery cost estimate for the Group B, younger, populations, due to their small population size (∼125,000) and high delivery costs.

[13] Assumed to be 2× delivery cost estimates at 70% coverage, based on UNICEF report 2021-2022 in which average delivery cost across LMICs at 20% coverage was double the cost at 70% coverage in leveraging fixed delivery sites scenario.

**Group B and C ('younger' population, middle-income countries)**

All disease costs for LMICs were available directly from a model-based cost estimations study [25]. The study used data from three LMICs (Ethiopia, Pakistan, and South Africa) as the model references to extrapolate the case management costs for home-based care, hospitalisation for severe care, and critical care across all LMICs. The original costs reported in the study were inflated to 2020 USD.

Home-based care costs are defined as the cost per mild-to-severe case requiring home-based care, including (1) the cost of home-based care bed-day; (2) the cost of community-based care via a clinician's visit. The number of bed-day and clinician visit was set at 5 and 2, respectively.

Hospitalised severe care costs were calculated per case and per day, including (1) general ward bed-day; (2) diagnostics. Hospitalised critical care costs were also presented per case and per day. Compared with severe cases, the additional costs per case per day were: (1) ICU bed-day; (2) additional resourcing per COVID-related complication. However, as the modelled epidemiological data is presented by ICU admission (rather than combining a patient who has received ICU and general ward care) the cost shown in this report likely underestimates the actual cost per day of a patient treated in an ICU. As general ward costs were considered representative of one-third of the bed day costs, we conservatively inflated the bed day cost by 20% when applying these costs in the economic model. Further clarification is presented in Tables D6 and D7.

**Table D6:** Disease management unit costs (2020 USD) by country.

| Country | Group | Non-hospitalised, per case | Hospitalised without ICU, per day | Hospitalised with ICU, per day | Death, per case |
|---|---|---|---|---|---|
| Australia | A | $53.50 | $271.60 | $4,284.30 | $64.50 |
| Japan | A | $54.00 | $208.70 | $2,120.50 | $64.50 |
| Korea, Rep. | A | $76.80 | $267.00 | $825.00 | $64.50 |
| Hong Kong | A | $114.70 | $657.20 | $3,144.30 | $64.50 |
| Fiji | B | $131.10 | $99.00 | $1,272.70 | $64.50 |
| Philippines | B | $47.00 | $48.80 | $410.70 | $64.50 |
| Samoa | B | $82.10 | $73.10 | $1,005.90 | $64.50 |
| Tonga | B | $79.00 | $70.20 | $964.30 | $64.50 |
| Mongolia | B | $59.80 | $53.00 | $441.80 | $64.50 |
| Cambodia | B | $22.90 | $35.90 | $308.20 | $64.50 |
| Lao PDR | B | $37.30 | $42.80 | $362.80 | $64.50 |
| Vanuatu | B | $37.90 | $43.90 | $370.80 | $64.50 |
| Kiribati | B | $20.80 | $34.30 | $295.10 | $64.50 |
| Micronesia, Fed. Sts. | B | $42.60 | $46.10 | $388.30 | $64.50 |
| Papua New Guinea | C | $34.80 | $41.70 | $353.50 | $64.50 |
| Solomon Islands | C | $28.00 | $39.90 | $340.90 | $64.50 |

**Table D7:** Units input and unit costs (2020 USD) for Japan and Australia.

| | Number of units per input | Japan | Australia |
|---|---|---|---|
| *Home-based care* | per case | | |
| Community-based care via clinical visit | 2 | $27.00 | $26.70 |
| *Total* | | $54.00 | $53.50 |
| *Hospital-based (severe)* | per case/day | | |
| Inpatient ward bed-day (severe) | 1 | $196.70 | $264.10 |
| Chest X-ray | 0.125 | $19.70 | $24.80 |
| Full blood count (including haemoglobin test) | 0.125 | $27.70 | $11.70 |
| Blood urea and electrolyte test (including C-reactive protein test) | 0.125 | $22.20 | $12.20 |

| | | | |
|---|---|---|---|
| HIV test | 0.125 | $26.20 | $10.80 |
| *Total* | | $208.70 | $271.60 |
| *Hospital-based (critical)* | per case/day | | |
| ICU bed-day | 1 | $1,359.10 | $3733.80 |
| Chest X-ray | 10 | $19.70 | $24.80 |
| Full blood count (including haemoglobin test) | 10 | $27.70 | $11.70 |
| Blood urea and electrolyte test (including C-reactive protein test) | 10 | $22.20 | $12.20 |
| Venous blood gas test | 10 | $4.20 | $4.20 |
| HIV test | 0.1 | $26.20 | $10.80 |
| Acute respiratory distress syndrome | 0.47 | $22.50 | $22.50 |
| Acute kidney injury days | 0.04 | $10.60 | $10.60 |
| Acute cardiac injury days | 0.06 | $46.30 | $46.30 |
| Liver dysfunction days | 0.06 | $89.30 | $89.30 |
| Pneumothorax days | 0.01 | $7.00 | $7.00 |
| Hospital-acquired pneumonia days | 0.05 | $18.90 | $18.90 |
| Bacteraemia days | 0.01 | $32.60 | $32.60 |
| Urinary tract infection days | 0.01 | $9.00 | $9.00 |
| Septic shock days | 0.05 | $0.80 | $0.80 |
| *Total* | | $2,120.50 | $4284.30 |

#### D.4 Health Outcomes

Health outcomes were presented as disability-adjusted life-years (DALYs) for each modelled scenario. DALYs were calculated as the sum of years of life lost (YLLs) and years lived with disability (YLDs).

##### Years of life lost (YLL)

YLLs following a premature death due to COVID-19 were calculated as the sum of the number of deaths ($N$) multiplied by life expectancy ($L$) for the age at death. We obtained the number of deaths and age at death (in ten-year age groups, up to 80 years plus) from the epidemiological model for each scenario. These were multiplied by a reference life expectancy for each exemplar country groupings from WHO lifetables for each 10-year age band. For Group A, we use the Japan life table given 'older' high-income countries in WPR have higher life expectancies than the global high-income country lifetable. For Groups B and C, we use the global lower-middle income lifetable, given 'younger' countries in WPR have a lower life expectancy than global upper-middle income and the WPR life table. In the base case, we discounted future YLLs at 3% annually according to the following formula:

$$YLL = \frac{N(1 - e^{-0.03L})}{0.03}. \tag{24}$$

##### Years lived with disability (YLD)

The YLD component was calculated for the acute phase of the disease and post-acute consequences following severe disease. We do not include long-COVID due to a lack of available data to specify this condition. We classified cases into four following mutually exclusive categories: asymptomatic, symptomatic non-hospitalized, hospitalized without ICU stay, and hospitalized with ICU stay. We specified an illness severity pathway for each category consisting of four health states (mild/moderate, severe, critical, and post-acute). The post-acute phase refers to the recovery period following hospitalisation, and has been expressed in other cost-effectiveness models [40]. Based on the illness severity pathway for these categories (Table D8), we calculated YLDs by summing up the product of time spent in each health state, the disability weight for that state, and the number of incident cases. YLDs have been calculated using the following formula:

$$YLD = \sum_i I_i \times L_i \times DW_i, \tag{25}$$

where $i$ is an index for health state, $I_i$ is the number of incident cases for each health state, $L_i$ is the duration of disability in years, and $DW_i$ is the disability weight. The duration of illness for each state was based on the average length of hospital and ICU stay from the literature (Table D8).

**Table D8:** Health states, duration of illness, and disability weights for calculating years lived with disability.

| **COVID-19 patient category** | **Health state** | **Days in state Base case (range)** | **Disability weight Base case (range)** | **Notes and sources** |
|---|---|---|---|---|
| A. asymptomatic cases | Not applicable | Not applicable | Not applicable | Zero disability for asymptomatic cases. |
| B. symptomatic non-hospitalized | Mild or moderate | 7.0 (2.0–9.5) | 0.051 (0.032–0.074) | GBD 2019 disability weight for moderate lower respiratory infections [41]. Days in mild/moderate state from literature [40, 42]. Assume no post-acute phase. |

| | | | | |
|---|---|---|---|---|
| C. hospitalized without ICU stay | Mild or moderate | 7.0 (2.0–9.5) | 0.051 (0.032–0.074) | Disability weights from GBD 2019 [41] for moderate lower respiratory infections severe lower respiratory infections and post-acute consequences (fatigue, emotional lability, insomnia) for infectious disease. Using 7 days from symptom onset to hospital, same as category B patients and a Belgian study [43]. Using same number of hospital days (5 days) used in category D patients [44]. Assume 1-week post-acute phase +/- 50% [40]. |
| | Severe | 5.0 (3.0–9.0) | 0.133 (0.088–0.190) | |
| | Post-acute | 7.0 (3.5–10.5) | 0.219 (0.148–0.308) | |
| D. hospitalized with ICU stay | Mild or moderate | 7.0 (2.0–9.5) | 0.051 (0.032–0.074) | Disability weights from Nomura 2021 (critical) [45] and GBD 2019 (others) [41]. Symptom onset to ICU discharge of 18 days [42]. Out of this number, assume 7 days from symptom to hospital [43] 5 days in hospital [44] and 7 days in ICU [44]. Assume 2 weeks post-acute phase +/- 50% [40]. |
| | Severe | 5.0 (3.0–9.0) | 0.133 (0.088–0.190) | |
| | Critical | 7.0 (4.0–11.0) | 0.675 (0.506–0.822) | |
| | Post-acute | 7.0 (3.5–10.5) | 0.219 (0.148–0.308) | |

#### D.5 Cost-effectiveness analysis

##### D.5.1 Cost-effectiveness thresholds

Cost-effectiveness thresholds (CETs) for the country groups were based on estimates by Woods et al [46]. This study estimated these thresholds for several countries using the opportunity cost of additional costs incurred from interventions, the relationship between country gross domestic product (GDP) per capita, and the value of a statistical life. These CETs were calculated in 2013 US dollar values. The study reports the lower and upper bounds (limits) of the CETs (per quality-adjusted life-year gained) as percentage of GDP per capita in 2013. Ochalek et al. [47] estimated CETs per DALYs averted, but only three countries in our study were included (Cambodia, Mongolia, and Philippines).

To obtain CETs for use in this study, we multiplied the CETs as percentage of GDP per capita with GDP per capita in 2020 from the World Bank [48], rounded to the nearest hundred. The estimates are presented in Table D9. Thereafter, we calculated the average threshold (separately for the lower and upper bounds) for each group. The CET was between $19,000 and $30,000 for Group A, between $200 and $1600 for Group B, and between $100 and $1000 for Group C countries. The thresholds for each country within the groups are presented in Table D10.

**Table D9:** Estimated willingness to pay thresholds.

| Age demographics / prior primary vaccine coverage | Representative countries / Areas | Threshold range (2020 USD) |
|---|---|---|
| Group A: Older population (all countries, high vaccination coverage) | Japan, Australia, Republic of Korea, Hong Kong, Brunei Darussalam, New Zealand, and Singapore | \$19,000–\$30,000 |
| Group B: Younger population (all countries, varying vaccine coverage) | Fiji, Samoa, Tonga, Mongolia, Cambodia, Philippines, Lao, Vanuatu, Kiribati, Micronesia, PNG, and Solomon Islands | \$200–\$1,600 |
| Group C: Younger population (low vaccine coverage) | PNG and Solomon Islands | \$100–\$1,000 |

**Table D10:** Willingness To Pay Thresholds for the Western Pacific Region.

| Country | GDP per capita in 2020 (A)[14] | % of GDP per capita Woods lower bound (B1)[15] | % of GDP per capita Woods upper bound (B2)[15] | Cost-effectiveness threshold, Woods lower bound (A*B1)[16] | Cost-effectiveness threshold, Woods upper bound (A*B2)[16] |
|---|---|---|---|---|---|
| Japan | \$40,193 | 48.30% | 48.50% | \$19,428 | \$19,512 |
| Australia | \$51,693 | 48.10% | 61.20% | \$24,855 | \$31,651 |
| Korea, Rep. | \$31,631 | 45.00% | 50.50% | \$14,228 | \$15,968 |
| Hong Kong | \$46,324 | 45.30% | 75.00% | \$20,999 | \$34,741 |
| Fiji | \$5,058 | 10.50% | 47.70% | \$530 | \$2,410 |
| Samoa | \$4,068 | 6.60% | 47.00% | \$267 | \$1,911 |
| Tonga | \$4,625 | 7.50% | 51.40% | \$348 | \$2,376 |
| Mongolia | \$4,061 | 12.40% | 47.80% | \$505 | \$1,939 |
| Cambodia | \$1,544 | 4.30% | 51.10% | \$67 | \$789 |
| Philippines | \$3,299 | 8.90% | 49.50% | \$294 | \$1,633 |
| Lao PDR | \$2,630 | 6.20% | 46.50% | \$162 | \$1,223 |

[14] Sourced from World Bank

[15] CETs were estimated based empirical estimates collected using marginal costs invested and marginal health outcomes across different NHS jurisdictions (k) assumed VSL = value of a life year = income elasticity for QALY, If similar elasticity for v and k exists than estimates were created based on differing GDP income elasticities.

[16] Estimate based on GDP per capita in 2020.

| | | | | | |
|---|---|---|---|---|---|
| Vanuatu | $2,870 | 4.70% | 57.00% | $135 | $1,637 |
| Papua New Guinea | $2,757 | 2.70% | 39.30% | $76 | $1,084 |
| Solomon Islands | $2,251 | 2.50% | 44.60% | $57 | $1,005 |
| Kiribati | $1,654 | 2.50% | 49.40% | $41 | $817 |
| Micronesia, Fed. Sts. | $3,565 | 5.40% | 55.00% | $193 | $1,960 |
| China | $10,435 | 16.40% | 64.80% | $1,711 | $6,763 |
| Guam | $34,624 | - | - | - | - |
| Brunei Darussalam | $27,443 | 35.90% | 87.80% | $9,854 | $24,102 |
| New Zealand | $41,441 | 47.80% | 50.30% | $19,821 | $20,847 |
| Singapore | $59,798 | 39.20% | 108.30% | $23,452 | $64,767 |

##### D.5.2 Cost-effectiveness results and interpretation

We present the results as incremental cost-effectiveness ratios (ICERs) for the modelled vaccination options compared to a counterfactual of no further vaccination. These ICERs are presented on a cost-effectiveness plane, which shows the DALYs averted and additional costs for the vaccination scenarios (described above) compared to the cost-effectiveness thresholds for the three scenarios. Results that fall below these thresholds indicate a vaccination strategy that is likely to be cost-effective. We have expressed these costs and outcomes per 100,000 people.

We have also performed one-way sensitivity analyses to determine the impact of various cost (vaccine prices and delivery costs) and epidemiological variables on the ICER. We have presented the results of these analyses on tornado diagrams, which indicates the change in ICER when varying parameters (see Figures E9, E5, E6), thus accounting for parameter uncertainty. For Groups B and C countries, we also explore a vaccine price of $0 per dose in a scenario analysis, to explore the cost-effectiveness from a government perspective of having donated vaccines available to some middle-income countries. Given the uncertainty of home-based care cost, we explore the cost-effectiveness if there is no home-based care cost in the scenario analysis.

Additionally, as we are already varying several key epidemiological and demographic parameters (for example, $R_0$ through high/low transmission scenarios and younger vs older populations), we can compare these parameters' influence on CE results to the costing parameters. This process has demonstrated apparent differences in CEA by allocation strategy across that broad variation, so it is unlikely that the conclusions of a more detailed uncertainty analysis would vary substantially.

##### D.5.3 Limitations

There are several limitations of this analysis that warrant mentioning.

1. We do not include testing costs, as previously explained. Testing costs can represent a substantial proportion of total costs related to COVID-19 in some countries, however these remain highly uncertain particularly across modelled future scenarios. It is unclear what impact the exclusion of testing costs would have on findings.
2. While indirect costs due to COVID-19, such as productivity losses, also make up a large proportion of total costs related to COVID-19, these costs have not been included in the current analysis. Accounting for indirect costs would make additional boosting vaccination programs appear more cost-effective than our findings indicate. In future work, a societal perspective may be considered.
3. We have not accounted for vaccine-related side effects, including both the costs and health impacts. These are unlikely to impact on cost-effectiveness findings.
4. We are currently not accounting for the costs or health impacts of long-COVID, due to data limitations.

### E Supplementary results

In this section, we have extended results with further details on scenarios from the main paper and additional scenarios. All scenarios and their associated figures and tables are noted in Table E1.

**Table E1:** Scenario summary table.

| Population type | Transmission potential | Vaccine coverage | Immune escape | Boosting strategy | Figures/Tables |
|---|---|---|---|---|---|
| older | high TP | high | 1.5 yrs | at 2 yrs, pediatric/high-risk/random boosting | Figs. 2(a), 3(a), 3(c), E1(a), E1(c), E5(b), Tab. 2(a) |
| younger | high TP | high | 1.5 yrs | at 2 yrs, pediatric/high-risk/random boosting | Figs. 2(b), 3(b), 3(d), E1(b), E1(d),Tab. 2(b) |
| older | high TP | high | 2.5 yrs | at 2 yrs, pediatric/high-risk/random boosting | Figs. 2(c), 3(a), 3(c), E2(a), E2(c), Tab. 2(c) |
| younger | high TP | high | 2.5 yrs | at 2 yrs, pediatric/high-risk/random boosting | Figs. 2(d), 3(b), 3(d), E2(b), E2(d), E6, Tab. 2(d) |
| younger | high TP | high | 1.5/2.5 yrs | high-risk boosting at 2 yrs, different costings | Fig. E7(a) |
| younger | low TP | high | 1.5/2.5 yrs | high-risk boosting at 2 yrs, different costings | Fig. E7(b) |
| older | low TP | high | 1.5 yrs | at 2 yrs, pediatric/high-risk/random boosting | Figs. E3(a), E4, E5(b), Tab. E2(a) |
| older | low TP | high | 2.5 yrs | at 2 yrs, pediatric/high-risk/random boosting | Figs. E3(b), E4, Tab. E2(b) |
| older | high TP | high | 1.5 yrs | at 1.75/2/2.25/2.5 yrs or half-yearly | Figs. 4(a), E8(a), Tab. E3(a) |
| younger | high TP | high | 1.5 yrs | at 1.75/2/2.25/2.5 yrs or half-yearly | Figs. 4(b), E8(b), E9(a), Tab. E3(b) |
| older | high TP | high | 2.5 yrs | at 1.75/2/2.25/2.5 yrs or half-yearly | Figs. 4(c), E8(c), Tab. E3(c) |
| younger | high TP | high | 2.5 yrs | at 1.75/2/2.25/2.5 yrs or half-yearly | Figs. 4(d), E8(d), E9(b), Tab. E3(d) |
| older | high TP | high | 2 yrs | at 2 yrs, 65+/55+/45+/35+/25+/16+/5+ yrs boosting | Figs. 5(a), 5(c), Tab. E4(a) |
| younger | high TP | high | 2 yrs | at 2 yrs, 65+/55+/45+/35+/25+/16+/5+ yrs boosting | Figs. 5(b), 5(d), Tab. E4(b) |
| older | low TP | high | 2 yrs | at 2 yrs, 65+/55+/45+/35+/25+/16+/5+ yrs boosting | Figs. E10(a), E11(a) Tab. E4(c) |
| younger | low TP | high | 2 yrs | at 2 yrs, 65+/55+/45+/35+/25+/16+/5+ yrs boosting | Figs. E10(b), E11(b), Tab. E4(d) |
| younger | high TP | low | 2 yrs | at 2 yrs, pediatric vaccination/general vaccination/high-risk boosting | Figs. 6(a), 6(c), 6(d), E14, Tab. E5(a) |
| younger | high TP | medium | 2 yrs | at 2 yrs, pediatric vaccination/general vaccination/high-risk boosting | Figs. 6(b), 6(c), 6(d), Tab. E5(b) |
| younger | low TP | low | 2 yrs | at 2 yrs, pediatric vaccination/general vaccination/high-risk boosting | Figs. E12(a), E13(a), E14, Tab. E5(c) |
| younger | low TP | medium | 2 yrs | at 2 yrs, pediatric vaccination/general vaccination/high-risk boosting | Figs. E12(b), E13(b), Tab. E5(d) |
| younger | high TP | low | 2 yrs | at 2 yrs, high-risk monovalent/bivalent boosting | Fig. E15(a), Tab. E6(a) |
| younger | high TP | medium | 2 yrs | at 2 yrs, high-risk monovalent/bivalent boosting | Fig. E15(b), Tab. E6(b) |
| younger | low TP | low | 2 yrs | at 2 yrs, high-risk monovalent/bivalent boosting | Fig. E16(a), Tab. E6(c) |
| younger | low TP | medium | 2 yrs | at 2 yrs, high-risk monovalent/bivalent boosting | Fig. E16(b), Tab. E6(d) |

#### E.1 High vaccination coverage scenarios: comparing target use groups - extended results

This set of scenarios (Section 3.1.1 in the main paper) considered allocating additional boosting to different target use groups in either pediatric, high-risk, or random boosting strategies. Figures E1 and E2 show the probability sensitivity analysis for selected scenarios from the main text, showing how high-risk boosting is largely cost-effective, while pediatric boosting is largely not cost-effective when considering economic uncertainty.

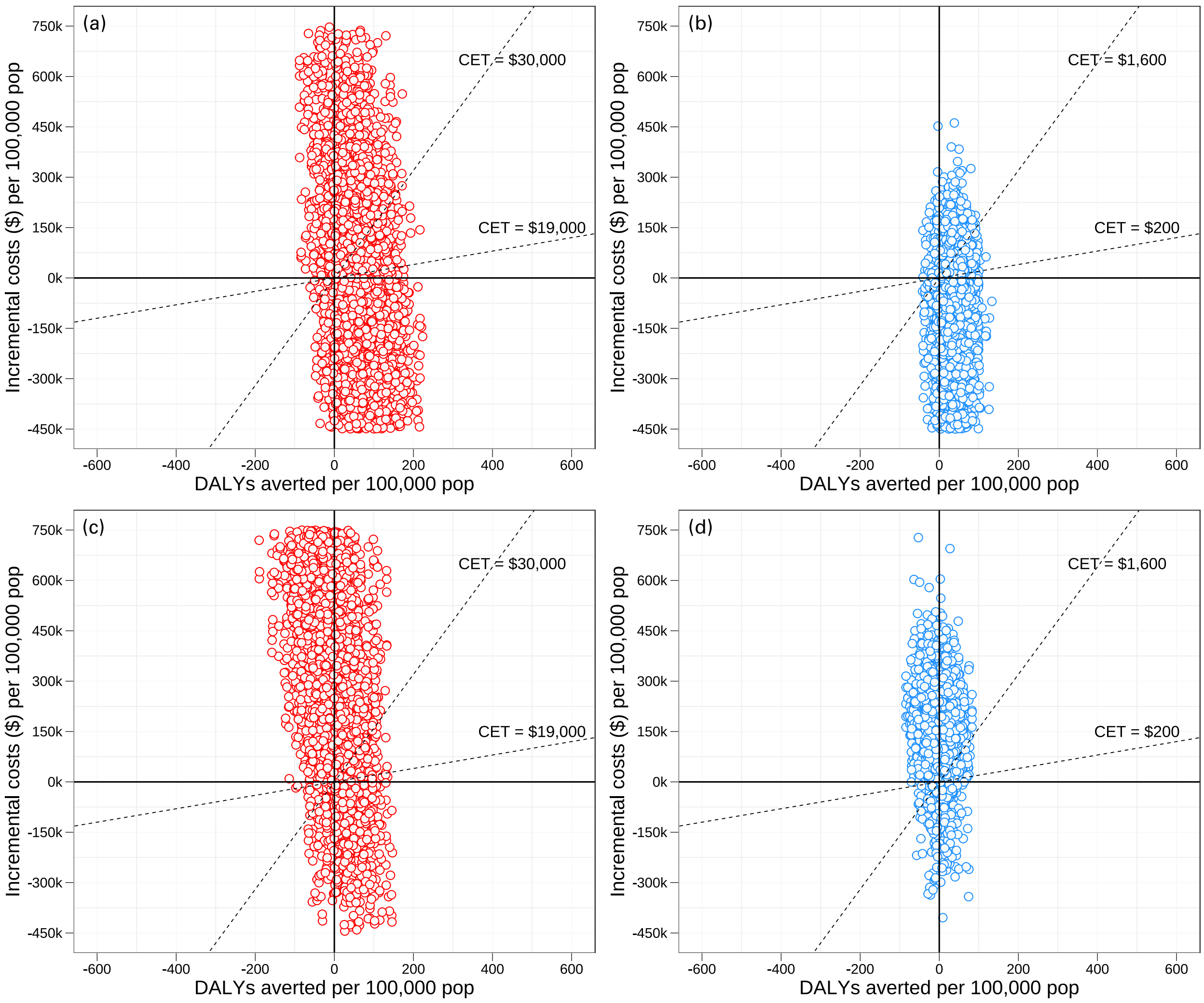

**Figure E1:** Probability sensitivity analysis of cost-effectiveness in the high transmission, high vaccination coverage setting, for older and younger demographics. Scenarios are run with boosting strategies at 2 years, with immune escape at 1.5 years. The dotted lines represent cost effective thresholds. **(a)** high risk boosting in the older population; **(b)** high risk boosting in the younger population; **(c)** pediatric boosting in the older population; **(d)** pediatric boosting in the younger population.

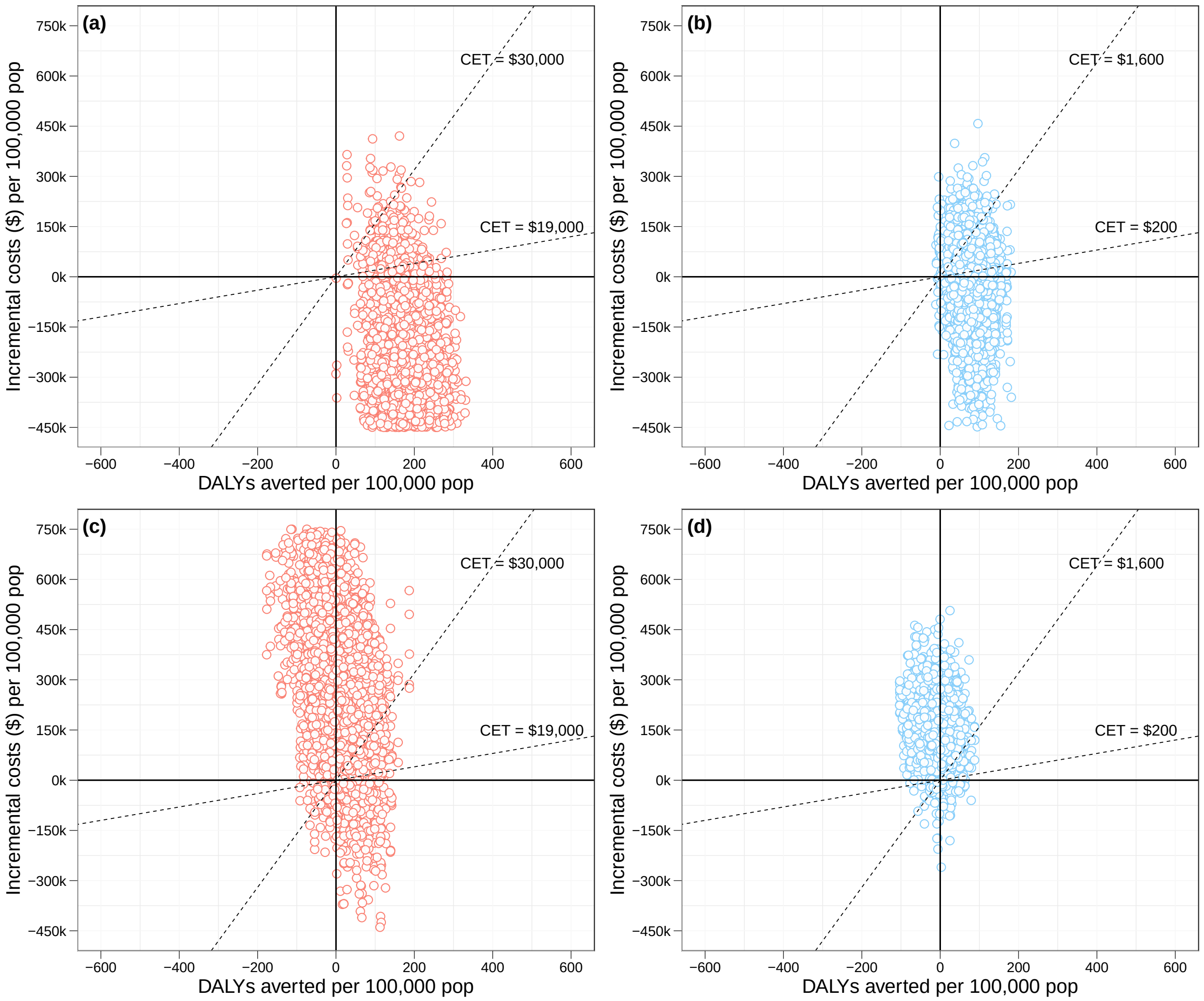

**Figure E2:** Probability sensitivity analysis of cost-effectiveness in the high transmission, high vaccination coverage setting, for older and younger demographics. Scenarios are run with boosting strategies at 2 years, with immune escape at 2.5 years. The dotted lines represent cost effective thresholds. **(a)** high risk boosting in the older population; **(b)** high risk boosting in the younger population; **(c)** pediatric boosting in the older population; **(d)** pediatric boosting in the younger population.

In the low transmission setting, Figure E3 shows the epidemic infection curves, Figure E4 shows the cost-effectiveness analysis, while Table E2 shows the median deaths. We find that high-risk boosting is likely to be cost-effective in the low-transmission setting too.

Figure E5 shows the one-way sensitivity analysis of a high-risk boosting strategy in the high and low transmission settings, with immune escape at 1.5 years. We find that vaccine delivery cost and unit cost per dose are the most influential costing parameters for older population scenarios.

Figure E6 shows the one-way sensitivity analysis for high-risk boosting, this time in the high-transmission high-vaccination setting with a younger demographic, with immune escape at 2.5 years. Here, we find that home-based care cost, vaccine delivery cost, and vaccine cost per dose are the most influential

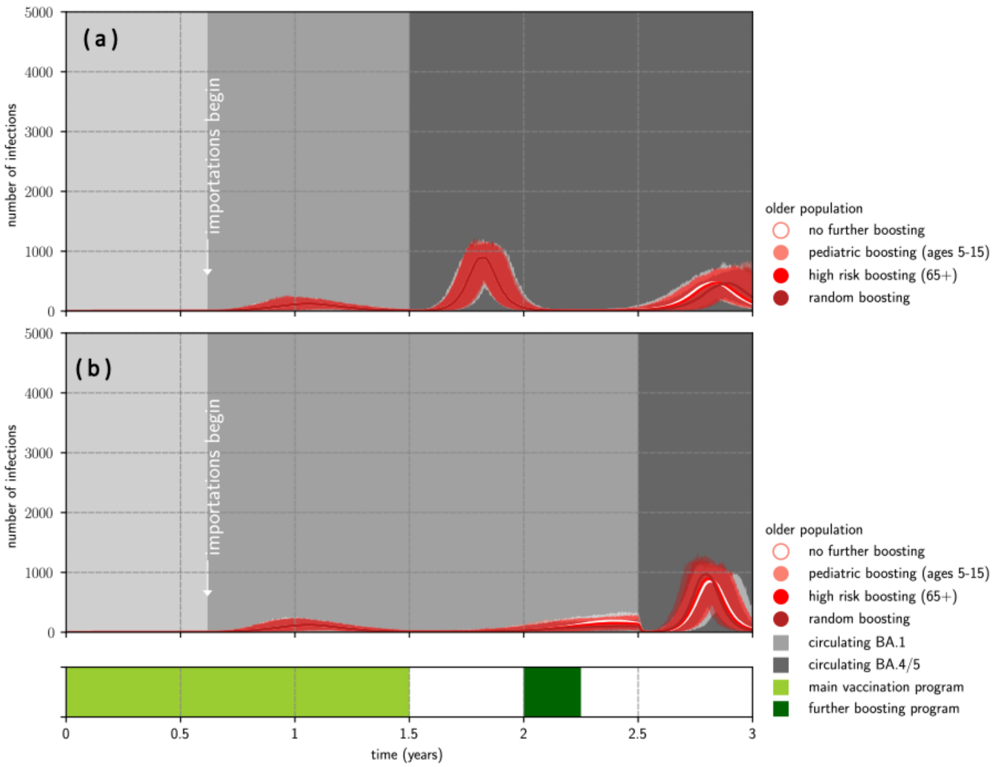

**Figure E3:** Outbreaks in the low transmission, high vaccination coverage setting, for older demographics. **(a)** epidemic waves given early immune escape (1.5 years); **(b)** epidemic waves given late immune escape (2.5 years). Scenarios are run with no further boosting, pediatric, high-risk and random boosting strategies at 2 years. The solid lines represent the pointwise median infections from 1000 simulations and the shaded regions represent the pointwise maximum and minimum infections.

costing parameters.

Figure E7 compares the cost-effectiveness of high-risk boosting in younger population, in four different scenarios (base, vaccine donated, no home care cost, vaccine donated and no home care cost). When vaccines are donated (i.e. vaccine dose costs are zero), high-risk boosting may be more cost-effective than non-donated scenarios. If home-based care costs are zero, then high-risk boosting is likely to be *not* cost-effective. If both vaccine dose costs and home care costs are zero, high-risk boosting may be cost-effective when immune escape starts 2.5 years.

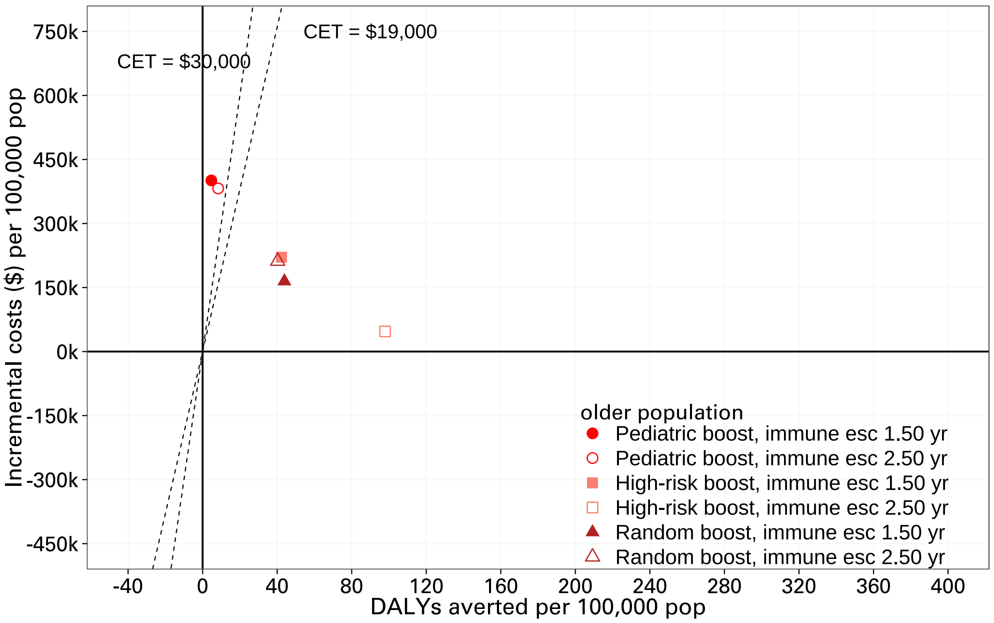

**Figure E4:** Cost-effectiveness analysis in the low transmission, high vaccination coverage setting, for older demographics. Scenarios are run with no further boosting, pediatric, high-risk and random boosting strategies at 2 years. The dotted lines represent cost effective thresholds.

**Table E2:** Median deaths (with 0.025 and 0.975 quantiles) between 1.5–3 years in the low-transmission high vaccination coverage setting, for older demographics, comparing target use groups. Early immune escape occurs at 1.5 years; late immune escape occurs at 2.5 years. The different boosting strategies run at 2 years.

| | Transmission potential | Immune escape | Boosting strategy | Median deaths |
|---|---|---|---|---|
| (a) | low TP | early | no further boosting | 39.0 (28.0, 51.0) |
| | low TP | early | pediatric boosting (ages 5–15) | 39.0 (28.0, 51.0) |
| | low TP | early | high-risk boosting (65+) | 36.0 (25.0, 48.0) |
| | low TP | early | random boosting | 36.0 (25.0, 48.0) |
| (b) | low TP | late | no further boosting | 31.0 (20.0, 43.0) |
| | low TP | late | pediatric boosting (ages 5–15) | 30.0 (19.0, 42.02) |
| | low TP | late | high-risk boosting (65+) | 23.0 (14.0, 34.0) |
| | low TP | late | random boosting | 28.0 (18.0, 40.0) |

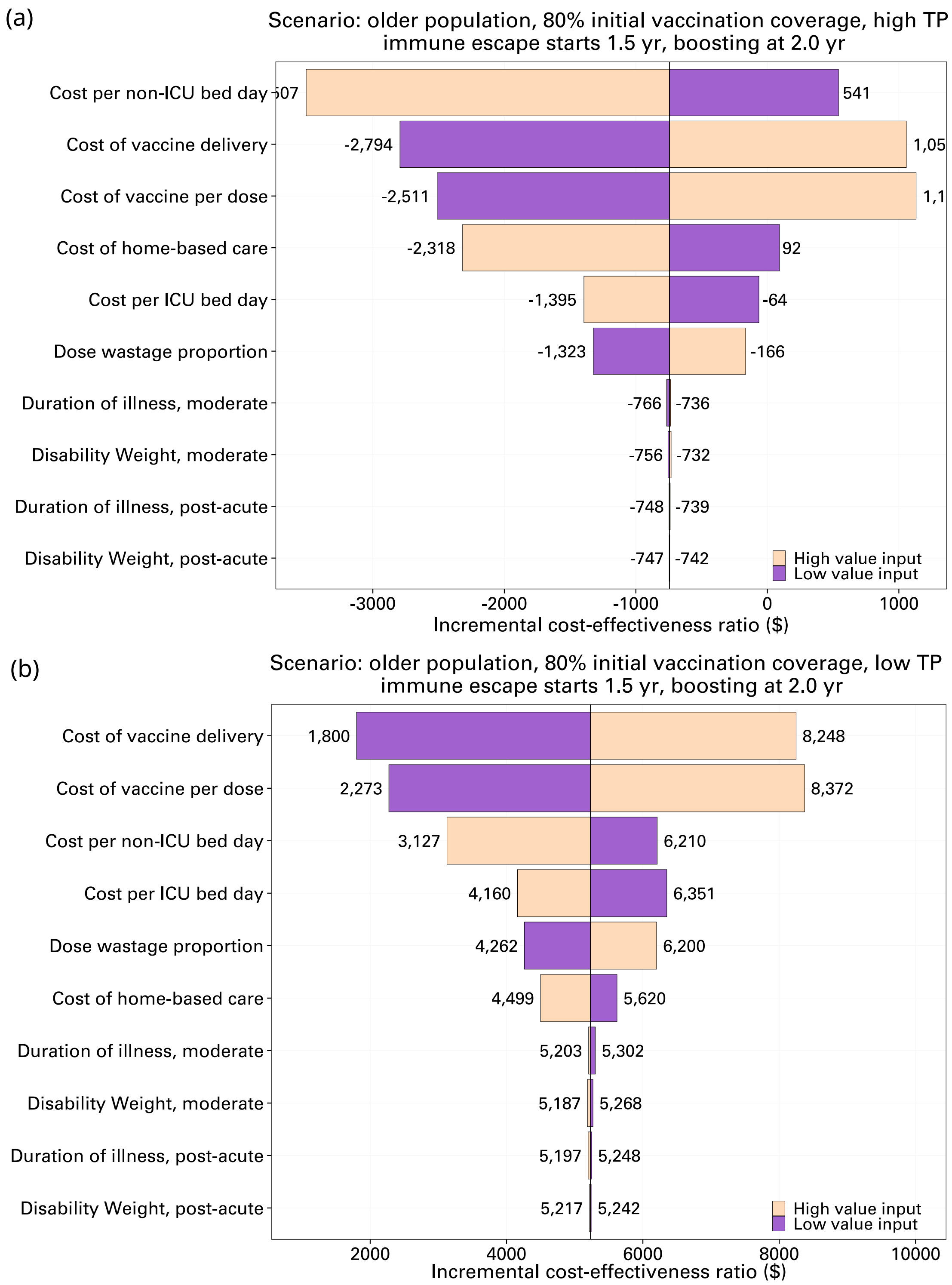

**Figure E5:** One-way sensitivity analysis for high-risk boosting at 2.0 years in an older population with high vaccination coverage. Here, immune escape occurs at 1.5 years. **(a)** in the high transmission potential scenario; **(b)** in the low transmission potential scenario. See Table D4 for the parameter ranges.

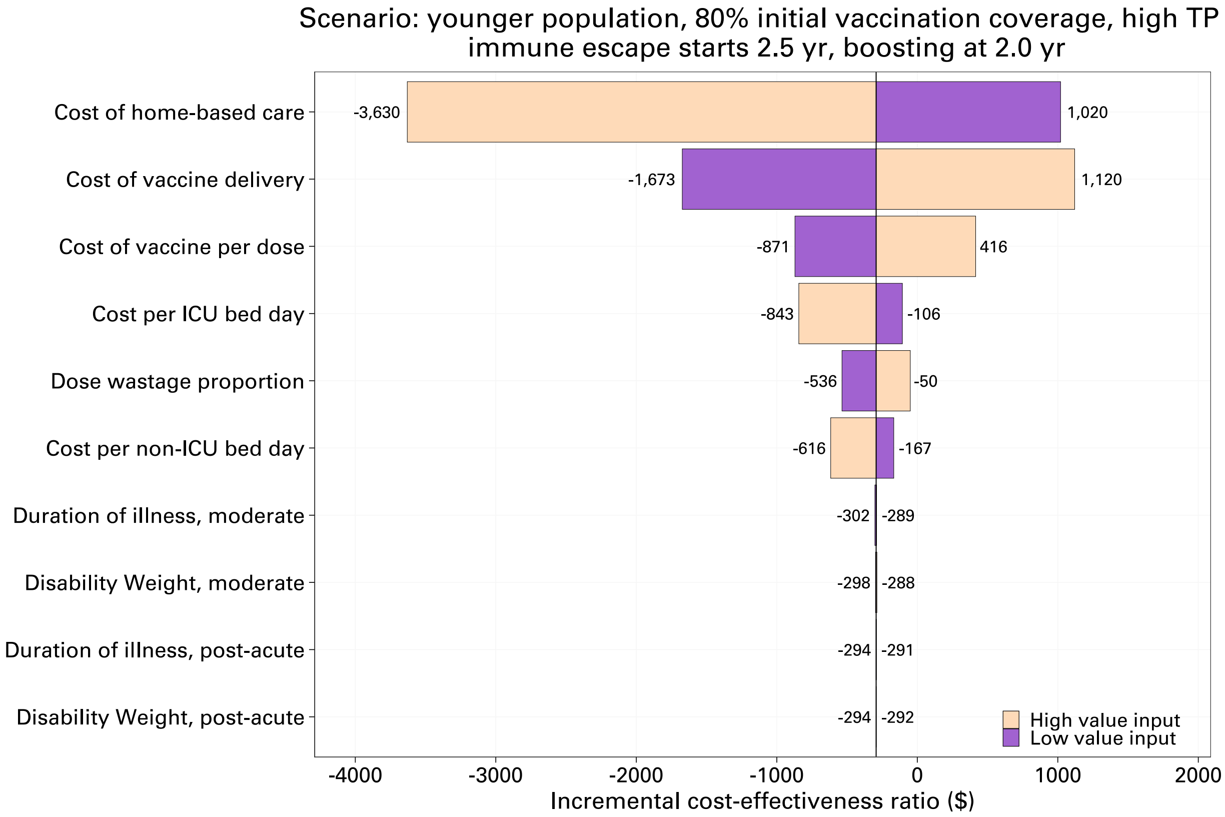

**Figure E6:** One-way sensitivity analysis for high-risk boosting at 2.0 years in a younger population with high transmission potential and high vaccination coverage. Here, immune escape occurs at 2.5 years. See Table D4 for the parameter ranges.

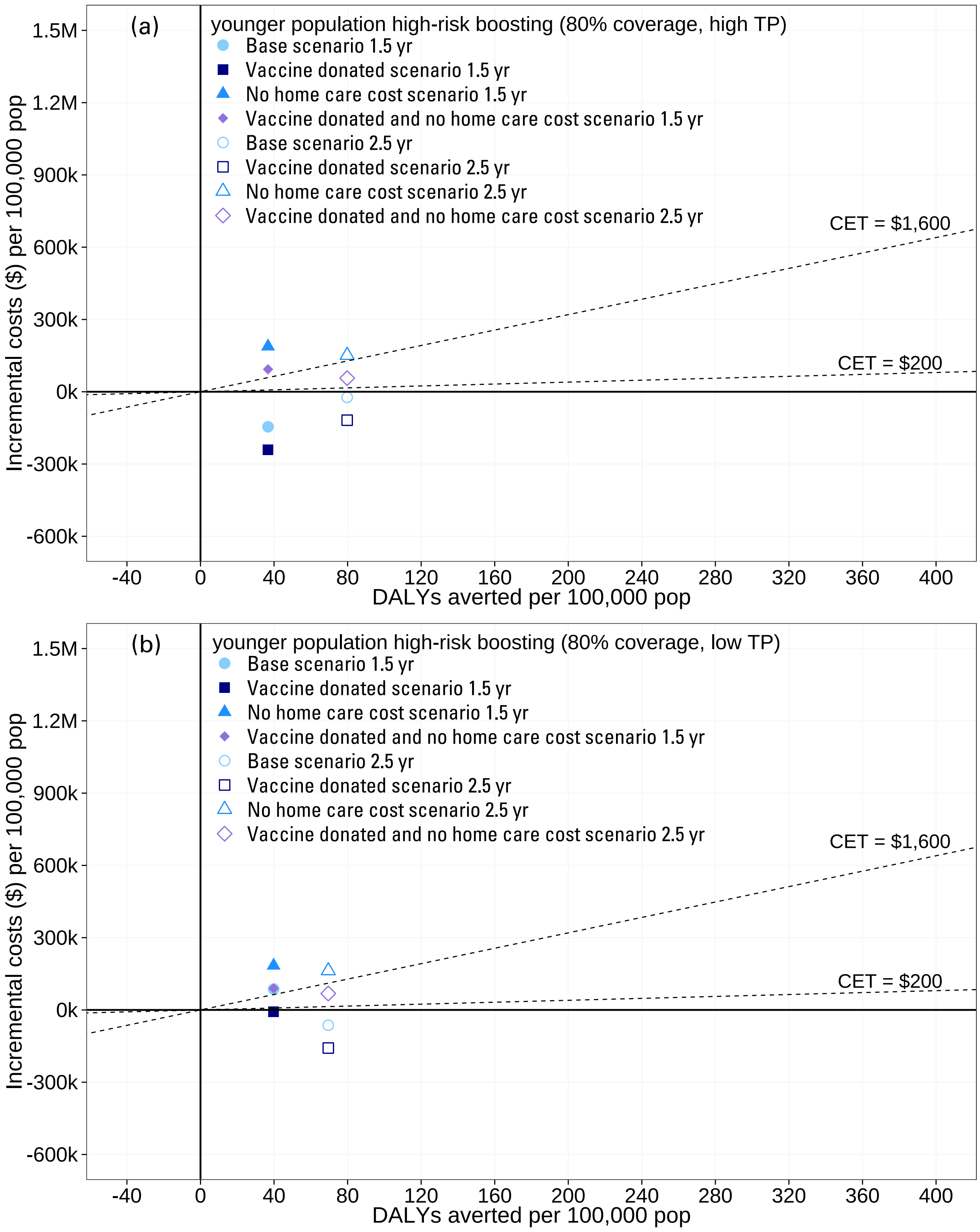

**Figure E7:** Cost-effectiveness analysis of high-risk boosting in the high vaccination coverage setting, for younger demographics, given different cost settings. **(a)** High transmission setting; **(b)** low transmission setting. Scenarios are run with boosting at 2 years. The dotted lines represent cost effective thresholds.

#### E.2 High vaccination coverage scenarios: Boosting frequency - extended results

This set of scenarios (Section 3.1.2 in the main paper) considered timing of boosting, and compared a once-off boosting with a half-yearly boosting schedule. Table E3 shows the median deaths (clinical outcomes), where we see a clear reduction in deaths under a half-yearly boosting schedule as compared to other boosting strategies. Figure E8 shows the cost-effectiveness analysis: high-risk half-yearly boosting is cost-effective in older populations, though it is more expensive than boosting only once. Meanwhile, Figure E9 shows one-way sensitivity analyses for the half-yearly boosting scenarios, showing that home-based care cost is the most influential costing parameter in younger populations.

**Table E3:** Median deaths (with 0.025 and 0.975 quantiles) between 1.5–3 years in high transmission settings with high vaccination coverage, comparing the boosting timing with half-yearly boosting. Early immune escape occurs at 1.5 years; late immune escape occurs at 2.5 years.

| | Population | Immune escape | Boosting timing | Median deaths |
|---|---|---|---|---|
| (a) | older | early escape | 1.75 years | 35.0 (24.0, 47.0) |
| | older | early escape | 2.0 years | 34.0 (23.0, 46.0) |
| | older | early escape | 2.25 years | 36.0 (25.0, 48.0) |
| | older | early escape | 2.5 years | 40.0 (28.0, 52.0) |
| | older | early escape | half-yearly | 32.0 (21.0, 42.0) |
| (b) | younger | early escape | 1.75 years | 9.0 (4.0, 15.0) |
| | younger | early escape | 2.0 years | 9.0 (4.0, 15.0) |
| | younger | early escape | 2.25 years | 9.0 (4.0, 16.0) |
| | younger | early escape | 2.5 years | 10.0 (5.0, 17.0) |
| | younger | early escape | half-yearly | 8.0 (3.0, 14.0) |
| (c) | older | late escape | 1.75 years | 35.0 (24.0, 47.0) |
| | older | late escape | 2.0 years | 34.0 (23.0, 47.0) |
| | older | late escape | 2.25 years | 34.0 (23.0, 47.0) |
| | older | late escape | 2.5 years | 36.0 (25.0, 50.0) |
| | older | late escape | half-yearly | 26.0 (17.0, 37.0) |
| (d) | younger | late escape | 1.75 years | 8.0 (3.0, 15.0) |
| | younger | late escape | 2.0 years | 9.0 (4.0, 16.0) |
| | younger | late escape | 2.25 years | 8.0 (3.0, 15.0) |
| | younger | late escape | 2.5 years | 9.0 (4.0, 15.0) |
| | younger | late escape | half-yearly | 4.0 (1.0, 9.0) |

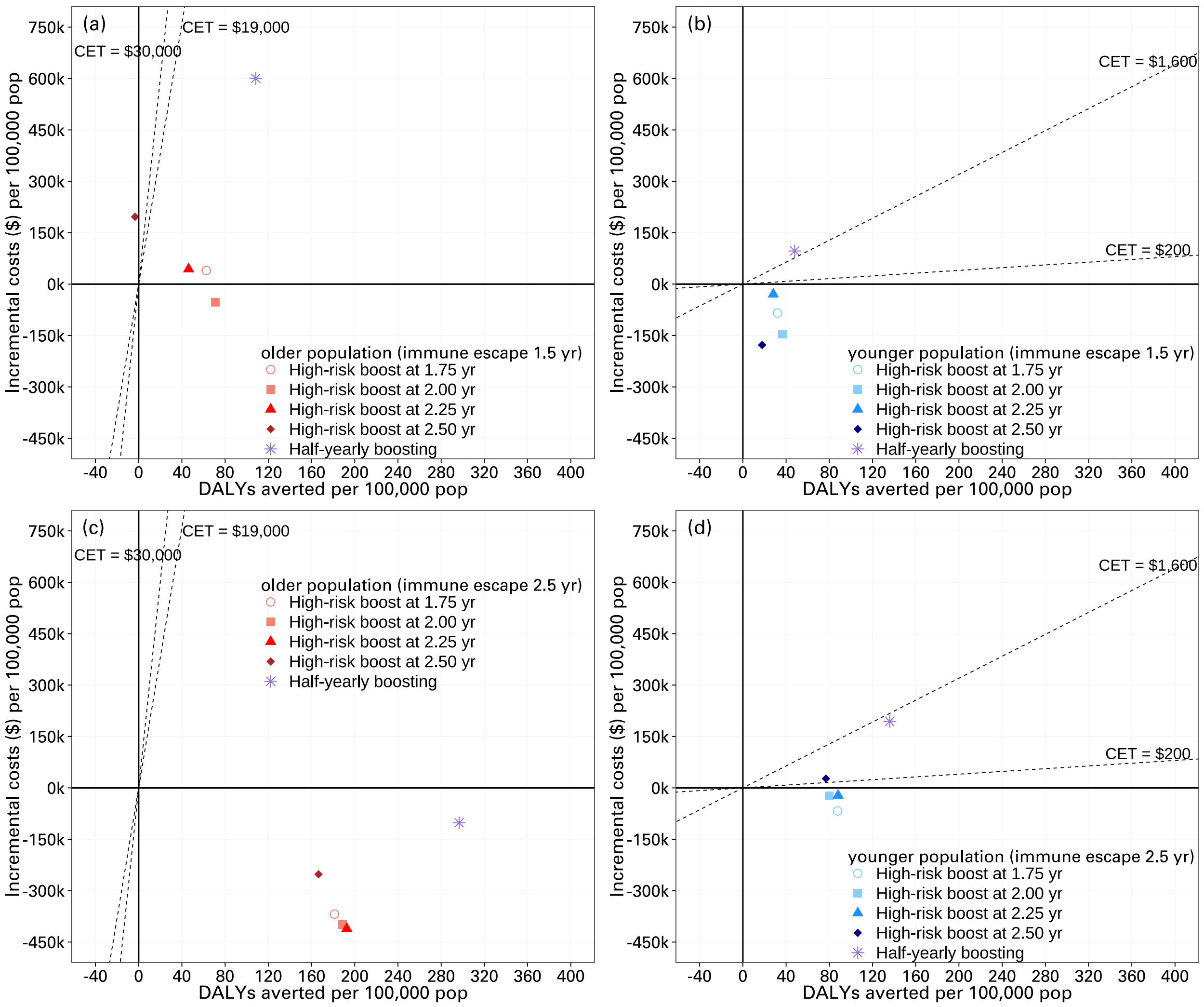

**Figure E8:** Cost-effectiveness analyses comparing frequent boosting and boosting once at a range of times in the high-transmission high-vaccination coverage settings, for older and younger demographics. **(a)** older population with early immune escape (1.5 years); **(b)** younger population with early immune escape (1.5 years); **(c)** older population with late immune escape (2.5 years); **(d)** younger population with late immune escape (2.5 years). The high-risk boosting (65+ in the older population, 55+ in the younger population) is rolled out at either 1.75 years, 2.0 years, 2.25 years, 2.5 years, or half-yearly starting from 1.75 years. High-risk boosting is likely to be cost-effective, though more frequent boosting may not be cost-effective in younger populations.

(a)

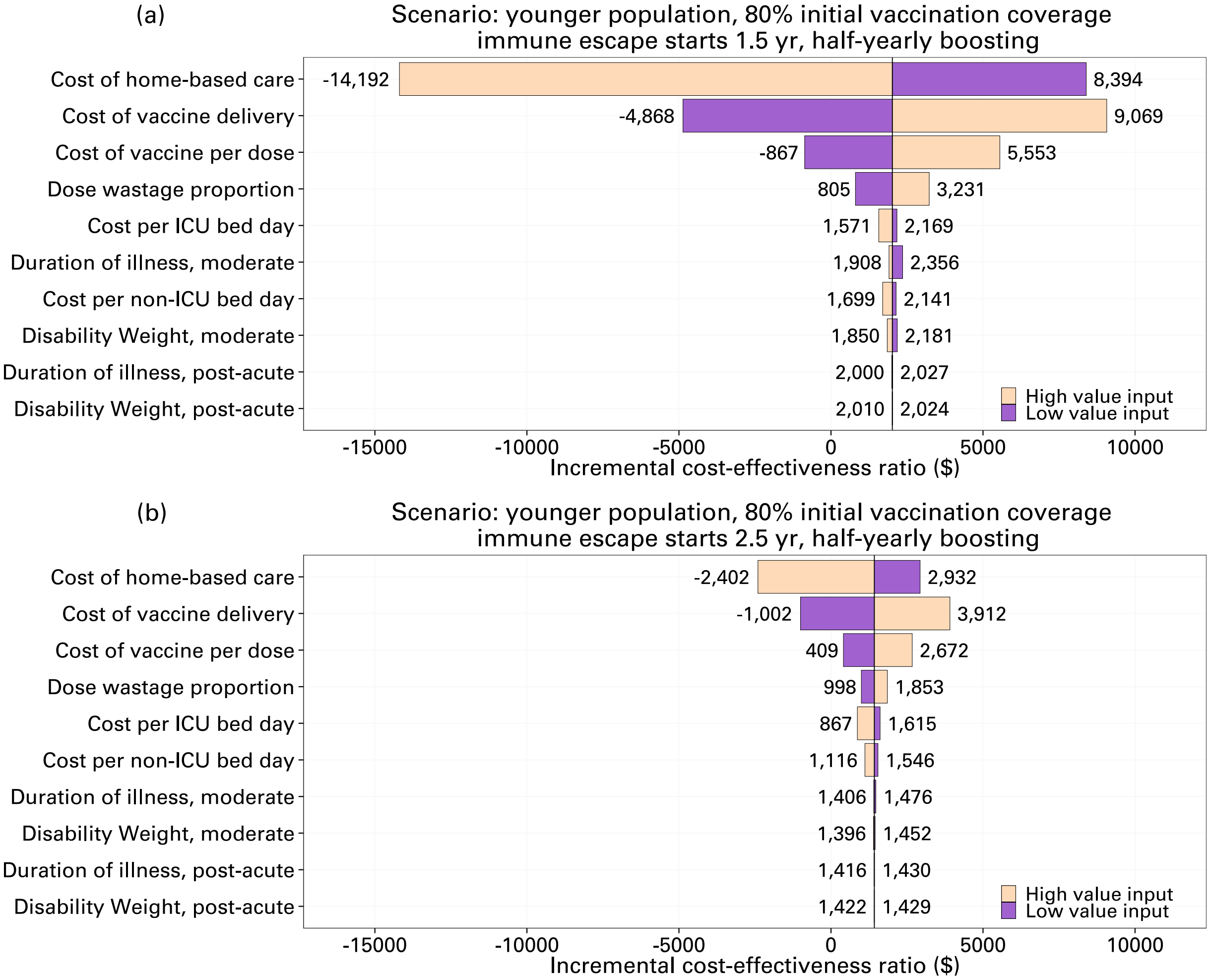

**Figure E9:** One-way sensitivity analysis for the half-yearly high-risk boosting strategy, in a younger population with high transmission potential and high vaccination coverage. **(a)** With early immune escape at 1.5 years; **(b)** with late immune escape at 2.5 years. See Table D4 for the parameter ranges.

#### E.3 High vaccination coverage scenarios: age-cutoff for cost-effective boosting - extended results

This set of scenarios (Section 3.1.3 in the main paper) considered systematically reducing the "high risk boosting" age threshold from 65+ downwards. Figure E10 shows the epidemic infection curves in a low-transmission setting, and Figure E11 shows the cost-effectiveness of different age-thresholds. Overall, the relative results are similar to that of the high-transmission setting, i.e. that boosting 45+ is cost-effective, and there are only minimal gains from decreasing the age threshold down further. Table E4 provides the median deaths in both high transmission and low transmission settings for completeness.

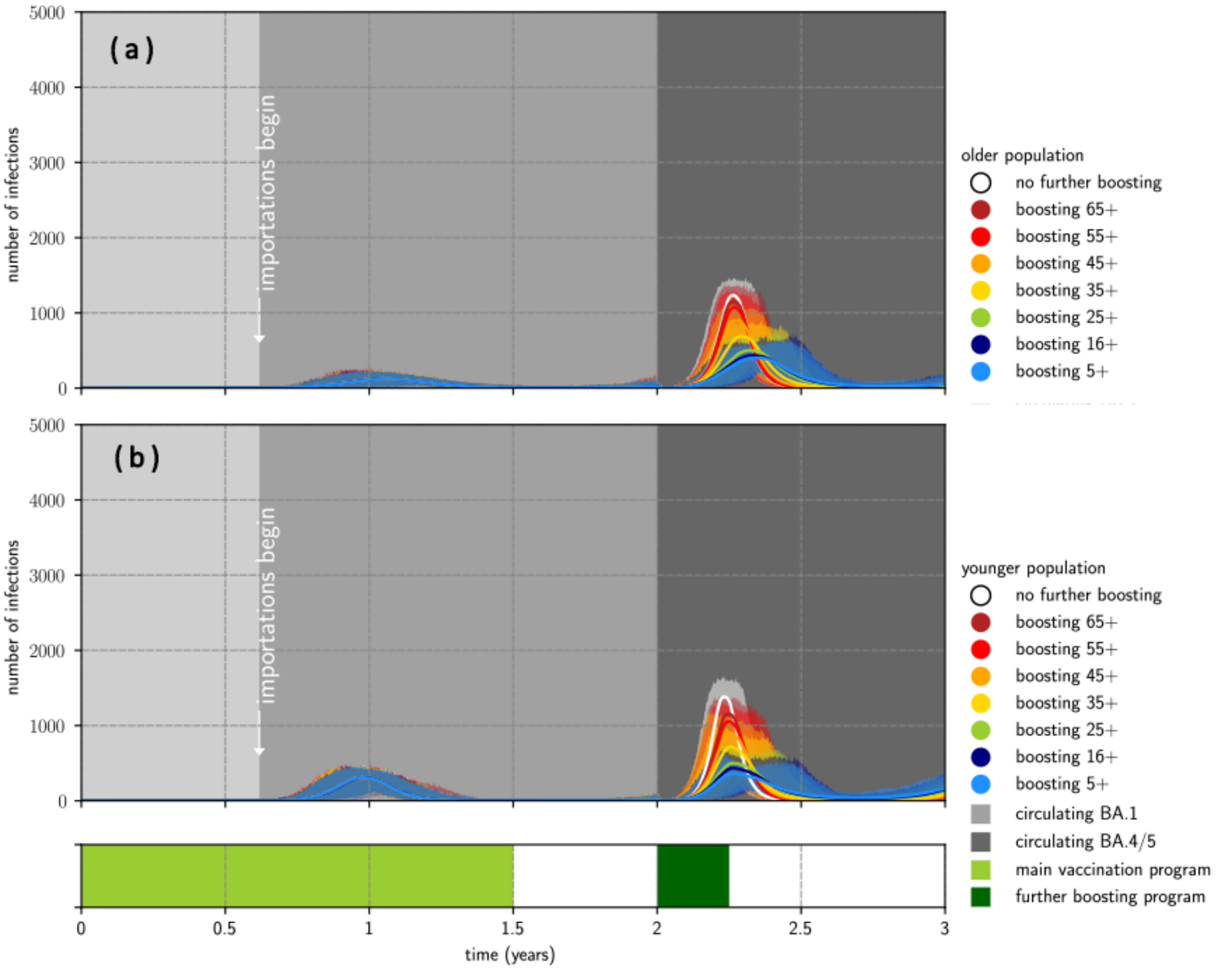

**Figure E10:** Outbreaks in the low transmission, high vaccination coverage setting, for older and younger demographics, comparing the impact of lowering the age cut-off for high risk boosting. **(a)** epidemic waves in the older population; **(b)** epidemic waves in the younger population. All scenarios here had an immune escape variant seeded at 2 years, with boosting at 2 years. The solid lines represent the pointwise median infections from 1000 simulations and the shaded regions represent the pointwise maximum and minimum infections.

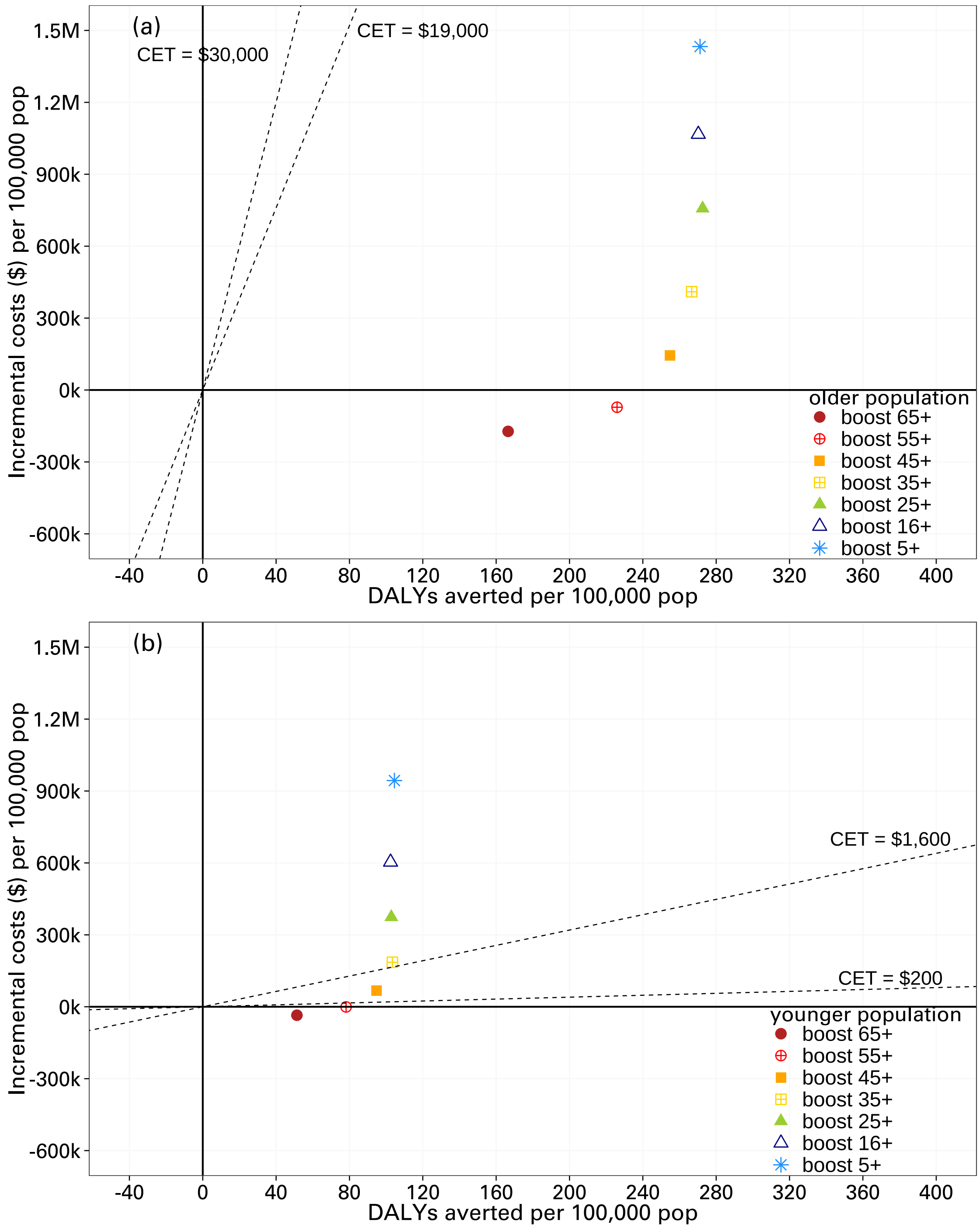

**Figure E11:** Cost-effectiveness analysis of vaccination in the low transmission, high vaccination coverage setting, for older and younger demographics, comparing the impact of lowering the age cut-off for high risk boosting. **(a)** older population; **(b)** younger population. All scenarios here had an immune escape variant seeded at 2 years, with boosting at 2 years. Boosting 65+ and 55+ is likely to be cost-effective or cost-saving.

**Table E4:** Median deaths (with 0.025 and 0.975 quantiles) between 1.5–3 years in the high vaccination coverage setting, for older and younger demographics, comparing the impact of lowering the age cut-off for high risk boosting. All scenarios here had an immune escape variant seeded at 2 years, with boosting at 2 years.

| | Population | Transmission potential | Age group | Median deaths |
|---|---|---|---|---|
| (a) | older | high TP | no further boosting | 47.0 (35.0, 60.0) |
| | older | high TP | boosting 65+ | 34.0 (24.0, 45.0) |
| | older | high TP | boosting 55+ | 31.0 (21.0, 42.0) |
| | older | high TP | boosting 45+ | 29.0 (19.0, 41.0) |
| | older | high TP | boosting 35+ | 30.0 (20.0, 42.0) |
| | older | high TP | boosting 25+ | 28.0 (18.0, 40.0) |
| | older | high TP | boosting 16+ | 28.0 (18.0, 38.0) |
| | older | high TP | boosting 5+ | 29.0 (19.0, 39.0) |
| (b) | younger | high TP | no further boosting | 15.0 (8.0, 23.0) |
| | younger | high TP | boosting 65+ | 12.0 (6.0, 20.0) |
| | younger | high TP | boosting 55+ | 11.0 (5.0, 18.0) |
| | younger | high TP | boosting 45+ | 10.0 (5.0, 18.0) |
| | younger | high TP | boosting 35+ | 10.0 (4.0, 17.02) |
| | younger | high TP | boosting 25+ | 10.0 (4.0, 17.0) |
| | younger | high TP | boosting 16+ | 10.0 (4.0, 16.0) |
| | younger | high TP | boosting 5+ | 10.0 (4.0, 17.0) |
| (c) | older | low TP | no further boosting | 33.0 (22.0, 46.0) |
| | older | low TP | boosting 65+ | 21.0 (12.0, 31.0) |
| | older | low TP | boosting 55+ | 17.0 (10.0, 26.0) |
| | older | low TP | boosting 45+ | 16.0 (8.98, 25.0) |
| | older | low TP | boosting 35+ | 15.0 (8.0, 24.0) |
| | older | low TP | boosting 25+ | 15.0 (8.0, 23.0) |
| | older | low TP | boosting 16+ | 15.0 (8.0, 24.0) |
| | older | low TP | boosting 5+ | 15.0 (8.0, 24.0) |
| (d) | younger | low TP | no further boosting | 15.0 (8.0, 23.0) |
| | younger | low TP | boosting 65+ | 10.0 (4.0, 17.0) |
| | younger | low TP | boosting 55+ | 8.0 (3.0, 15.0) |
| | younger | low TP | boosting 45+ | 7.0 (3.0, 13.0) |
| | younger | low TP | boosting 35+ | 7.0 (2.0, 13.0) |
| | younger | low TP | boosting 25+ | 7.0 (2.0, 13.0) |
| | younger | low TP | boosting 16+ | 7.0 (2.0, 13.0) |
| | younger | low TP | boosting 5+ | 7.0 (2.0, 13.0) |

#### E.4 Low-medium vaccination coverage: comparing primary and booster strategies - extended results

This set of scenarios (Section 3.2.1 in the main paper) considered the trade-off between new primary vaccination and high risk boosting strategies in younger populations with low or medium vaccination coverage. Figure E12 shows the epidemic infection curves in a low-transmission setting, Figure E13 shows the cost-effectiveness of different strategies for both high and low transmission settings, and Table E5 provides the median deaths in high and low transmission settings. The results show that high-risk boosting may be cost-effective in medium vaccination coverage settings. Finally, Figure E14 shows a cost-effectiveness analysis of high-risk boosting in the low vaccination coverage setting given different cost settings, including the scenario where vaccines are donated, or if there are no home care costs. We find that high-risk boosting would be more cost-effective or even cost-saving if vaccines are donated.

**Table E5:** Median deaths (with 0.025 and 0.975 quantiles) between 1.5–3 years in the low-medium vaccination coverage setting, for younger demographics, comparing primary and booster strategies. All scenarios had an immune escape variant seeded at 2 years, with additional vaccination or boosting at 2 years.

| | Transmission potential | Vaccine coverage | Vaccination strategy | Median deaths |
|---|---|---|---|---|
| (a) | high TP | low | no further boosting | 16.0 (9.0, 24.0) |
| | high TP | low | new pediatric vaccination (ages 5–15) | 15.0 (8.0, 24.0) |
| | high TP | low | high-risk boosting (65+ first) | 12.0 (5.0, 19.0) |
| | high TP | low | new primary vaccinations | 14.0 (7.0, 22.0) |
| (b) | high TP | medium | no further boosting | 14.0 (7.0, 21.0) |
| | high TP | medium | new pediatric vaccination (ages 5–15) | 14.0 (7.0, 21.0) |
| | high TP | medium | high-risk boosting (65+ first) | 10.0 (5.0, 16.0) |
| | high TP | medium | new primary vaccinations | 13.0 (7.0, 21.0) |
| (c) | low TP | low | no further boosting | 16.0 (9.0, 24.0) |
| | low TP | low | new pediatric vaccination (ages 5–15) | 16.0 (9.0, 24.0) |
| | low TP | low | high-risk boosting (65+ first) | 11.0 (5.0, 17.0) |
| | low TP | low | new primary vaccinations | 14.0 (8.0, 22.0) |
| (d) | low TP | medium | no further boosting | 16.0 (9.0, 25.0) |
| | low TP | medium | new pediatric vaccination (ages 5–15) | 16.0 (9.0, 24.0) |
| | low TP | medium | high-risk boosting (65+ first) | 8.0 (3.0, 15.0) |
| | low TP | medium | new primary vaccinations | 15.0 (8.0, 24.0) |

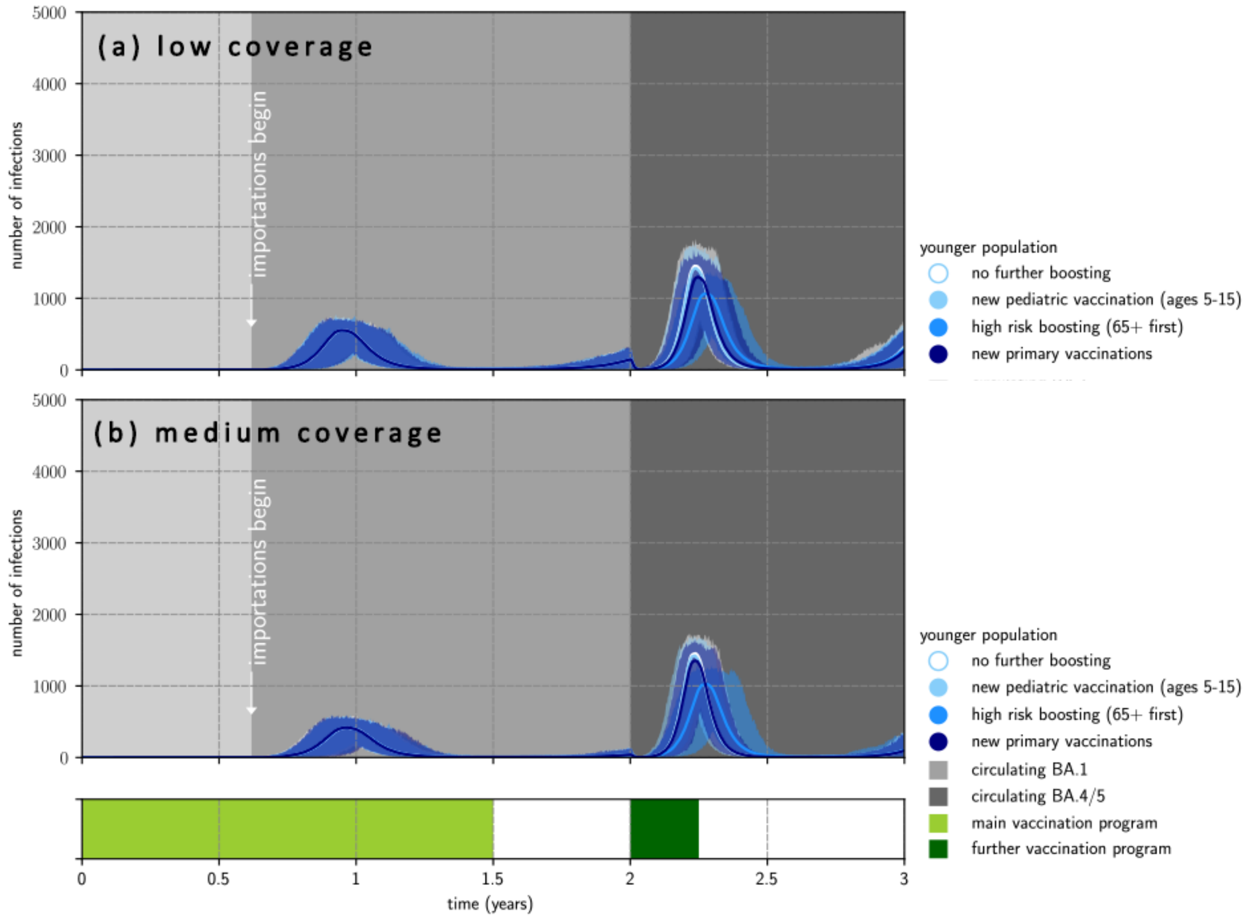

**Figure E12:** Outbreaks in the low transmission, low- and medium-vaccination coverage settings, for younger demographics. **(a)** epidemic waves in a younger population with low vaccination coverage (initial coverage 20%); **(b)** with medium vaccination coverage (initial coverage 50%). All scenarios had an immune escape variant seeded at 2 years, with additional vaccination or boosting at 2 years. The solid lines represent the pointwise median infections from 1000 simulations and the shaded regions represent the pointwise maximum and minimum infections.

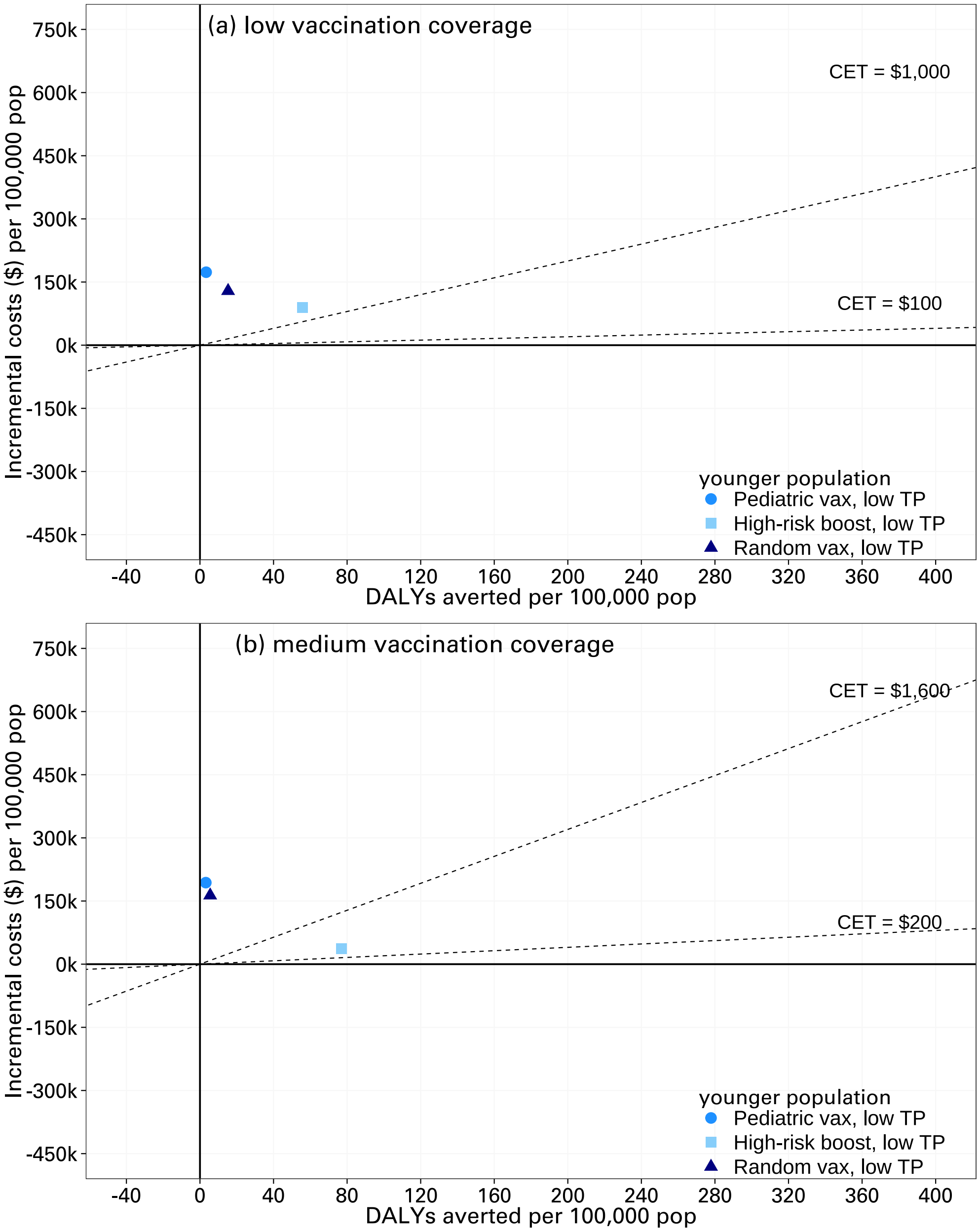

**Figure E13:** Cost-effectiveness analysis of vaccination in low transmission, low- and medium-vaccination coverage settings, for younger demographics. **(a)** cost-effectiveness analysis in a younger population with low vaccination coverage (initial coverage 20%); **(b)** with medium vaccination coverage (initial coverage 50%). All scenarios had an immune escape variant seeded at 2 years, with additional vaccination or boosting at 2 years.

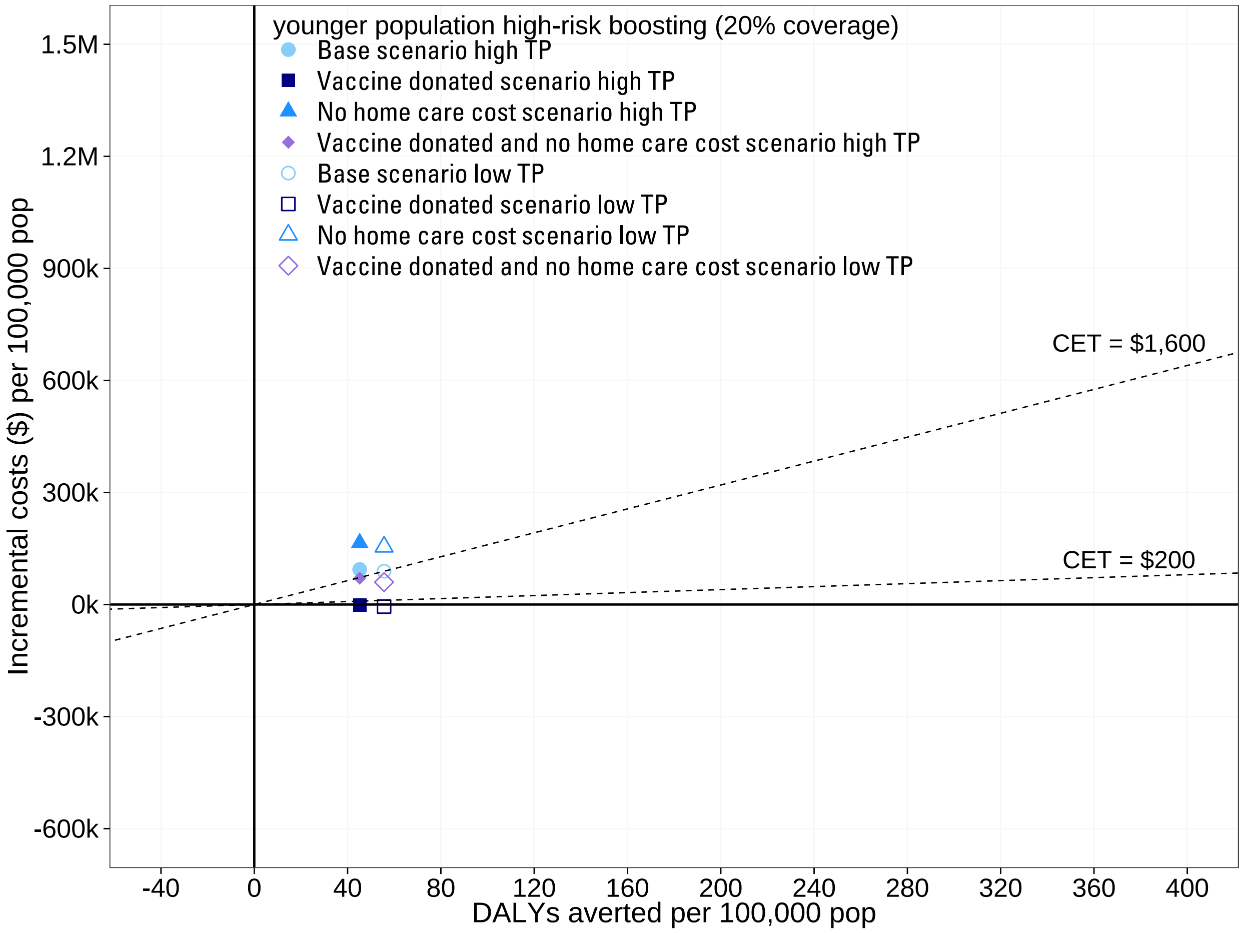

**Figure E14:** Cost-effectiveness analysis of high-risk boosting in the low vaccination coverage setting, for younger demographics, given different cost settings. Scenarios are run with boosting at 2 years. The dotted lines represent cost effective thresholds.

#### E.5 Low-medium vaccination coverage: impact of bivalent boosting - extended results

This set of scenarios (Section 3.2.2 in the main paper) considered the impact of bivalent boosting over monovalent boosting, in younger populations with low or medium vaccination coverage.

Figure E15 shows the epidemic infection curves in the high-transmission setting, while Figure E16 shows the epidemic infection curves in the low-transmission setting. Table E6 shows the median deaths in both transmission settings. We find that there is a slightly larger benefit of bivalent boosters over monovalent boosters in the ow transmission potential scenario compared to the high-transmission scenarios.

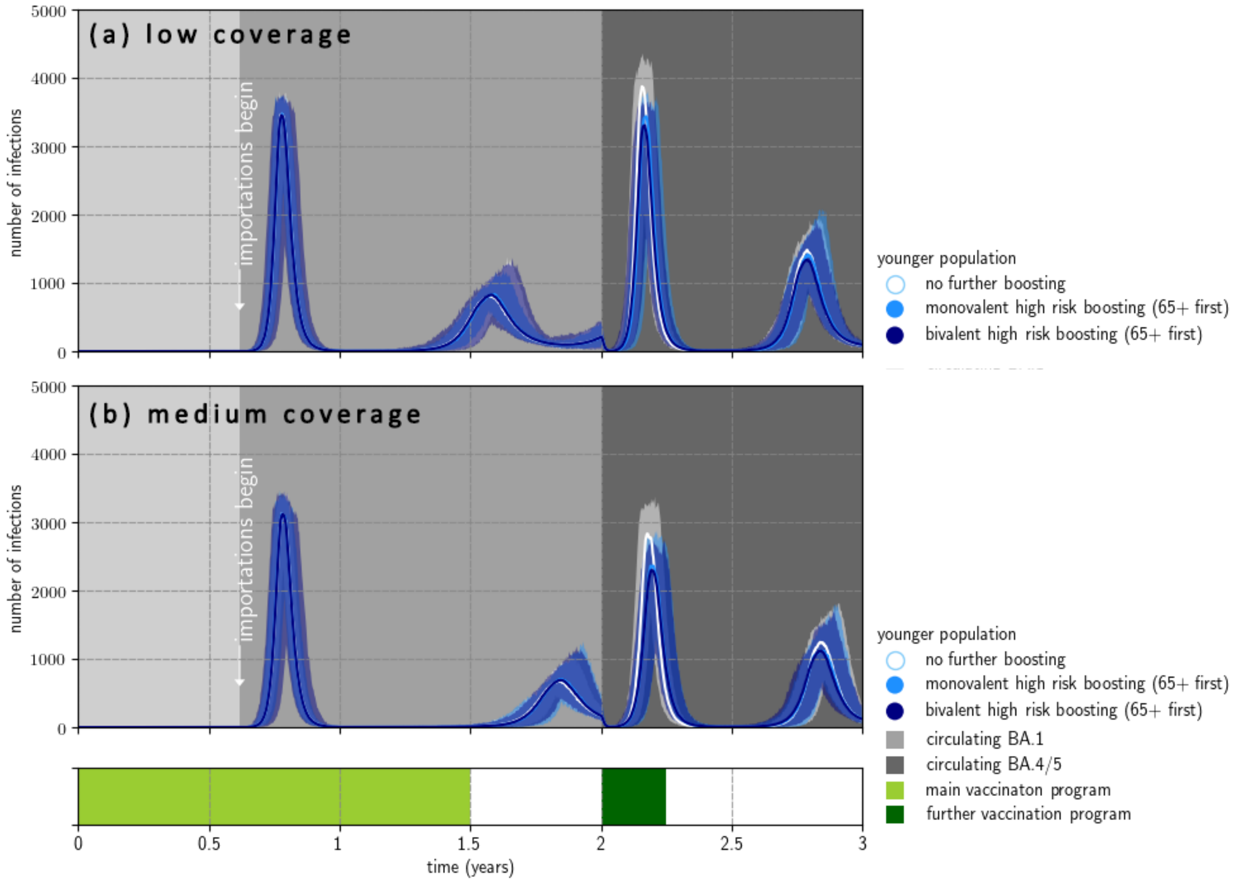

**Figure E15:** Outbreaks in high transmission settings with low and medium vaccination coverage in younger demographics, comparing monovalent vs bivalent boosting. **(a)** low vaccination coverage (initial coverage 20%); **(b)** medium vaccination coverage (initial coverage 50%). All scenarios had an immune escape variant seeded at 2 years, and had high-risk boosting at 2 years. The solid lines represent the pointwise median infections from 1000 simulations and the shaded regions represent the pointwise maximum and minimum infections. Bivalent high risk boosting gives a slight decrease in total numbers of infections, independent of vaccination coverage.

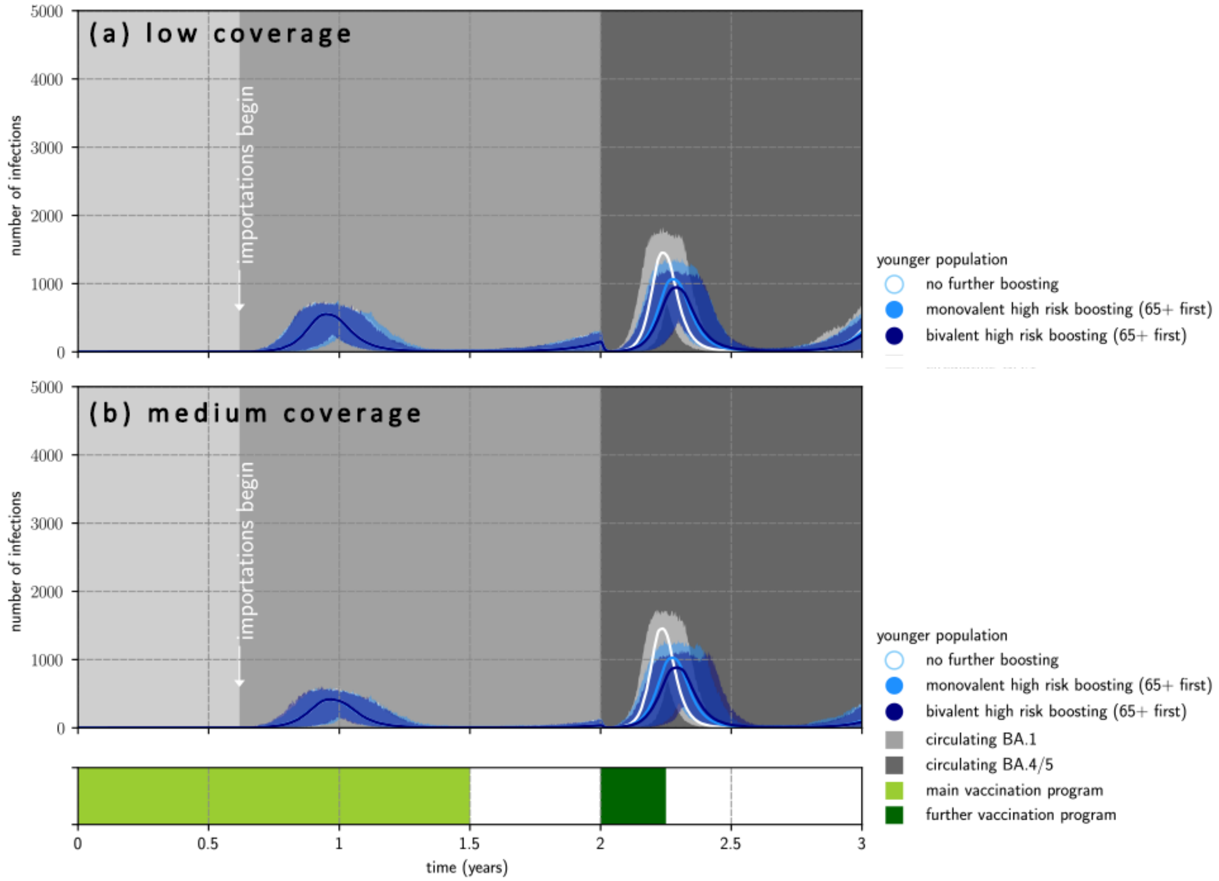

**Figure E16:** Outbreaks in the low transmission, low- and medium-vaccination coverage settings, for younger demographics, comparing monovalent vs bivalent boosting. **(a)** epidemic waves in a younger population with low vaccination coverage (initial coverage 20%); **(b)** with medium vaccination coverage (initial coverage 50%). All scenarios had an immune escape variant seeded at 2 years, with additional high-risk boosting at 2 years. The solid lines represent the pointwise median infections from 1000 simulations and the shaded regions represent the pointwise maximum and minimum infections.

**Table E6:** Median deaths between 1.5–3 years in the low and medium coverage scenarios for younger demographics, comparing high risk booster strategies using monovalent and bivalent vaccines, with 0.025 and 0.975 quantiles. Each scenario has immune escape at 2.0 years, with boosting at 2.0 years.

| | **Transmission potential** | **Vaccine coverage** | **Boosting strategy** | **Median deaths** |
|---|---|---|---|---|
| (a) | high TP | low | no further boosting | 16.0 (9.0, 24.0) |
| | high TP | low | monovalent high-risk boosting | 12.0 (5.0, 19.0) |
| | high TP | low | bivalent high-risk boosting | 11.0 (5.0, 18.0) |
| (b) | high TP | medium | no further boosting | 14.0 (7.0, 21.0) |
| | high TP | medium | monovalent high-risk boosting | 10.0 (5.0, 16.0) |
| | high TP | medium | bivalent high-risk boosting | 10.0 (4.0, 16.0) |
| (c) | low TP | low | no further boosting | 16.0 (9.0, 24.0) |
| | low TP | low | monovalent high-risk boosting | 11.0 (5.0, 17.0) |
| | low TP | low | bivalent high-risk boosting | 9.0 (4.0, 16.0) |
| (d) | low TP | medium | no further boosting | 16.0 (9.0, 25.0) |
| | low TP | medium | monovalent high-risk boosting | 8.0 (3.0, 15.0) |
| | low TP | medium | bivalent high-risk boosting | 7.0 (3.0, 13.0) |

[28] Japanese Government. The National Treasury's burden for the vaccination measures against COVID-19 report.

[35] Australian Government. Medicare Benefits Schedule Book Operating from 21 July 2020. In: Department of Health, 2020.

[41] Global Burden of Disease Collaborative Network. Global Burden of Disease Study 2019 (GBD 2019) Disability Weights. *Seattle, USA: Institute for Health Metrics and Evaluation (IHME)*, 2020.
